# Adjunctive ruxolitinib attenuates inflammation and enhances antiparasitic immunity in human volunteers experimentally infected with *Plasmodium falciparum*

**DOI:** 10.1101/2025.03.26.25324737

**Authors:** Rebecca Webster, Damian A. Oyong, Azrin N. Abd-Rahman, Adam J. Potter, Reena Mukhiya, Nischal Sahai, Indika Leelasena, Eniko Ujvary, Sue Mathison, Dean W. Andrew, Luzia Bukali, Fabian de Labastida Rivera, Jessica Engel, Megan S.F. Soon, Teija Frame, Julianne Hamelink, Mayimuna Nalubega, Nicholas L. Dooley, Jessica R. Loughland, Tran Nguyen, Yael Rosenberg-Hasson, Sofia Maysel-Auslender, Natalia Sigal, Kira Foygel, Jeremy Gower, Jenny Peters, Ria Woo, Fiona Amante, Timothy N.C. Wells, Stephan Chalon, Joerg J. Moehrle, James S. McCarthy, Geoffrey W. Birrell, Michael D. Edstein, Michael Leipold, Gerlinde Obermoser, Holden Maecker, Christian R. Engwerda, Bridget E Barber, Michelle J. Boyle

**Affiliations:** QIMR Berghofer, Brisbane, QLD, Australia; Burnet Institute, Melbourne, VIC, Australia; Griffith University, Brisbane, Australia; University of Sunshine Coast Clinical Trials, Brisbane, QLD, Australia; University of Queensland, Brisbane, QLD, Australia; Stanford University, Stanford, CA, USA; Medicines for Malaria Venture, Geneva, Switzerland; Peter Doherty Institute for Infection and Immunity, Melbourne, VIC, Australia; Australian Defence Force Malaria and Infectious Disease Institute, Brisbane, QLD, Australia; Royal Brisbane & Women’s Hospital, Brisbane, QLD, Australia; Menzies School of Health Research, Darwin, NT, Australia; University of Basel, Basel, Switzerland; Swiss Tropical and Public Health Institute, Allschwil, Switzerland

## Abstract

Inhibiting the host inflammatory response to malaria represents a potential strategy to improve clinical outcomes and inhibit immunoregulatory pathways that underlie suboptimal development of antiparasitic immunity. Ruxolitinib is a JAK 1/2 inhibitor that reduces inflammatory biomarkers when used in myeloproliferative disorders, and inhibits type-1 interferons and enhances CD4+ T cell immunity when combined with anti-parasitic drugs in animal models. Here we report the results of a double-blind randomised placebo-controlled trial evaluating the ability of ruxolitinib to reduce inflammatory responses and boost anti-parasitic immunity in malaria-naïve volunteers inoculated with blood-stage *Plasmodium falciparum*. Twenty participants were inoculated, and randomized on day 8 to receive artemether-lumefantrine with either ruxolitinib or placebo. Ninety days later, participants who remained eligible were re-inoculated with a second infection. Ruxolitinib was safe and well-tolerated, and attenuated the host inflammatory response to the initial infection, with reduced post-treatment increases in the inflammatory biomarker CRP, as well as markers of disease severity including angiopoietin-2 and ICAM-1. Further, ruxolitinib enhanced the immune memory response following a second inoculation, with increased plasma levels of HLA-DR and CXCL13, indicating enhanced immune activation and germinal centre responses, respectively. These data support the further evaluation of ruxolitinib as an adjunctive treatment to improve clinical outcomes and boost anti-parasitic immunity in clinical malaria.

## Introduction

Malaria remains a major public health challenge, with 263 million cases and 597,000 deaths reported globally in 2023, the majority from *Plasmodium falciparum* [1]. Current efforts to control malaria have stalled, and case-fatality from severe malaria remains high despite the use of highly effective anti-parasitic drug treatments [2]. While the recent introduction of two malaria vaccines offers promise, efficacy and duration of protection is suboptimal, particularly in populations with the highest malaria burdens [3–5]. Thus, novel strategies are urgently needed to improve clinical outcomes and enhance the effectiveness of current vaccines.

Most symptoms of clinical malaria arise from the inflammatory response to the release of parasites from red blood cells (RBCs), prior to re-invasion of RBCs to continue parasite expansion [6–8]. These inflammatory responses are primarily driven by IFNγ and TNF produced by innate-like lymphocytes such as γδ T cells and NK cells, as well as monocytes, CD4^+^ and CD8^+^ T cells [9–12]. IFNγ and TNF, along with other proinflammatory cytokines, activate vascular endothelium allowing parasitised RBCs to bind via integrins such as ICAM-1, thereby initiating tissue sequestration or accumulation of parasitised RBCs [13]. This process initiates an inflammatory cascade that includes local recruitment of immune cells, damage to vascular and tissue structural integrity and breakdown of blood barriers compromising tissue functions [14]. To counteract these potentially lethal anti-parasitic responses, powerful immune regulatory networks are established by the host to protect tissues [15]. Type I interferons (IFN) play an important role in the initiation of these parasite-specific anti-inflammatory responses via the recruitment of Janus Activated Kinases (JAK) 1 and 2 to cytosolic domains of the type I IFN receptor, leading to phosphorylation of signal transducer and activator of transcription (STAT)1, 2 and 3 [16]. The resulting cell signalling cascade drives development of a number of cell autologous immunoregulatory mechanisms that includes immune cell expression of co-inhibitory receptors, development of atypical B cells, suboptimal T follicular helper (Tfh) cell functions, and Th1 cell responses characterised by autologous interleukin-10 production [17–20].

Although immunoregulatory systems that develop during malaria can protect against inflammation-mediated tissue damage, they may also impede the development of protective anti-parasitic immunity either following natural infection, vaccination or anti-parasitic drug treatment [21, 22]. Additionally, if these regulatory networks are not sufficiently calibrated, they may also fail to prevent tissue-specific pathology [6–8]. Hence, host-directed strategies that either transiently suppress or deviate these responses to allow development of protective anti-parasitic immunity without causing or even preventing immune-mediated pathology have substantial potential as new malaria treatment and control measures [23–25].

Ruxolitinib is a licensed small molecule orally bioavailable JAK1 and JAK2 inhibitor that has been shown to block type I IFN-mediated inhibition of anti-parasitic CD4^+^ T cell responses in visceral leishmaniasis [26]. Ruxolitinib has been approved for the treatment of adults with myelofibrosis, polycythemia vera, and steroid-refractory graft-versus-host disease, and has also been used in children with type-1 interferonopathy [27]. We hypothesised that ruxolitinib may modulate the host immune response to improve protective immunity if administered as an adjunctive treatment in malaria. The safety and absence of significant drug interactions of ruxolitinib administered in combination with the first-line antimalarial artemether-lumefantrine has recently been demonstrated [28]; however, the use of ruxolitinib in individuals with malaria has not been evaluated.

Here, we report the results of a double-blind randomised placebo-controlled trial to evaluate the ability of ruxolitinib to modulate the host immune responses and boost protective immunity when administered alongside artemether-lumefantrine in malaria-naïve volunteers experimentally infected with blood-stage *P. falciparum*. The primary endpoint was safety and tolerability. Secondary endpoints included the impact of ruxolitinib on phosphorylation of STAT3 (pSTAT3), antimalarial drug concentrations, parasite clearance, inflammatory signals and immune responses, and parasite growth following a second homologous *P. falciparum* infection.

## Results

### Study overview and participants

The study was conducted from 9 June 2021 to 7 July 2023. A total of 20 malaria-naïve volunteers were enrolled across 6 cohorts (**Figure 1**). Baseline characteristics of the participants are shown in **Table 1**. Participants were aged between 18 and 55 years, 11 were male (55%), and 17 (85%) self-selected their race as white.

**Figure 1.**
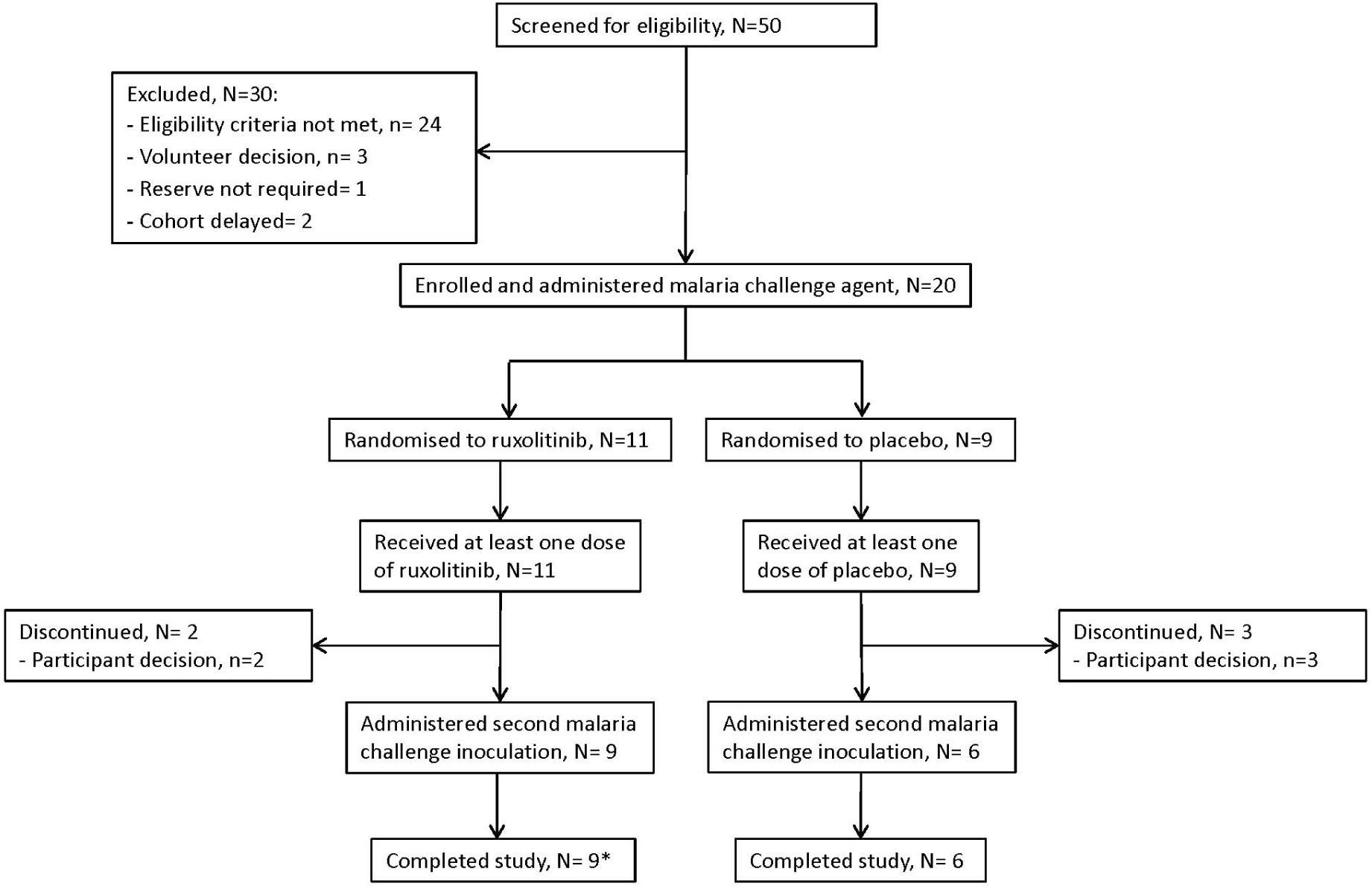
Trial profile. Twenty participants were inoculated with P. falciparum-infected erythrocytes and randomised on the day of dosing to receive artemether-lumefantrine with either ruxolitinib or placebo. All participants received at least one dose of ruxolitinib or placebo. 5 participants, 2 in the ruxolitinib group and 3 in the placebo, discontinued voluntarily prior to administration of a second P. falciparum inoculation. *one participant in the ruxolitinib group tested positive for COVID-19 on day 6 following the second inoculation, necessitating early administration of artemether-lumefantrine.

**Table 1.** Participant characteristics and parasitaemia at time of ruxolitinib/placebo administration.

|  | All (n=20) | AL + ruxolitinib (n=11) | AL + placebo (n=9) |
| --- | --- | --- | --- |
| **Male sex, n (%)** | 11 (55) | 7 (64) | 4 (44) |
| **Age, years (median, range)** | 33 (18 – 55) | 38 (21 – 55) | 27 (18 – 41) |
| **BMI, kg/m2 (median, range)** | 25 (20 – 31) | 26 (21 – 31) | 25 (20 – 29) |
| **Race, n (%)** |  |  |  |
| **White** | 17 (85) | 8 (73) | 9 (100) |
| **Asian** | 1 (5) | 1 (9) | 0 (0) |
| **Other\*** | 2 (10) | 2 (18) | 0 (0) |
| **Parasitaemia, parasites/ml (geometric mean, range)** | 11,307 (363 – 251,802) | 11,084 (968 – 149,520) | 11,586 (363 – 251,802) |

AL = artemether-lumefantrine; BMI: body mass index
\*One participant in the ruxolitinib group reported their race as Columbian, and one as Iraqi

On day 0, participants were intravenously inoculated with approximately 2,800 viable *P. falciparum* 3D7 infected erythrocytes. Parasitemia was monitored daily by qPCR targeting the *P. falciparum* 18S rRNA gene from day 4. On day 8 (cohorts 3 to 6) or 9 (cohorts 1 and 2), participants were randomised to orally receive 20 mg ruxolitinib or placebo, administered 2 hours after artemether-lumefantrine. All study drugs were administered twice daily for 3 days, under direct observation by clinical staff. Participants were confined at the clinical trial site for 72 hours following the 1^st^ dose, and returned as outpatients for follow-up visits until day 28. On day 90, participants still available and eligible were inoculated again with the same dose of *P. falciparum* infected erythrocytes, and treated with artemether-lumefantrine when parasitaemia reached >50,000 parasites/mL, or earlier if other pre-specified clinical criteria were met; participants were again monitored as out-patients for 28 days following administration of artemether-lumefantrine.

Following the first inoculation, parasitaemia progressed as expected, with no difference in pre-treatment parasitemia between treatment groups (**Table 1**). All 20 participants received artemether-lumefantrine and at least one dose of ruxolitinib (n=11) or placebo (n=9) (**Figure 1**) commencing on day 8 or 9. In the ruxolitinib group, 9 participants received all 6 doses as per protocol, while 2 had dosing discontinued due to protocol specified toxicity rules. One of these participants received 3 doses (20 mg twice on day 8 and a single 20 mg dose on day 9) before dosing was discontinued due to a decreased platelet count >50% from baseline. The other participant received 2 doses (20 mg twice on day 8) with dosing discontinued due to a grade 3 reduction in neutrophil count. In the placebo group, 8 participants received all 6 doses as per protocol, and one received 3 doses before dosing was discontinued due to a grade 3 elevation in aspartate aminotransferase (AST) and alanine aminotransferase (ALT).

Five participants (2 in the ruxolitinib group and 3 in the placebo group) voluntarily discontinued the study after the first inoculation phase (**Figure 1**). One other participant (in the ruxolitinib group) tested positive for COVID-19 on day 6 following the second inoculation, necessitating early administration of artemether-lumefantrine.

### Safety

In the first inoculation phase, a total of 126 adverse events (AEs) were reported after ruxolitinib/placebo (investigation medicinal product [IMP]) dosing, with 18/20 (90%) participants experiencing at least one AE (**Table 2**). There was no evidence that the incidence or severity of AEs were different between treatment groups, including those deemed to be IMP related or related to malaria challenge (**Table 2**). Following administration of IMP, participants in the ruxolitinib group experienced a median of 5 AEs per participant (range 1 – 16), compared to a median of 7 AEs per participant (range 1 – 18) in the placebo group (**Table S1**).

**Table 2.** Overall summary of adverse event incidence and severity following ruxolitinib/placebo dosing in the first inoculation phase (Day 8 to Day 89)

|  | **Rux**<br>**(N=11)** |  | **Placebo**<br>**(N=9)** |  | **Total**<br>**(N=20)** |  |
| --- | --- | --- | --- | --- | --- | --- |
|  | **n (%)** | **E** | **n (%)** | **E** | **n (%)** | **E** |
| Any AE | 9 (81.8%) | 51 | 9 (100.0%) | 75 | 18 (90.0%) | 126 |
| Mild | 8 (72.7%) | 35 | 9 (100.0%) | 50 | 17 (85.0%) | 85 |
| Moderate | 7 (63.6%) | 15 | 6 (66.7%) | 21 | 13 (65.0%) | 36 |
| Severe | 1 (9.1%) | 1 | 2 (22.2%) | 4 | 3 (15.0%) | 5 |
| IMP-related AE | 8 (72.7%) | 17 | 5 (55.6%) | 18 | 13 (65.0%) | 35 |
| Mild | 5 (45.5%) | 7 | 4 (44.4%) | 9 | 9 (45.0%) | 16 |
| Moderate | 5 (45.5%) | 9 | 3 (33.3%) | 7 | 8 (40.0%) | 16 |
| Severe | 1 (9.1%) | 1 | 1 (11.1%) | 2 | 2 (10.0%) | 3 |
| Challenge agent related AE | 8 (72.7%) | 37 | 7 (77.8%) | 50 | 15 (75.0%) | 87 |
| Mild | 7 (63.6%) | 23 | 6 (66.7%) | 32 | 13 (65.0%) | 55 |
| Moderate | 6 (54.5%) | 13 | 6 (66.7%) | 14 | 12 (60.0%) | 27 |
| Severe | 1 (9.1%) | 1 | 2 (22.2%) | 4 | 3 (15.0%) | 5 |
| SAE | 0 (0.0%) | 0 | 0 (0.0%) | 0 | 0 (0.0%) | 0 |

n(%): number and percentage of participants experiencing an AE; E:number of AEs

Most AEs were mild (85/126, 68%) or moderate (36/126, 29%) in severity, and most (87/126, 69%) were considered related to the malaria challenge agent (**Table 2**). There were no serious adverse events (SAEs). There were 35 AEs that were considered related to the IMP. Most of these were also considered related to the malaria challenge agent since it was not possible to distinguish the causality. Headache was the most commonly reported IMP-related AE overall, with 7 events reported in 6 participants (3 participants in each treatment group) (**Table 3**). The other IMP-related AEs consisted mostly of laboratory abnormalities, including elevations in liver function enzymes (ALT, AST, GGT), and decreases in white blood cells, platelets and haemoglobin. The majority of IMP-related AEs were mild or moderate in severity (each 16/35 AEs). There were 3 severe (grade 3) IMP-related AEs that resulted in discontinuation of IMP dosing. A concomitant elevation in ALT (456 U/L; normal range 0-45 U/L) and AST (360 U/L; normal range 0-41 U/L) in one participant resulted in discontinuation of dosing after three doses of placebo had been administered. Additionally, a neutrophil count decrease (0.67×10^9^/L; normal range 1.5-8.0×10^9^/L) in another participant resulted in discontinuation of dosing after two doses of ruxolitinib had been administered. A decrease in platelet count resulted in discontinuation of IMP dosing in another participant after three doses of ruxolitinib had been administered. The platelet count decrease (81×10^9^/L; normal range 150-400×10^9^/L) was moderate in severity (grade 2) but met a protocol-specified stopping criterion (platelet count drop of >50% from baseline).

**Table 3.** Ruxolitinib/placebo related adverse events.

| AE preferred term | Rux<br>(N=11) |  | Placebo<br>(N=9) |  | Total<br>(N=20) |  |
| --- | --- | --- | --- | --- | --- | --- |
|  | n (%) | E | n (%) | E | n (%) | E |
| Abdominal discomfort | 1 (9.1%) | 1 | 1 (11.1%) | 1 | 2 (10.0%) | 2 |
| Alanine aminotransferase increased | 1 (9.1%) | 1 | 1 (11.1%) | 1 | 2 (10.0%) | 2 |
| Aspartate aminotransferase increased | 0 (0.0%) | 0 | 1 (11.1%) | 1 | 1 (5.0%) | 1 |
| Gamma-glutamyltransferase increased | 0 (0.0%) | 0 | 1 (11.1%) | 1 | 1 (5.0%) | 1 |
| Haemoglobin decreased | 3 (27.3%) | 3 | 1 (11.1%) | 1 | 4 (20.0%) | 4 |
| Headache | 3 (27.3%) | 3 | 3 (33.3%) | 4 | 6 (30.0%) | 7 |
| Hyperhidrosis | 0 (0.0%) | 0 | 1 (11.1%) | 2 | 1 (5.0%) | 2 |
| Lymphocyte count decreased | 2 (18.2%) | 2 | 2 (22.2%) | 2 | 5 (25.0%) | 5 |
| Nausea | 1 (9.1%) | 1 | 2 (22.2%) | 3 | 3 (15.0%) | 4 |
| Neutrophil count decreased | 2 (18.2%) | 4 | 0 (0.0%) | 0 | 2 (10.0%) | 4 |
| Platelet count decreased | 1 (9.1%) | 1 | 0 (0.0%) | 0 | 1 (5.0%) | 1 |
| White blood cell count decreased | 1 (9.1%) | 1 | 2 (22.2%) | 2 | 3 (15.0%) | 3 |

n(%): number and percentage of participants experiencing an AE; E=:number of AEs.

Safety and tolerability of ruxolitinib following the first inoculation was further evaluated with a malaria clinical score, calculated based on number and severity of malaria-related symptoms and evaluated 3 times per day during confinement (**Table S2**). The sum of the malaria clinical score (sum) during this post-treatment confinement period was associated with pre-treatment parasitemia (r=0.71, p=0.04). The median sum was 7 (range 0 – 38) in the placebo group, compared to 2 (range 0 – 6) in the ruxolitinib group (**Table S3).**

Following the second inoculation, a total of 103 AEs were reported, with all participants experiencing at least one AE (**Table S4**). The median number of AEs experienced per participant following the second inoculation (median 7 [range 1-18]) was comparable to the first inoculation (**Table S1**). There was no evidence of an effect of ruxolitinib on the incidence or severity of AEs experienced during the second inoculation.

### Effect of ruxolitinib on artemether-lumefantrine and metabolite concentrations

Plasma concentrations of ruxolitinib, artemether, dihydroartemisinin (DHA), lumefantrine and desbutyl-lumefantrine (DBL) were quantified by LC MS/MS. In the ruxolitinib treated group, ruxolitinib plasma concentration time profiles showed marked variability between participants (**Figure S1**). Ruxolitinib was rapidly absorbed, with a mean maximum concentration (C_max_) of 192 ng/mL (SD 1.38), a median time to C_max_ of 0.97 hours (range 0.93 – 2.98) after the first dose, and with a short mean elimination half-life of 2.4 hours (SD 1.55) (**Table S5**).

Ruxolitinib reduced artemether absorption, with a mean artemether C_max_ after the first dose of 35.41 ng/mL (SD 1.77) in the ruxolitinib group compared to 67.80 ng/mL (SD 1.68) in the placebo group (p=0.048, **Table S6**). The mean artemether C_max_ was also significantly lower in the ruxolitinib group after the last dose on day 3, as was the artemether area under concentration-time curve (AUC) following the 1^st^ and last dose (**Table S6**). Artemether is quickly hydrolysed to its active metabolite, DHA. Mean C_max_ for DHA was 59.93 ng/mL (SD 1.62) in the placebo group compared to 41.45 ng/mL (SD 1.28) in the ruxolitinib group after the first dose (p=0.095), and 71.05 ng/mL (SD 2.30) and 27.89 ng/mL (SD 2.17) for placebo and ruxolitinib groups, respectively, after the last dose (p=0.019, **Table S6**). The AUC for DHA was also significantly lower in ruxolitinib treated individuals (p=0.005). In contrast to artemether and DHA, ruxolitinib had no impact on concentrations of lumefantrine or DBL (**Table S7)**.

### Parasite clearance following treatment with artemether-lumefantine plus ruxolitinib/placebo

Parasite clearance was evaluated using the parasite reduction ratio over 48 hours (PRR_48_) and the parasite clearance half-life (PC_t1/2_), estimated using the slope of the optimal fit of the log-linear relationship of the parasite decay, as previously described [29]. All 11 participants administered artemether-lumefantrine with ruxolitinib had significant regression models (slope coefficient from the log-linear parasite decay regression) and contributed towards the treatment-specific estimate of PRR_48_ and corresponding PC_t1/2_. However, in the placebo group only 7 of 9 participants had significant regression models and contributed towards the parasite clearance parameter estimates (**Table S8)**. The parasite clearance rate (weighted mean slope of parasitaemia decay) between treatment groups differed significantly (*Q*_*B*_=35.5, p<0.001) (**Table 4)**. However, a sensitivity analysis with removal of one outlying participant from the placebo group who had particularly rapid clearance time (R007) indicated clearance rates between groups were no longer significantly different (*Q*_*B*_=0.56, p=0.45). No individuals in either group experienced recrudescence.

**Table 4.**
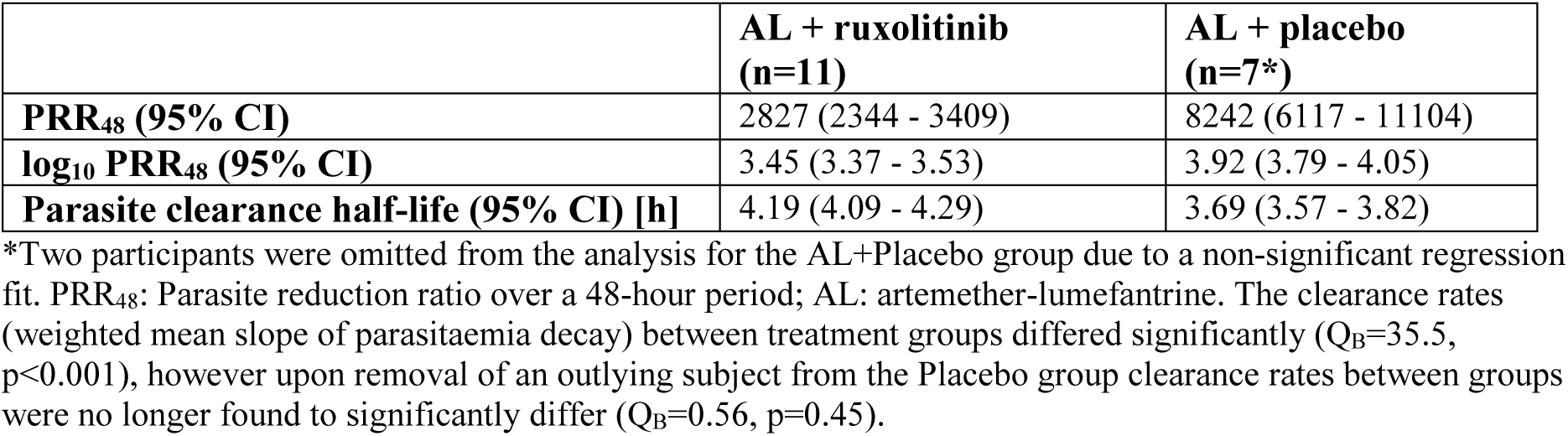
Parasite clearance parameters by treatment group.

|  | **AL + ruxolitinib**<br>**(n=11)** | **AL + placebo**<br>**(n=7\*)** |
| --- | --- | --- |
| **PRR<sub>48</sub> (95% CI)** | 2827 (2344 - 3409) | 8242 (6117 - 11104) |
| **log<sub>10</sub> PRR<sub>48</sub> (95% CI)** | 3.45 (3.37 - 3.53) | 3.92 (3.79 - 4.05) |
| **Parasite clearance half-life (95% CI) [h]** | 4.19 (4.09 - 4.29) | 3.69 (3.57 - 3.82) |

\*Two participants were omitted from the analysis for the AL+Placebo group due to a non-significant regression fit. PRR<sub>48</sub>: Parasite reduction ratio over a 48-hour period; AL: artemether-lumefantrine. The clearance rates (weighted mean slope of parasitaemia decay) between treatment groups differed significantly ( $Q_B=35.5$ , $p<0.001$ ), however upon removal of an outlying subject from the Placebo group clearance rates between groups were no longer found to significantly differ ( $Q_B=0.56$ , $p=0.45$ ).

### Impact of ruxolitinib on STAT phosphorylation

To assess the impact of ruxolitinib on cell signalling, STAT1, STAT3 and STAT5 phosphorylation was measured following IL-6 stimulation of whole blood. Samples were collected immediately prior to inoculation, immediately prior to treatment, 1 day after treatment (2 hours post ruxolitinib/placebo dose 3), and 5 days after treatment. Whole blood was stimulated with the JAK/STAT activating cytokine IL-6 or left unstimulated for 15 minutes and STAT phosphorylation (pSTAT) assessed in myeloid cells and CD4^+^ T cells using cytometry by time-of-flight (CyTOF) (**Figure 2A, Table S9**). Following batch correction, myeloid cells and CD4^+^ T cells were identified based on expression of cell lineage markers (**Figure 2B**).

**Figure 2:**
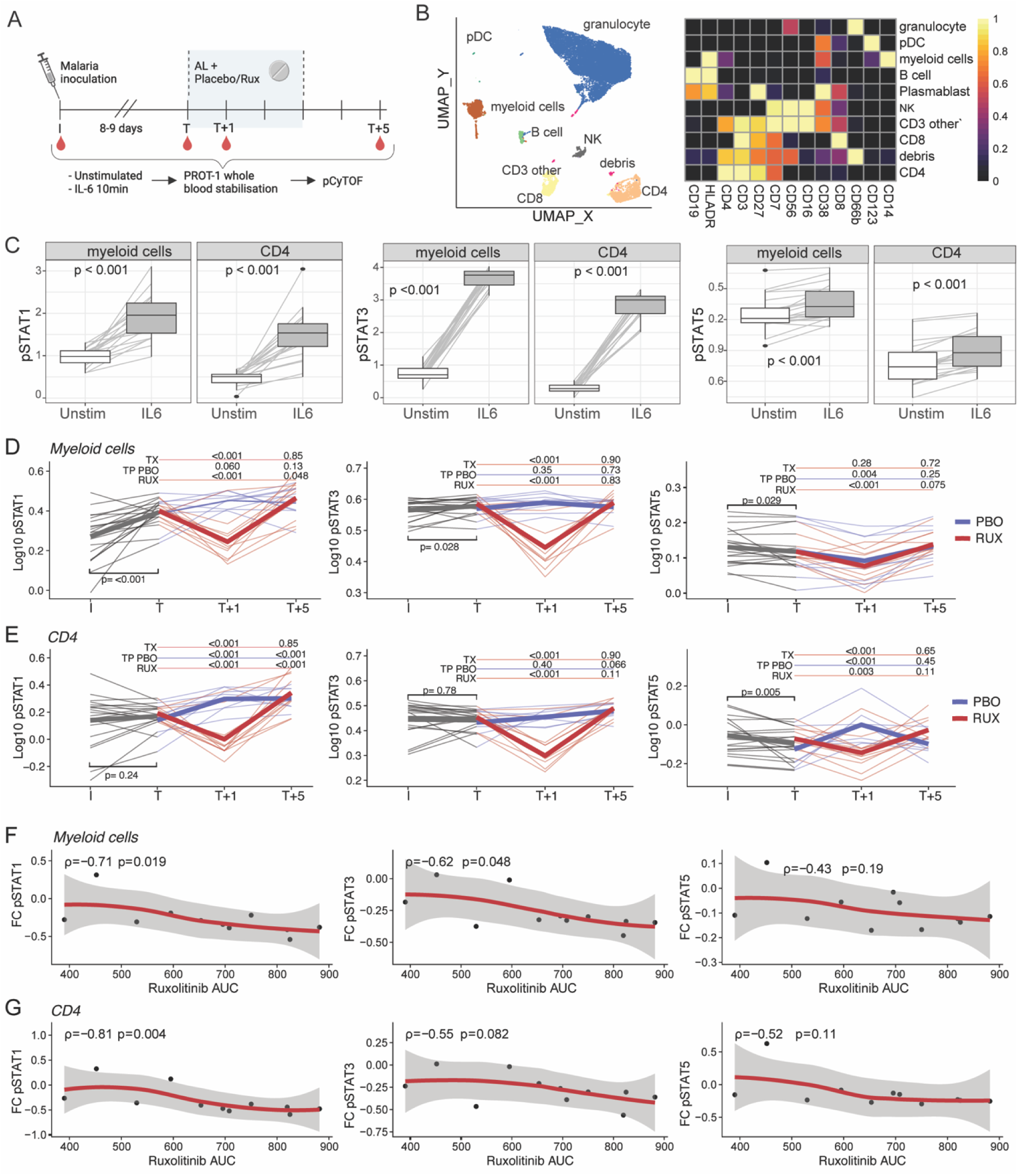
Impact of ruxolitinib treatment on cell signalling. **A)** Whole blood was collected at inoculation (I), treatment (T), treatment + 1 (2 hours after 3rd dose of ruxolitinib/placebo) and treatment +5 days. Blood was stimulated with IL-6 or left unstimulated, fixed in PROT1 and analysed by phosph-CyTOF. **B)** Cell subsets were identified by expression profiles of linage markers. **C)** pSTAT1, pSTAT3 and pSTAT5 expression as median signalling intensity values in myeloid cells and CD4 T cells in unstimulated and IL-6 stimulated samples at inoculation. P is Wilcoxon signed-rank test. **D/E)** pSTAT1, pSTAT3, pSTAT5 expression in myeloid cells **(D)** and CD4 T cells **(E)** at I, T, T+1, T+5. Data are log transformed median signalling intensity, with thin lines representing individuals and coloured by treatment group. Prior to randomisation is in grey, following randomisation ruxolitinib individuals are red and placebo are blue. P values are from linear mixed effect models, with bold lines representing the mean of the predicted values from the fitted models for each group. TX is FDR corrected P values for the interaction term between each timepoint (compared to T) and treatment groups. TP is FDR corrected P values for the comparison between each timepoint at T for either placebo (PBO) or ruxolitinib (RUX) groups **(F/G).** Fold Change (FC) of MFI of pSTAT1/3/5 at T compared to T+1 in ruxolitinib treated group in myeloid **(F)** or CD4 T cells **(G)**, correlated with ruxolitinib area under the curve (AUC). Spearmans rho and p are indicated.

IL-6 stimulation increased phosphorylation of STAT1, STAT3 and STAT5 (pSTAT) in both myeloid and CD4^+^ T cells (**Figure 2C**). Prior to randomisation, IL-6-induced expression of pSTAT1 and pSTAT3 on myeloid cells was significantly increased on the day of treatment compared to baseline (**Figure 2D**), indicating that infection with *P. falciparum* increased sensitivity of myeloid cells to IL-6. There was no increase in expression of pSTAT1/3 on CD4^+^ T cells, and levels of IL-6-induced pSTAT5 expression decreased on myeloid and CD4^+^ T cells (**Figure 2D/E**). Following randomisation and treatment, there was a marked decrease in IL-6-induced expression of pSTAT1 and pSTAT3 in myeloid and CD4^+^ T cells in the ruxolitinib group, but not in the placebo group (**Figure 2D/E)**. pSTAT5 was impacted by ruxolotinib treatment in CD4^+^ T cells but not myeloid cells. The fold-change reduction of IL-6-induced pSTAT1 and pSTAT3 expression on myeloid cells and CD4^+^ T cells was correlated with ruxolitinib AUC (**Figure 2F/G**).

### Impact of ruxolitinib on haematological and biochemical parameters

As expected in malaria volunteer infection studies, reductions in haemoglobin, platelets and lymphocytes were observed in both treatment groups following inoculation (**Figure 3A-C**). Haemoglobin reached a nadir on day 7 following treatment, with no difference between treatment groups. In the placebo group, reductions following treatment were also observed for platelets and lymphocytes, reaching nadirs on days 3 and 1, respectively. These post-treatment reductions were not observed in the ruxolitinib group, with a significant interaction term between treatment and time on day 3 for platelets (p=0.022) and day 1 for lymphocytes (p<0.001). Neutrophils declined following treatment in both groups, reaching a nadir on day 3, but with no difference between treatment groups (**Figure 3D)**.

**Figure 3:**
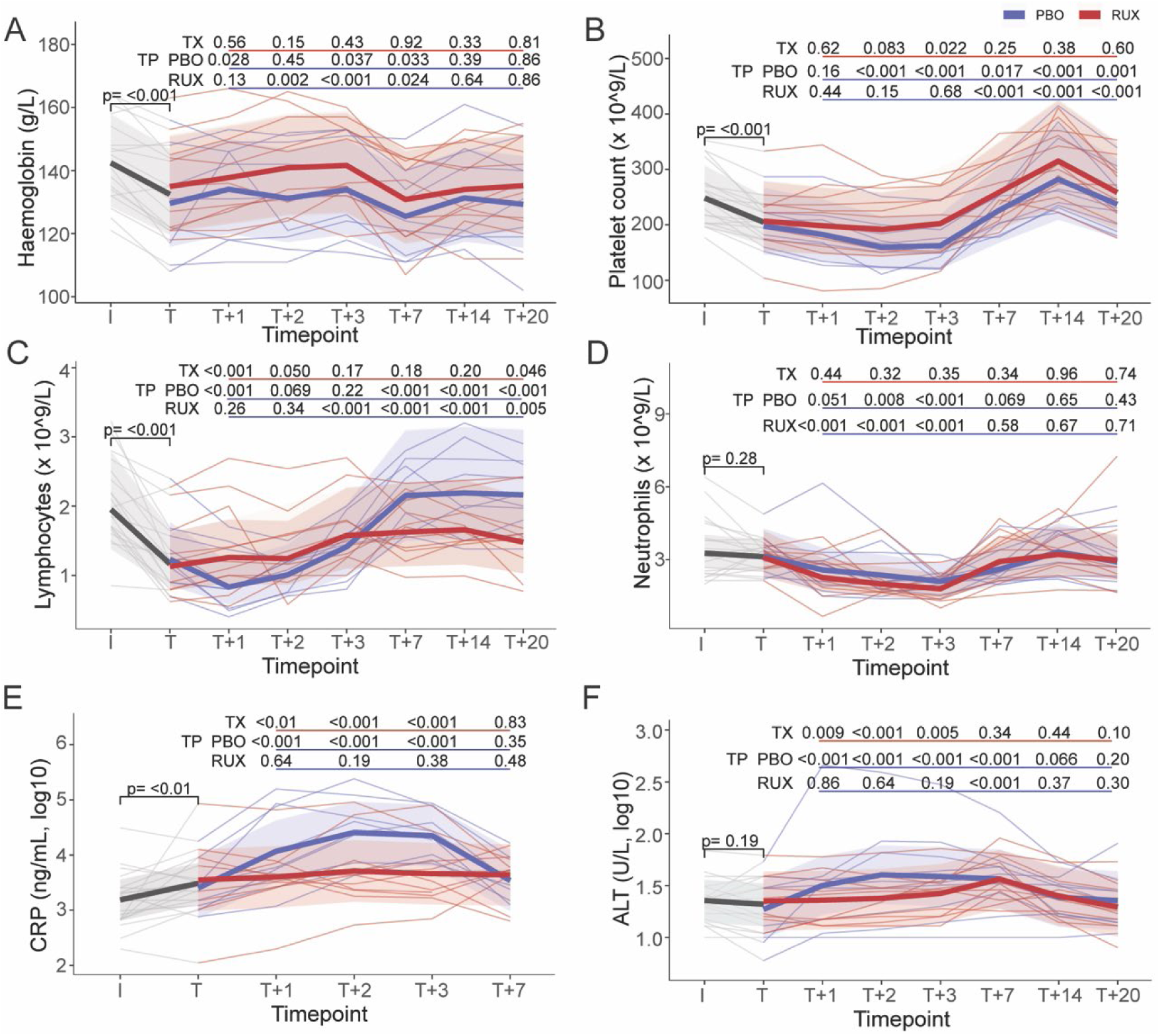
Impact of ruxolitinib on haematological and biochemical parameters. Haematological and biochemical parameters **A)** haemoglobin, **B)** platelet count, **C)** lymphocytes, **D)** neutrophils, **E)** CRP, and **F)** ALT at inoculation (I), treatment (T), and 3, 7 and 21 days after treatment (T+3, T+7, T+21). Data are raw data (log transformed for CRP and ALT) with thin lines representing individuals and coloured by treatment group, and bold lines representing the mean of the predicted values from the fitted models for each group. P values are FDR corrected from linear mixed effect models. TX is P values FDR corrected for the interaction term between each timepoint (compared to T) and treatment groups. TP is P FDR corrected for the comparison between each timepoint at T for either placebo (PBO) or ruxolitinib (RUX) groups.

The inflammatory marker C-reactive protein (CRP, measured by ELISA) increased in both groups following inoculation. Following treatment, CRP increased further in the placebo group, but not in the ruxolitinib group, with a significant impact of ruxolitinib treatment observed on days 1 to 3 post-treatment (**Figure 3E**). Notably, the peak CRP during the post-treatment period was strongly correlated with the pre-treatment parasitemia in the placebo group (r=0.80, p=0.013), but not in the ruxolitinib group (r=-0.04, p=0.91). The peak CRP was also associated with the total malaria clinical score during the confinement period (r=0.64, p=0.003) (**Table S3**).

As previously observed in malaria volunteer infection studies, ALT also increased modestly following treatment; this increase was less marked in those in the ruxolitinib group, with a significant impact of treatment observed on days 1 to 3 post-treatment (**Figure 3F**).

### Impact of ruxolitinib on systemic inflammation

The impact of ruxolitinib on the inflammatory response following treatment was further assessed by Alamar NULISAseq (250 analytes) on plasma collected prior to inoculation, prior to treatment, and days 3, 7 and 21 following treatment. First, inflammatory markers known to be associated with disease severity in *P. falciparum* malaria, including IL-6 [30], the endothelial activation marker angiopoietin-2 [31], ICAM-1 [32], and syndecan-1 (a marker of breakdown of endothelial glycocalyx) [33], were analysed. Of these, IL-6, angiopoietin-2 and ICAM-1 increased significantly from pre-inoculation to day of treatment (**Figure 4A-D**). Following treatment, angiopoietin-2, ICAM-1 and syndecan-1 all increased significantly in the placebo group, with ICAM-1 peaking at day 3, and angiopoietin-2 and syndecan-1 peaking at day 7 (**Figure 4A-D**). These post-treatment increases were not observed, or significantly reduced, in the ruxolitinib group, with a significant impact of treatment detected on day 3 post-treatment for ICAM-1, days 3 and 7 for angiopoietin-2, and day 7 for syndecan-1.

**Figure 4:**
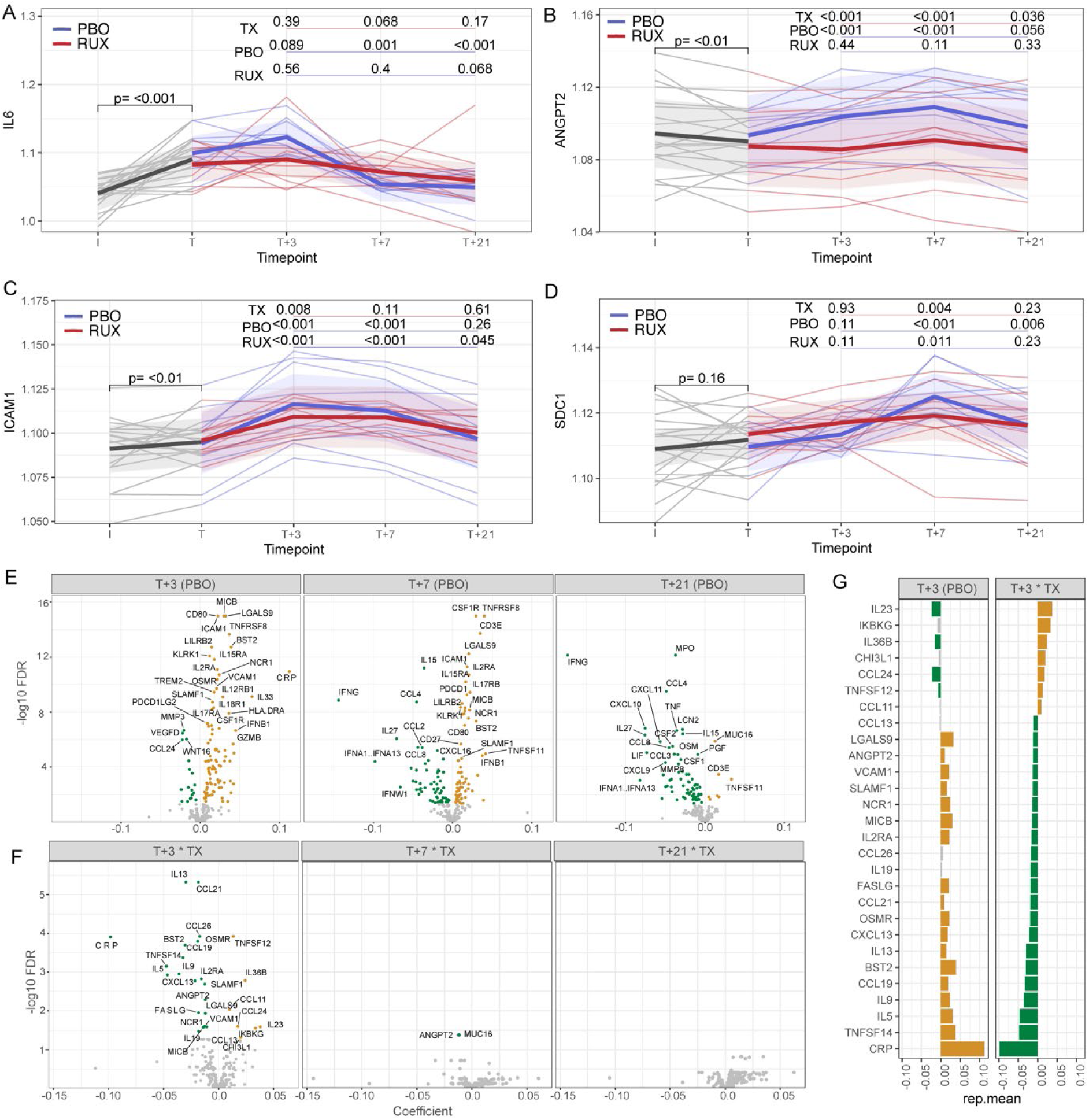
Impact of ruxolitinib on systemic inflammatory response. Alamar NULISAseq was used to quantify 250 analytes in plasma at inoculation (I), treatment (T), and 3, 7 and 21 days after treatment (T+3, T+7, T+21). **A)** IL-6, **B)** angiopoietin-2 (ANGPT2), **C)** ICAM1 and **D)** syndecan-1 (SDC1) over the course of trial. Data are log-transformed with thin lines representing individuals and coloured by treatment group (grey prior to inoculation, red in ruxoltinib treated and blue in placebo treated groups), and bold lines representing the mean of the predicted values from the fitted models for each group. P values (unadjusted) are from linear mixed effect models. TX is P values (un-adjusted) for the interaction term between each timepoint (compared to T) and treatment groups. TP is P for the comparison between each timepoint at T for either placebo (PBO) or ruxolitinib (RUX) groups. **E)** Volcano plots of coefficient and FDR (log10) of change of analyte at each timepoint compared to T in the placebo group. Coefficient and FDR values are from linear mixed effect model analysis. Analytes that are upregulated are in gold, and downregulated in green **F)** Volcano plots of coefficient (rep.mean) and FDR (log_10_) of interaction term between ruxolitinib treatment group and change of analyte at each timepoint compared to T. Coefficient and FDR values are from linear mixed effect model analysis. Analytes that are upregulated compared to the change in placebo are in gold, and those that are downregulated compared to the change in placebo are in green. **G)** Individual analytes at T+3 that were modified by ruxolitinib. Data are the coefficient for the change compared to T in the placebo group (T+3 (PBO)) and the coefficient for the interaction term between time and ruxolitinib treatment (T+3*TX)

To further examine the impact of ruxolitinib on the host inflammatory response, all analytes were analysed. As expected, *P. falciparum* infection significantly modulated the host response in both groups, with 139 (55.6%) analytes increased and 7 (2.8%) decreased between inoculation and treatment (false discovery rate [FDR] corrected, **Figure S2A**). IFNγ was the most upregulated analyte, with other inflammatory markers including TNF, CXCL9/10/11, CCL4, IL6, and CRP also strongly upregulated (**Figure S2A/B)**. Pathway analysis of upregulated analytes identified an inflammatory response, with TNF signalling via NK-kB and IL-2/STAT4 signalling, and IL-6/JAK/STAT3 signalling amongst the top upregulated pathways (**Figure S2C**).

Following treatment, 96 analytes increased and 18 decreased at day 3 in the placebo group (compared to the treatment timepoint) (**Figure 4E)**. Of these 114 analytes that changed significantly following treatment in the placebo group, a significant impact of ruxolitinib was detected for 28 (24.5%) analytes, 21 (18.4%) of which were down regulated by ruxolitinib, and 7 (6.1%) of which were increased (**Figure 4F)**. Analytes down regulated by ruxolitinib included CRP (confirming above analysis of the ELISA data), and TNFSF14 (LIGHT), the blockade of which we have previously shown protects against cerebral malaria in mice [34] (**Figure 4G)**. The modulation of ruxolitinib on angiopoietin-2 at both day 3 and day 7 post-treatment remained significant after FDR correction for all 250 analytes (p=0.005 and 0.042 respectively). At day 7 following treatment, 70 analytes remained elevated in the placebo group, but an impact of ruxolitinib was only detected for 2 analytes (angiopoietin-2 and MUC16) (**Figure 4F).** Pathway analysis of down regulated analytes identified down regulation of IL-6/JAK/STAT3 signalling and the inflammatory response (**Figure S2D).** Thus, the data indicated that ruxolitinib had a major modulatory impact on post treatment inflammation.

### Parasite growth and inflammatory responses following the second inoculation

To assess the impact of ruxolitinib on anti-parasitic immune responses to a subsequent infection, participants were re-inoculated with the same inoculum of *P. falciparum* 3D7 at 90 days after the first inoculation. No evidence of a difference in time to parasitaemia (qPCR positive) was found between treatment groups following second malaria inoculation (Fisher’s exact, p-value=0.29). The parasite growth rate was 0.08 log_10_ parasites/ml per day lower (95% CI 0.02 – 0.14) following the second inoculation compared to the first (p=0.009) (**Figure 5A, Table S10).** However, there was no evidence of a difference in the change in parasite growth rate between infections between ruxolitinib and placebo groups (log-likelihood ratio test - χ²(2)=4.24, p=0.12)(**Table 5, Table S10**).

**Figure 5.**
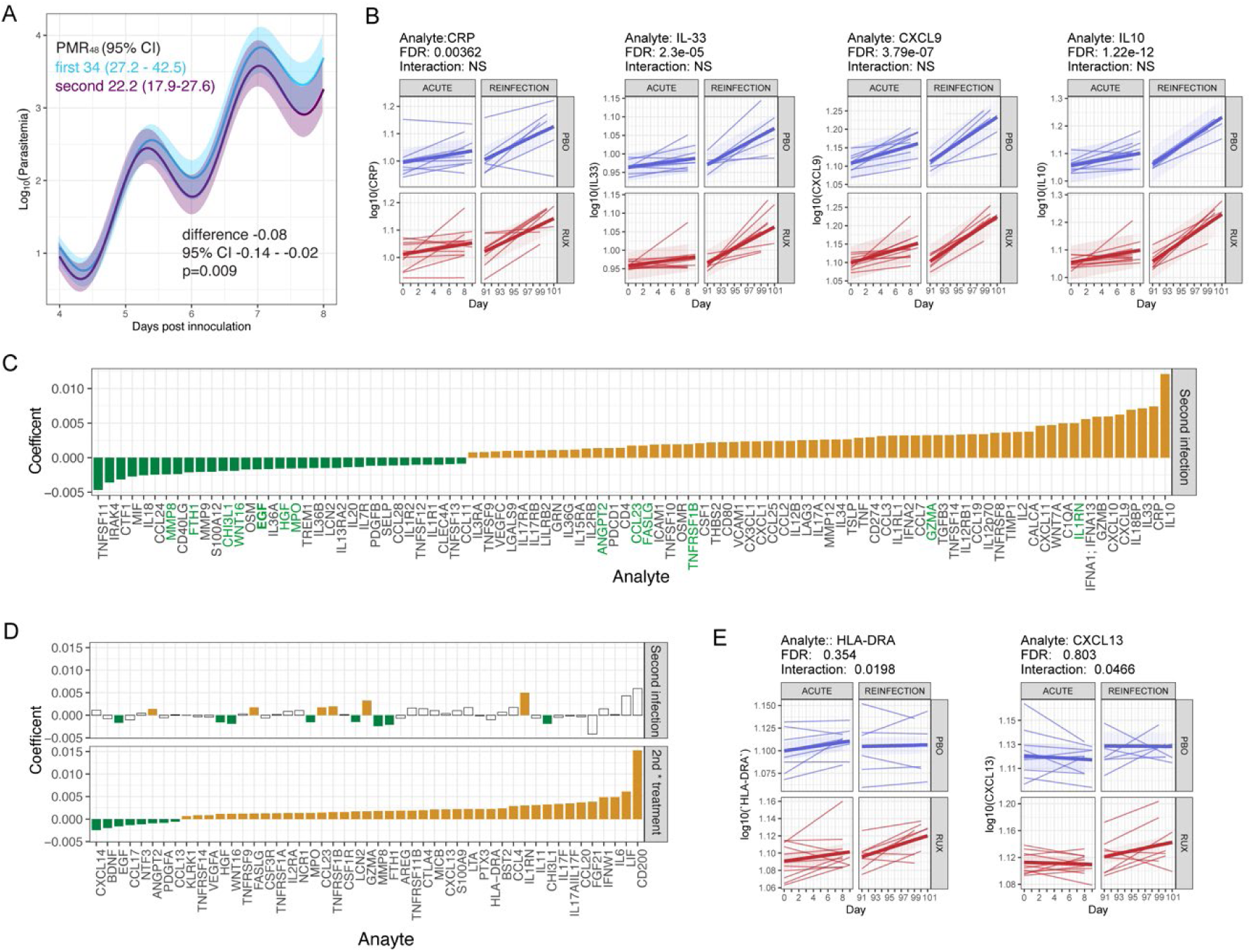
Immune response to second infection. **A)** Parasite multiplication rate (PMR) in first and second infection in all participants. The difference between first and second infection calculated from mixed effect segmented regression sine wave model. **B)** Change in CRP, IL-33, CXCL9 and IL-10 between inoculation and treatment in first and second infection in placebo (PBO) and ruxolitinib treated (RUX) groups. FDR is for a significant difference in the slopes for each analyte between infections. Interaction (all non-significant NS) is for the interaction term for an impact of treatment on the change in induction between infections. **C)** Coefficient for the change in slope between first and second infection for each analyte with an FDR for a difference of <0.05. The change in response of analytes in green was modified by ruxolitinib. **D)** Coefficient for the change in slope between first and second infection and for the interaction term of impact of treatment group for all analytes were FDR <0.05. Change for second infection is in top panel, impact of treatment is in bottom panel (2^nd^ * treatment). Non-significant changes in second infection only are in white. **E)** HLA-DR and CXCL13 responses in first and second infection in placebo and ruxolitinib treated individuals.

**Table 5.** Parasite multiplication rate (PMR_48_) estimates by inoculation phase and treatment group.

| Parameter | PMR<sub>48</sub> (95% CI) |
| --- | --- |
| Inoculation 1 | 34.0 (27.2 – 42.5) |
| Inoculation 2 |  |
| Placebo | 21.6 (16.4 – 31.2) |
| Ruxolitinib | 22.4 (16.0 – 31.4) |

To assess the memory immune response to second infection, NULISAseq data were analysed to assess the changes between inoculation and treatment in first compared to second infection. Consistent with an induced memory response and increased parasite control in second infection, multiple inflammatory responses were increased in second infection relative to first infection including CRP, IL-33, CXCL9, and IL-10 (**Figure 5B**). Overall, 95 analytes (38%) were modified in second infection, with 62 upregulated and 33 down regulated in comparison to first infection (**Figure 5C**). In addition, for 49 analytes (19.6%), the change in response in second infection was significantly modified by ruxolitinib treatment (**Figure 5D**). This included increased induction of HLA-DR, consistent with increased immune activation, and CXCL13, a plasma marker of germinal centre responses [35], in ruxolitinib treated individuals. Together data support the induction of a memory response to malaria following re-inoculation, which was enhanced in ruxolitinib treated individuals.

## Discussion

In this randomised placebo-controlled trial in volunteers infected with *P. falciparum*, host-directed adjunctive therapy with ruxolitinib was safe and well-tolerated, transiently reduced cellular pSTAT signalling, and attenuated the host inflammatory response following antimalarial treatment. Additionally, analysis of plasma milieu data following a second inoculation indicated that an immune memory response was induced by the initial *P. falciparum* infection, and was modulated in participants who had received ruxolitinib treatment. These findings highlight the potential of adjunctive ruxolitinib to improve clinical outcomes and enhance immune development in malaria.

Our key hypothesis underlying this study was that ruxolitinib, a JAK 1/2 inhibitor, would block type-I IFNs which are upregulated in malaria, thereby inhibiting the malaria-induced immunoregulatory networks that impair the development of antiparasitic immunity following either natural infection or vaccination [21]. We first confirmed that ruxolitinib effectively inhibited JAK/STAT signalling by measuring phosphorylation of STATs following stimulation with the JAK/STAT activating cytokine IL-6. Administration of ruxolitinib led to a marked decrease in IL-6 induced pSTAT1 and pSTAT3 expression in myeloid and CD4+ T cells, and pSTAT5 expression in CD4+ T cells, with these reductions significantly associated with ruxolitinib plasma concentration. These data therefore confirmed the ability of ruxolitinib to inhibit JAK/STAT signalling, consistent with an inhibitory effect of ruxolitinib on type-1 IFNs.

A key finding in this study was the impact of ruxolitinib on the post-treatment inflammatory response, evaluated with standard haematological/biochemical parameters as well as with the highly sensitive NULISA platform. As expected, inflammatory markers were increased in participants following inoculation with *P. falciparum*. Notably, in the placebo group, this response increased further following administration of antimalarial treatment, with the key inflammatory marker CRP peaking 2 - 3 days following treatment. Other key markers of disease severity also increased following treatment, including angiopoietin-2 and ICAM-1, markers of endothelial activation [36] and cytoadherence [37], respectively. Both markers are associated with disease severity in children and adults with severe falciparum malaria [31, 32, 38, 39]. This post-treatment increase in these inflammatory markers likely reflects the host response to parasite death and rupture of infected erythrocytes, and likely contributes to the high mortality from severe malaria even after commencing highly effective antiparasitic treatment [2]. While this post-treatment inflammatory response therefore represents a key target for adjunctive therapies aimed at improving clinical outcomes in severe malaria, to-date, no adjunctive treatment has been shown to reduce mortality [40]. Here we found that ruxolitinib substantially attenuated post-treatment increases in CRP, angiopoietin-2 and ICAM-1. Notably, peak CRP levels in placebo-treated participants correlated strongly with pre-treatment parasitaemia, whereas this correlation was absent in the ruxolitinib group, further supporting the role of ruxolitinib in inhibiting the parasitaemia-induced inflammatory process. These data are consistent with the potent anti-inflammatory effects of ruxolitinib in *in vitro* models [41], and the reduced levels of inflammatory biomarkers observed when ruxolitinib is used in myeloproliferative disorders [42]. Together, these data suggest a potential role for adjunctive ruxolitinib for improving clinical outcomes in severe malaria. Of note, another immunomodulator, imatinib, has recently been found to be safe in patients with uncomplicated malaria [43], providing additional confidence for proceeding with this treatment strategy.

Our data provide reassurance of the safety of ruxolitinib in patients with malaria. There was no evidence of an increased number of adverse events in participants who received ruxolitinib, nor any increase in the malaria clinical score during the post-treatment confinement period. Furthermore, we found that the malaria clinical score was correlated with CRP, suggesting that by reducing the inflammatory response, ruxolitinib would potentially reduce the severity of symptoms in clinical malaria. Although these data provide reassurance about the safety to ruxolitinib, we did find that ruxolitinib reduced plasma concentrations of artemether and its active metabolite DHA. Additionally, parasite clearance was slower in participants in the ruxolitinib group compared to the placebo group. Although this latter finding appeared partly driven by a single participant in the placebo group with particularly rapid parasite clearance (with no significant difference in parasite clearance observed once this outlier was excluded), parasite clearance parameters will need to be carefully monitored in future trials evaluating adjunctive ruxolitinib therapy.

To assess how ruxolitinib affects the development of long-lasting anti-parasitic immunity, participants who remained available underwent a second *P. falciparum* infection three months after the initial challenge. After re-infection, both treatment groups showed increased levels of immune molecules linked to parasite control. These included chemokines like CCL19 and CCL21, which help guide CD4+ T cells to secondary lymphoid tissues [44]. Inflammatory markers such as CRP, granzyme B (GZMB), and granzyme A (GZMA) were also elevated, along with immunoregulatory molecules like IL-18BP, LAG3, and IL-10. However, ruxolitinib treatment during the first infection appeared to improve the shaping of memory and recall immune responses. Notably, levels of CXCL13, a chemokine crucial for germinal centre B cell organisation and a known biomarker of germinal centre activity [45, 46], were higher in ruxolitinib-treated participants compared to controls during reinfection. Additionally, ruxolitinib increased the expression of molecules linked to T cell activation tissue repair, such as amphiregulin [47], IL-11 [48], and colony-stimulating factor 1 receptor (CSF1R) [49]. These findings suggest that ruxolitinib during primary infection leaves a lasting immunological imprint, influencing immune responses upon re-infection. While this phenomenon is well known in viral infections [50], its relevance to malaria is gaining recognition [21]. Further studies are needed to identify the specific immune cell populations affected by ruxolitinib and to fully understand its impact on host immunity.

Our study had limitations. First, due to difficulties with recruitment, only 20 of a planned 26 participants were enrolled, with only 15 participants proceeding to a second inoculation. This limited our power to detect between-group difference, such as in parasite growth following the second inoculation. Nonetheless, significant differences in inflammatory and immune parameters were observed between treatment groups. Second, for the analysis of between-group differences in parasite clearance, 2 participants in the placebo group had non-significant regression models and were unable to be included in the analysis. Together with the fact that another participant in the placebo group appeared to be an outlier with particularly rapid parasite clearance, uncertainty exists around our finding that parasite clearance may have been delayed by ruxolitinib, and further data are required. Third, plasma samples for analysis of inflammatory parameters by NULISAseq were collected only on days 3, 7 and 21 following treatment of the first infection, and impacts of adjunctive ruxolitinib treatment during the first 24 – 48 hours following treatment may have been missed. Finally, our study was conducted in malaria-naïve volunteers in Australia, and results may not reflect the effects of ruxolitinib in malaria-exposed individuals who represent the target population. Studies in these populations will be critical for better understanding the potential utility of adjunctive ruxolitinib.

In conclusion, in the randomised placebo-controlled trial in volunteers infected with *P. falciparum*, adjunctive treatment with ruxolitinib was safe and well tolerated, attenuated the post-treatment inflammatory response, and enhanced the immune memory response to a subsequent infection. While the potential impact of ruxolitinib on parasite clearance will require close monitoring, this study provides the basis for evaluating ruxolitinib as an adjunctive treatment to improve clinical outcomes and boost anti-parasitic immunity in clinical malaria.

## Methods

### Study design and participants

This was a randomised, double-blind, placebo-controlled, phase 1b clinical trial using the *P. falciparum* induced blood stage malaria (IBSM) model. Healthy malaria naïve males and females (non-pregnant, non-lactating) aged 18-55 years were eligible for inclusion (full eligibility criteria are in the Clinical Trial Protocol [Supplementary Material]). The study was conducted at the University of the Sunshine Coast Clinical Trails Unit (Morayfield, Australia) and was registered on the Australian New Zealand Clinical Trials Registry on 06 July 2021 (ACTRN12621000866808). This study was approved by the QIMR Berghofer Medical Research Institute Human Research Ethics Committee (P3696) and ethically reviewed in minimising duplication of ethics by the Australian Departments of Defence and Veterans’ Affairs Human Research Ethics Committee. All participants gave written informed consent.

### Procedures

Participants were inoculated intravenously with ∼2,800 viable *P. falciparum*-3D7 infected human erythrocytes on day 0. On day 9 (cohorts 1-2), or day 8 (cohorts 3-6), participants were admitted to the clinical unit and randomised in a 1:1 ratio to receive oral twice daily doses of artemether-lumefantrine (Riamet, Novartis Pharmaceuticals) in conjunction with either ruxolitinib (Jakavi, Novartis Pharmaceuticals) or placebo (microcrystalline cellulose, PCI Pharma). Artemether-lumefantrine dosing was in accordance with manufacturer recommendations for treatment of malaria (4 tablets [each containing 20 mg artemether and 120 mg lumefantrine] twice daily over 3 consecutive days with food or drink rich in fat). One ruxolitinib tablet (20 mg) or one placebo tablet was administered with 250 mL water 2 hours after each AL dose (total course of 6 tablets). A randomisation list was generated electronically using STATA 15 and was available only to authorised persons. Participants and investigators were blinded to treatment allocation (dosing was performed by an unblinded staff member and participants were blindfolded since ruxolitinib and placebo tablets were not matched in terms of appearance). Participants were confined to the clinical unit for 72 hours for drug dosing, blood sampling and clinical assessment, and were subsequently followed-up as outpatients to day 28±2.

A second (homologous) intravenous inoculation with ∼2,800 viable *P. falciparum*-infected human erythrocytes was administered on Day 90±7 to all participants who remained eligible (a repeat eligibility screening was performed on day 85±7). Participants were administered a standard course of artemether-lumefantrine (as described above) on an individualised basis when any of the following criteria were met: parasitaemia ≥50,000 parasites/mL, malaria clinical score >6 and presence of parasitaemia, SAE related to malaria challenge, grade 3 AE related to malaria challenge and not self-resolved or relieved with concomitant medications, investigator discretion based on participant safety, or 28 days after second inoculation if the preceding criteria were not met. The end of study visit occurred approximately 28 days after AL treatment initiation.

### Assessments

Quantification of *P. falciparum* parasitaemia was determined in venous blood samples by qPCR targeting the gene encoding 18S rRNA as described previously [51]. Plasma drug concentrations were determined via liquid chromatography tandem mass spectrometry (LC MS/MS) using validated methods. Rux was measured as described previously [28], with the internal standard Ruxolitinib-d9. The calibration range for Rux was 1.0 to 1,000 ng/mL using 50 μL of plasma. The analytical method was precise and accurate. The descriptive statistics of the quality control (QC) samples for Rux showed that the inter assay precision (% Coefficient of Variation) ranged from 5.3% and 8.2% (n=14) whereas the inter accuracy ranged from 100.1% to 102.9% (n=14) of the nominal concentrations. The QC concentrations were: 3 ng/mL for low QC, 20 ng/mL for mid QC, and 750 ng/mL for high QC for Rux.

Artemether and lumefantrine, and their active metabolites (dihydroartemisinin [DHA] and desbutyl-lumefantrine [DBL]), were measured as previously described [52]. The calibration ranges were 1.0 to 1,024 ng/mL using 50 μL of plasma. The QC concentrations were 4 ng/mL for low QC, 32 ng/mL for mid QC, and 512 ng/mL for high QC. The inter assay precision ranged from 3.5% to 5.9% (n=8) for artemether, 4.7% to 7.7% (n=8) for DHA, 4.6% to 8.5% (n=12) for lumefantrine and 5.7% to 7.3% (n=12) for DBL. The inter accuracy ranged from 94.9% to 103.7% (n=8) for artemether, 99.5% to 105.6% (n=8) for DHA, 94.3% to 101.5% (n=12) for lumefantrine and 98.7% to 101.8% (n=12) for DBL. For quality assurance, the analytical laboratory at the Australian Defence Force Malaria and Infectious Disease Institute participates in the WWARN proficiency testing/QC program for the measurement of plasma concentrations of artemether, DHA, lumefantrine and DBL [53].

Safety and tolerability were assessed by monitoring AEs, vital signs, clinical laboratory parameters (haematology, biochemistry and urinalysis), physical examination, and electrocardiographs (ECGs). A malaria clinical score was generated by grading 14 signs/symptoms frequently associated with malaria using a 4-point scale (absent 0; mild: 1; moderate: 2; severe: 3) and summing to generate a total score (maximum score possible is 42). The timing of all study assessments is indicated in the clinical trial protocol (Supplementary Material).

The inflammatory marker C-reactive protein (CRP) was measured using commercially available Human C-Reactive Protein ELISA Kit (Thermo Fisher Scientific #KHA0031) on plasma diluted 1/30 000 collected from a K2 EDTA tube. Samples were tested in duplicate according to manufacturer’s instructions. Absorbance was read at 450 nm on Biotek PowerWaveTM XS2 Microplate Reader, and concentrations calculated from a 4-parameter logistic regression of standard curve.

*Phosphoflow whole blood CYTOF:* 200 μL of whole blood (from Li-heparin) was stimulated with IL-6 (0.1ug/ml, Biosciences cat# BMS341) for 15 min at room temperature or left unstimulated, and then fixed with Proteomic Stabilizer (Smart Tube, Inc., San Carlos, CA) and kept at -80C. CyTOF was by the Human Immune Monitoring Center at Stanford University. Fixed samples were thawed, samples were washed with Smart Tube Thaw-Lyse buffer twice, and with Cell Staining Buffer (CSB; Standard Biotools) once. The samples where then washed twice with perm buffer (Invitrogen 10× permabilization buffer #00-8333-56, diluted to 1× in PBS from Standard Biotools). The samples were then resuspended in 900 μL perm buffer and 10 μL of each Standard Biotools CellID 20plex Pd barcode was added before incubating at RT for 30min. Next, the samples were centrifuged, aspirated, and washed three times by resuspension in CSB. The pellets were then resuspended in 450 μL CSB and pooled into a single tube. After centrifugation and aspiration, the pooled sample was counted on a Bio-Rad TC20 cell counter. Surface cocktail was added at 1 test = 20 μL = 1M counted cells and stained at RT for 30min. This cocktail was combined from two -80°C frozen cocktails (HIMC surface, Boyle surface) to which fresh 143Nd-CD45RA (Standard Biotools) was added. Next, the sample was washed twice by resuspension with CSB. For fixation, 4% PFA in PBS was added and incubated for 10 min at RT before adding PBS to 10 mL, centrifugation, and aspiration. Then, 1 mL of -20°C cold Methanol was added and the sample placed at -80°C overnight. On Day 2, the sample was centrifuged and the methanol aspirated before resuspension in 10 mL PBS and the centrifugation and aspiration repeated. The sample was resuspended in 1 mL CSB, and counted by TC20. Intracellular cocktail was added: 1 test = 20 μL = 1M counted cells and stained at RT for 30min. This cocktail was combined from two -80°C frozen cocktails (HIMC intracellular, Boyle intracellular). After staining, the sample was washed twice with CSB, centrifugation, and aspiration. The pellet was then resuspended in 2% PFA/PBS containing 250 nM Ir intercalator (Standard Biotools) and incubated for 20 min at RT. Cells were washed once with CSB and twice with MilliQ water. On the same day as staining finished, cells were diluted to 0.75M /mL in MilliQ water containing 10× diluted EQ4 normalization beads (Standard Biotools) and acquired on CyTOF. Data analysis was performed using FlowJo v10 by gating on intact cells based on the iridium isotopes from the intercalator, then on singlets by Ir191 vs cell length followed by cell clustering analyses. No live-dead was used as the sample was fresh complete whole-blood. For clustering analyses, data was batch corrected using the CyCombine package [54], clustered using FlowSOM [55], and annotated by expression profiles of lineage markers.

#### NULISAseq Inflammation Panel 250

This assay was performed by the Human Immune Monitoring Center (HIMC) at Stanford University. Samples were run on the NULISA Inflammation panel (Alamar BioSciences, Fremont, California), a multiplexed proximity ligation assay targeting 250 inflammation-associated proteins. The assay was processed automatically in the ARGO HT system (Alamar BioSciences). 25 μL of each sample was loaded on the sample plate, along with three sample controls (SC), four negative controls (NC), and three Inter-plate controls (IPC). After completion of the automated run, next-generation sequencing (Illumina, Foster City, California) was performed on the pooled library. Data were generated using ACC (Alamar Command Center) and NAS (NULISA Analysis Software) via normalization to Internal controls (IC) and Inter-Plate controls (IPC). Then data were rescaled using log2 transformation. These steps produced an output in NULISA Protein Quantification (NPQ) units. The participant who tested positive for COVID-19 in second infection was excluded from analysis for re-inoculation time points.

### Outcomes

The primary outcome was the incidence, severity, and causality of AEs. Secondary outcomes were parasite growth parameters (time to parasitaemia and parasite multiplication rate [PMR]), parasite clearance kinetics (parasite reduction ratio [PRR] and clearance half-life), drug pharmacokinetic (PK) parameters, immune response parameters, and pSTAT3 inhibition.

### Statistical analysis

The planned sample size was 26 participants, randomised 1:1 to ruxolitinib and placebo. This was based on the secondary outcome of the anti-parasitic immune response (IFNγ and IL-10 levels). Sample size calculations utilised data from a study which measured cytokine production following *P. falciparum* parasite stimulation of PBMCs collected from participants in previous *P. falciparum* IBSM trials [19]. With 13 participants per group in the current study, a two-sided two sample t-test was estimated to have 80% power to detect a 23% difference in mean IFNγ levels 7 days post-treatment, and 8% difference in mean IL-10 levels at the same timepoint, with a type I error rate of 5%. This sample size was also considered appropriate to assess the primary outcome associated with safety and tolerability. Statistical analyses were performed using R statistical package (version 4.1.0 or higher) or Stata version 18.

Severity of AEs following first and second inoculations was compared between treatment groups separately using Fisher’s exact tests overall. Spearman’s correlation was used to evaluate the association between the malaria clinical score, peak parasitaemia and peak CRP.

Plasma artemether, DHA, lumefantrine, DBL and RUX concentration-time data were analysed by noncompartmental methods using PKanalix (version 2023R1). Differences in PK parameters between placebo and RUX groups were tested using an unpaired two-sample t-test or Wilcoxon test.

The parasite reduction ratio over a 48-hour period (PRR_48_) and parasite clearance half-life (PCt_1/2_) were estimated using the slope of the optimal fit for the log-linear relationship of the parasitaemia decay [29]. Differences between treatment groups in parasite clearance parameters after first inoculation were assessed using an omnibus test.

The PMR was calculated by applying a sine-wave growth model to the parasitaemia data [56]. Mixed-effects models were used to obtain parameter estimates overall following first inoculation and for each treatment group following second inoculation. Changes in PMR between inoculation phases were assessed by applying a mixed effect segmented regression sine wave model with a treatment group by time interaction following second inoculation. Full model specification is provided in **Supplementary Text**. Time to parasitaemia after second inoculation was compared between groups using a Fisher’s exact test.

The change in response over time from either the inoculation or the treatment time-point was assessed using log-linear mixed effects models with REML estimation, incorporating a random intercept at the subject level. Treatment, time-point, and their interaction were included as fixed effects to evaluate how the response differs between treatment groups and how the change in response over time differs between treatment groups. The change over time for the Rux group assessed using contrasts. Residual bootstrapping at 5000 iterations was applied to account for potential deviations from normality in the residuals, ensuring robust and reliable estimates.

## Supporting information

Clinical Trial Protocol

## Data Availability

All data produced in the present study are available upon reasonable request to the authors.
NUlisa and phospho CyTOF whole blood data is publicly available: DOI 10.5281/zenodo.15080321 and DOI 10.5281/zenodo.14872946

## Acknowledgements

We thank the volunteers who participated in the clinical trial; staff at the University of Sunshine Coast clinical trial site; staff at the Queensland Paediatric Infectious Diseases Laboratory (QPID) for conducting the malaria PCR assays; members of the Stanford Human Immune Monitoring Center for technical assistance; and Karin Van Breda for technical assistance in the analysis of drug concentrations.

## Data availability

NUlisa and phospho CyTOF whole blood data is publicly available: DOI 10.5281/zenodo.15080321and DOI 10.5281/zenodo.14872946

## Funding

This study was funded by the Australian National Health and Medical Research Council (NHMRC); Ideas Grant to BEB and MJB (GNT2002957). BEB and JCM are supported by NHMRC Investigator Grants (2016792 and 2016396 respectively); MJB is supported by a Snow Medical Fellowship (2022/SF167) and CSL Centenary Fellowship. The QIMR Berghofer Clinical Malaria group receives core funding from Medicines for Malaria Venture (MMV) to support the conduct of malaria volunteer infection studies.

Work in the Stanford Human Immune Monitoring Center was supported by the Gates Foundation [INV-008378]. The conclusions and opinions expressed in this work are those of the author(s) alone and shall not be attributed to the Foundation. Under the grant conditions of the Foundation, a Creative Commons Attribution 4.0 License has already been assigned to the Author Accepted Manuscript version that might arise from this submission. Please note works submitted as a preprint have not undergone a peer review process.

## Competing interests

All authors report no competing interests.

## DISCLAIMER

The views expressed in this article are those of the authors and do not necessarily reflect the official policy or position of the Australian Defence Force, Joint Health Command or any extant Australian Defence Force policy.

## Notes

### Competing Interest Statement

The authors have declared no competing interest.

### Clinical Trial

Australian New Zealand Clinical Trials Registry (ACTRN12621000866808).

### Author Declarations

This study was approved by the QIMR Berghofer Medical Research Institute Human Research Ethics Committee (P3696) and ethically reviewed in minimising duplication of ethics by the Australian Departments of Defence and Veterans Affairs Human Research Ethics Committee.

