## Supplementary Material for "Adjunctive ruxolitinib attenuates inflammation and enhances antiparasitic immunity in human volunteers experimentally infected with *Plasmodium falciparum*"

**Supplementary Text.**

**Analysis of PMR between study phases**

The impact of inoculation phase and treatment group were assessed using a mixed effects segmented regression sine wave model with REML estimation, using all subjects' growth data. Parasitaemia was measured from day 4 following each inoculation. For modelling purposes time following inoculation 2 was considered 'time from inoculation + 6 days' (i.e. day 4 converted to day 10). The segmented regression model can be described as follows for participant *i*, at time *j*:

$$\log_{10}(Y_{ij}) = a + m \times time_{ij} + c \times sin\left(\left(2 \times \frac{\pi}{period}\right) \times inoctime_{ij} + k\right) + p \times phase_{ij} + g \times group_{ij} + pt \times phase_{ij} \times [time_i - time_{p2}] + pgt \times phase_{ij} \times group_{ij} \times [time_i - time_{p2}]$$

Where:

- $Y$ = parasites per mL measured by qPCR at multiple times from inoculation to first anti-malarial treatment
- $a + c \times \sin(k)$ = intercept
- $m$ = parasite growth rate
- $c$ = amplitude of the sine wave
- $period$ = length of a parasite life-cycle in days
- $k$ = phase shift in sine wave.
- $inoctime_{ij}$= days from inoculation (first or second)
- $time_{ij}$= Days from first inoculation
- $time_{p2}$= Day of second inoculation (day 6 for modelling purposes)
- $phase_{ij}$= phase (Acute or Reinfection)
- $group_{ij}$= treatment group (Rux or placebo) in phase 2

Interactions:

- $phase \times [time_i - time_{p2}]$= Phase by time from second inoculation
- $phase \times group \times [time_i - time_{p2}]$= Phase by group by time from second inoculation

A subject-specific random effect for the intercept (*a* parameter) and phase parameter were included in the model and random slopes for each of the time main effects and time interaction terms assessed and included based on model AIC and random effect magnitude (m parameter, phase by time interaction). Models considered independent correlation structure only. Importance of the phase by group by time interaction term in was assessed using a log-likelihood ratio test compared to a treatment group main effects model.

The growth rate parameter from this model were transformed into the $PMR_{48}$ for reporting by: $PMR_{48} = 10^{(2m)}$, where *m* was the parasite growth rate per day estimated by the log-linear or sine-wave growth model, and 2 days is the accepted parasite life-cycle of 48 hours. The PMR can be considered a fold-change in the parasite density over a 48 hour life-cycle of *P. falciparum.* Analysis was run in Stata version 18.

### Supplementary Tables

**Table S1. Number of adverse events per participant by study phase**

| Study Phase | Rux | Placebo | Total |
|---|---|---|---|
| First inoculation (Day 0 to Day 89) | 6 (1 - 18) | 8 (1 - 25) | 7 (1 - 25) |
| pre-IMP dosing | 2 (1 - 6) | 3 (1 - 7) | 3 (1 - 7) |
| post-IMP dosing | 5 (1 - 16) | 7 (1 - 18) | 5.5 (1 - 18) |
| Second inoculation (Day 90 to Day 130) | 7 (1 - 18) | 7.5 (1 - 10) | 7 (1 - 18) |

*Values are median (range)

**Table S2. Malaria Clinical Score**

| Symptom or sign | Clinical Score/CTCAE grade | | | |
|---|---|---|---|---|
| | Absent (0) | Mild (1) | Moderate (2) | Severe (3) |
| | | CTCAE 1 | CTCAE 2 | CTCAE 3 |
| Headache | | Mild pain | Moderate pain; limiting instrumental activities of daily living (ADL) | Severe pain; limiting self-care ADL |
| Myalgia | | Mild pain | Moderate pain; limiting instrumental ADL | Severe pain; limiting self-care ADL |
| Arthralgia | | Mild pain | Moderate pain; limiting instrumental ADL | Severe pain; limiting self-care ADL |
| Fatigue | | Fatigue relieved by rest | Fatigue not relieved by rest; limiting instrumental ADL | Fatigue not relieved by rest; limiting self-care ADL |
| Malaise | | Uneasiness or lack of well-being | Uneasiness or lack of well-being; limiting instrumental ADL | Uneasiness or lack of well-being limiting self-care ADL |
| Chills | | Mild sensation of cold; shivering; chattering of teeth | Moderate tremor of the entire body; narcotics indicated | Severe or prolonged, not responsive to narcotics |
| Sweating/hot spells | | Mild sweating/hot spells not affecting ADL | Moderate sweating/hot spells; narcotics indicated | Severe or prolonged, not responsive to narcotics |
| Reduced appetite | | Loss of appetite without alteration in eating habits | Oral intake altered without significant weight loss or malnutrition; oral nutritional supplements indicated | Associated with significant weight loss or malnutrition (e.g. inadequate oral caloric and/or fluid intake); tube feeding or TPN indicated |
| Nausea | | Loss of appetite without alteration in eating habits | Oral intake decreased without significant weight loss, dehydration or malnutrition | Inadequate oral caloric or fluid intake; tube feeding, TPN, or hospitalisation indicated |
| Vomiting | | Intervention not indicated | Outpatient IV hydration; medical intervention indicated | Tube feeding, TPN, or hospitalisation indicated |
| Abdominal discomfort | | Mild pain | Moderate pain; limiting instrumental ADL | Severe pain; limiting self-care ADL |
| Fever | | 38.0-38.9°C | ≥39.0-39.9°C | ≥40.0°C |
| Tachycardia | | HR ≥100 Asymptomatic, intervention not indicated | HR ≥100 Symptomatic; non-urgent medical intervention indicated | HR ≥100 Urgent medical intervention indicated |

| Symptom or sign | Clinical Score/CTCAE grade | | | |
|---|---|---|---|---|
| Hypotension | | SBP ≤80 Asymptomatic, intervention not indicated | SBP ≤80 Symptomatic; non-urgent medical intervention indicated | SBP ≤80 Urgent medical intervention indicated |

The malaria clinical score gives an indication of the severity of the induced malaria infection in each volunteer. Fourteen signs/symptoms frequently associated with malaria are graded using a 4-point scale (absent 0; mild: 1; moderate: 2; severe: 3) and summed to generate a total malaria clinical score (maximum score possible is 42). Severity was graded in accordance with the CTCAE Version 5.0 Published: November 27, 2017.

**Table S3. Parasitemia, CRP, and Malaria Clinical Score following ruxolitinib/placebo dosing in the first inoculation phase**

| Participant number | Malaria clinical score | | | | | | | | | | Pre-treatment parasitemia (parasites/ml) | Peak CRP* (ng/mL) |
|---|---|---|---|---|---|---|---|---|---|---|---|---|
| | Hours post first dose | | | | | | | | | | | |
| | 6 | 12 | 24 | 30 | 36 | 48 | 54 | 60 | 72 | Total | | |
| **RUX** | | | | | | | | | | | | |
| **2** | 0 | 0 | 1 | 0 | 0 | 0 | 0 | 0 | 0 | 1 | 37,107 | 4,500 |
| **5** | 0 | 1 | 0 | 0 | 2 | 0 | 0 | 0 | 0 | 3 | 149,520 | 5,589 |
| **6** | 0 | 0 | 0 | 0 | 0 | 0 | 0 | 0 | 0 | 0 | 7,831 | 7,770 |
| **8** | 0 | 1 | 0 | 1 | 2 | 0 | 0 | 0 | 0 | 4 | 13,463 | 8,007 |
| **9** | 0 | 2 | 0 | 0 | 0 | 0 | 0 | 0 | 0 | 2 | 6,091 | 91,307 |
| **11** | 0 | 1 | 1 | 0 | 0 | 0 | 1 | 0 | 0 | 3 | 12,959 | 4,322 |
| **15** | 0 | 0 | 0 | 0 | 2 | 0 | 3 | 0 | 1 | 6 | 11,572 | 80,711 |
| **16** | 0 | 0 | 0 | 0 | 0 | 0 | 0 | 0 | 0 | 0 | 968 | 7,718 |
| **17** | 0 | 0 | 0 | 0 | 0 | 0 | 0 | 0 | 0 | 0 | 7,331 | 701 |
| **19** | 0 | 0 | 0 | 0 | 0 | 0 | 0 | 0 | 0 | 0 | 4,772 | 4,023 |
| **20** | 1 | 1 | 0 | 0 | 0 | 1 | 0 | 1 | 0 | 4 | 17,135 | 5,077 |
| **Placebo** | | | | | | | | | | | | |
| **1** | 1 | 1 | 1 | 2 | 1 | 1 | 0 | 0 | 0 | 7 | 121,691 | 157,814 |
| **3** | 3 | 5 | 2 | 0 | 2 | 1 | 0 | 0 | 0 | 13 | 251,801 | 85,716 |
| **4** | 0 | 2 | 0 | 1 | 2 | 0 | 0 | 2 | 0 | 7 | 52,677 | 40,470 |
| **7** | 1 | 0 | 0 | 0 | 0 | 0 | 0 | 0 | 0 | 1 | 33,566 | 10,187 |
| **10** | 0 | 0 | 0 | 1 | 0 | 0 | 0 | 0 | 0 | 1 | 1,028 | 22,987 |
| **12** | 5 | 1 | 3 | 4 | 7 | 7 | 4 | 5 | 2 | 38 | 33,955 | 240,994 |
| **13** | 0 | 0 | 0 | 0 | 0 | 0 | 0 | 0 | 0 | 0 | 362 | 7,247 |
| **14** | 1 | 2 | 0 | 0 | 5 | 0 | 0 | 0 | 0 | 8 | 3,952 | 38,721 |
| **18** | 0 | 0 | 0 | 0 | 0 | 0 | 0 | 0 | 0 | 0 | 1,387 | 8,166 |

*peak CRP measured during the 72 hours following ruxolitinib/placebo dosing

**Table S4. Overall summary of adverse events during the second inoculation phase (Day 90 to Day 130)**

| | **Rux (N=9)** | | **Placebo (N=6)** | | **Total (N=15)** | |
|---|---|---|---|---|---|---|
| | **n (%)** | **E** | **N (%)** | **E** | **n (%)** | **E** |
| Any AE | 9 (100.0%) | 67 | 6 (100.0%) | 36 | 15 (100.0%) | 103 |
| Mild | 9 (100.0%) | 41 | 5 (83.3%) | 20 | 14 (93.3%) | 61 |
| Moderate | 7 (77.8%) | 24 | 5 (83.3%) | 15 | 12 (80.0%) | 39 |
| Severe | 2 (22.2%) | 2 | 1 (16.7%) | 1 | 3 (20.0%) | 3 |
| Challenge agent related AE | 8 (88.9%) | 48 | 5 (83.3%) | 26 | 13 (86.7%) | 74 |
| Mild | 8 (88.9%) | 31 | 4 (66.7%) | 15 | 13 (86.7%) | 74 |
| Moderate | 6 (66.7%) | 15 | 4 (66.7%) | 10 | 12 (80.0%) | 46 |
| Severe | 2 (22.2%) | 2 | 1 (16.7%) | 1 | 10 (66.7%) | 25 |
| SAE | 0 (0.0%) | 0 | 0 (0.0%) | 0 | 3 (20.0%) | 3 |

n(%): number and percentage of participants experiencing an AE; E=:number of AEs

**Table S5: Pharmacokinetic parameters for ruxolitinib following multiple-dose administration.**

| Parameters[a] | Ruxolitinib (n = 11) |
|---|---|
| First dose | |
| $AUC_{0\text{-}last}$ (h·ng/mL) | 644.69 (1.30) |
| $AUC_{0\text{-}inf}$ (h·ng/mL) | 778.58 (1.40)[c] |
| Cmax (ng/mL) | 192.14 (1.38) |
| tmax (h)[b] | 0.97 (0.93 – 2.98) |
| t½ (h) | 2.40 (1.55)[d] |
| Λz (/h) | 0.29 (1.55)[d] |
| CL/F (L/h) | 28.08 (1.42)[e] |
| Vz/F (L) | 74.28 (1.81)[e] |
| Last dose | |
| $AUC_{0\text{-}last}$ (h·ng/mL) | 640.15 (1.53)[f] |
| $AUC_{0\text{-}inf}$ (h·ng/mL) | 682.77 (1.33)[g] |
| Cmax (ng/mL) | 81.44 (1.30)[f] |
| tmax (h)[b] | 4.00 (3.98 – 9.98)[f] |

AUC0-last = area under the curve from 0 to last measurable concentration, AUC0-inf = area under the curve from 0 to infinity, Cmax = maximum concentration, tmax = time at maximum concentration, t½ = elimination half-life, λz = first-order terminal elimination rate constant, CL/F = apparent total clearance, Vz/F = apparent total volume of distribution.

a Values in geometric mean (geometric standard deviation) unless otherwise stated.
b Values in median (range).
c Values estimated in 4 of 11 subjects.
d Values estimated in 5 of 11 subjects.
e Value estimated in 3 of 11 subjects.
f Values estimated in 9 subjects as 2 subjects did not complete the 3-day ruxolitinib course.
g Values estimated in 6 of 9 subjects.

**Table S6. Pharmacokinetic parameters for artemether and dihydroartemisinin by treatment group following multiple-dose administration**

| Parameters[a] | Artemether | | | Dihydroartemisinin | | |
|---|---|---|---|---|---|---|
| | **Placebo (n = 9)** | **Ruxolitinib (n = 11)** | ***p*-value** | **Placebo (n = 9)** | **Ruxolitinib (n = 11)** | ***p*-value** |
| First dose | | | | | | |
| $AUC_{0\text{-last}}$ (h·ng/mL) | 183.99 (1.76) | 111.76 (1.65) | 0.048 | 168.67 (1.56) | 135.19 (1.31) | 0.129 |
| $AUC_{0\text{-inf}}$ (h·ng/mL) | 193.23 (1.78) | 117.50 (1.67) | 0.048 | 176.68 (1.56) | 142.73 (1.31) | 0.129 |
| $C_{max}$ (ng/mL) | 67.80 (1.68) | 35.41 (1.77) | 0.028 | 59.93 (1.62) | 41.45 (1.28) | 0.095 |
| $t_{max}$ (h)[b] | 2 (1 – 4) | 2 (1 – 3) | 0.936 | 2 (1 – 4) | 2 (1 – 3) | 0.968 |
| $t_{½}$ (h) | 1.48 (1.17)[c] | 1.52 (1.29)[d] | 0.706 | 1.33 (1.16)[g] | 1.47 (1.34) | 0.366 |
| $\lambda_z$ (/h) | 0.47 (1.17)[c] | 0.46 (1.29)[d] | 0.936 | 0.52 (1.16)[g] | 0.47 (1.34) | 0.55 |
| CL/F (L/h) | 512.34 (1.46)[c] | 703.16 (1.60)[d] | 0.16 | 430.77 (1.68)[g] | 534.18 (1.31) | 0.34 |
| Vz/F (L) | 1095.79 (1.49)[c] | 1544.02 (1.63)[d] | 0.371 | 824.82 (1.68)[g] | 1132.01 (1.46) | 0.233 |
| Last dose | | | | | | |
| $AUC_{0\text{-last}}$ (h·ng/mL) | 147.28 (1.62) | 59.06 (1.83) | 0.001 | 353.76 (1.75) | 161.92 (1.63) | 0.005 |
| $AUC_{0\text{-inf}}$ (h·ng/mL) | 172.21 (1.69)[e] | 96.90 (1.46)[f] | 0.043 | 360.59 (1.74) | 165.82 (1.68)[h] | 0.008 |
| $C_{max}$ (ng/mL) | 25.65 (2.13) | 8.96 (2.46) | 0.019 | 71.05 (2.30) | 27.89 (2.17) | 0.019 |
| $t_{max}$ (h)[b] | 1.93 (1.62 – 5.93) | 1.92 (1.47 – 6.00) | 0.97 | 1.93 (1.62 – 5.93) | 1.97 (1.47 – 6.00) | 0.676 |

$AUC_{0\text{-last}}$ = area under the curve from 0 to last measurable concentration, $AUC_{0\text{-inf}}$ = area under the curve from 0 to infinity, $C_{max}$ = maximum concentration, $t_{max}$ = time at maximum concentration, $t_{½}$ = elimination half-life, $\lambda_z$ = first-order terminal elimination rate constant, CL/F = apparent total clearance, Vz/F = apparent total volume of distribution.

[a] Values in geometric mean (geometric standard deviation) unless otherwise stated.

[b] Values in median (range).

[c] Values estimated in 7 of 9 subjects.

[d] Values estimated in 7 of 11 subjects.

[e] Values estimated in 8 of 9 subjects.

[f] Values estimated in 5 of 11 subjects.

[g] Values estimated in 6 of 9 subjects.

[h] Values estimated in 10 of 11 subjects.

**Table S7. Pharmacokinetic parameters for lumefantrine and desbutyl-lumefantrine by treatment group following multiple-dose administration**

| Parameters[a] | Lumefantrine | | | Desbutyl-lumefantrine | | |
|---|---|---|---|---|---|---|
| | Placebo (n = 9) | Ruxolitinib (n = 11) | *p*-value | Placebo (n = 9) | Ruxolitinib (n = 11) | *p*-value |
| First dose | | | | | | |
| $AUC_{0\text{-last}}$ (h·ng/mL) | 12975.43 (2.11) | 13910.97 (1.52) | 0.649 | 21.06 (2.30) | 23.16 (1.38) | 0.939 |
| $C_{max}$ (ng/mL) | 3285.29 (2.17) | 3715.02 (1.56) | 0.494 | 6.11 (2.49) | 6.78 (1.39) | 0.82 |
| $t_{max}$ (h)[b] | 6.00 (5.00 – 8.00) | 6.00 (5.00 – 8.07) | 0.747 | 8.00 (5.00 – 8.08) | 8.00 (6.00 – 8.07) | 0.532 |
| $t_{lag}$ (h)[b] | 1.00 (0 – 1.05) | 1.00 (1.00 – 1.03) | 0.272 | 3.00 (1.00 – 3.02) | 3.00 (2.00 – 3.00) | 0.26 |
| Last dose | | | | | | |
| $AUC_{0\text{-last}}$ (h·ng/mL) | 683761.20 (1.73) | 692812.90 (2.43) | 0.63 | 12741.52 (1.66) | 10200.41 (2.71) | 0.615 |
| $AUC_{0\text{-inf}}$ (h·ng/mL) | 706105.60 (1.72) | 901768.90 (1.62)[c] | 0.353 | 14922.87 (1.61)[f] | 15090.50 (1.31)[g] | 0.829 |
| $C_{168}$ (ng/mL) | 1042.40 (2.08) | 1264.19 (1.88)[c] | 0.653 | 34.00 (1.66) | 36.19 (1.42)[c] | 0.488 |
| $C_{max}$ (ng/mL) | 12671.10 (1.73) | 13718.63 (1.59) | 0.843 | 95.56 (1.96) | 92.94 (1.35) | 0.82 |
| $t_{max}$ (h)[b] | 6.07 (5.85 – 21.93) | 6.00 (0 – 21.58) | 0.319 | 21.85 (5.95 – 22.45) | 20.47 (0 – 21.93) | 0.138 |
| $t_{½}$ (h) | 96.49 (1.18)[d] | 74.01[e] | NE | 161.11 (1.43)[f] | 178.66 (1.13)[h] | 0.826 |
| $\lambda_z$ (/h) | 0.01 (1.18)[d] | 0.01[e] | NE | 0.004 (1.43)[f] | 0.004 (1.13)[h] | 0.826 |
| CL/F (L/h) | 0.49 (1.56)[d] | 0.33[e] | NE | 38.28 (1.38)[d] | 28.43 (1.31)[g] | 0.156 |
| Vz/F (L) | 68.34 (1.76)[d] | 34.92[e] | NE | 7039.25 (1.68)[d] | 6954.25 (1.30)[g] | 0.796 |

$AUC_{0\text{-last}}$ = area under the curve from 0 to last measurable concentration, $AUC_{0\text{-inf}}$ = area under the curve from 0 to infinity, $C_{168}$ = concentration at 168 hours post-first dose, $C_{max}$ = maximum concentration, $t_{max}$ = time at maximum concentration, $t_{lag}$ = lag absorption time, $t_{½}$ = elimination half-life, $\lambda_z$ = first-order terminal elimination rate constant, CL/F = apparent total clearance, Vz/F = apparent total volume of distribution, NE = not estimated.

[a] Values in geometric mean (geometric standard deviation) unless otherwise stated.

[b] Values in median (range).

[c] Values estimated in 10 of 11 subjects.

[d] Values estimated in 3 of 9 subjects.

[e] Value estimated in 1 of 11 subjects.

[f] Values estimated in 5 of 9 subjects.

[g] Values estimated in 6 of 11 subjects.

[h] Values estimated in 8 of 11 subjects.

**Table S8. Individual participant parasite clearance parameters following treatment with artemether-lumefantrine plus ruxolitinib/placebo**

| Subject | Iter. | $R^2$ | p-value | $\hat{\beta}_1$ | $SE(\hat{\beta}_1)$ | $PRR_{48}(95\%CI)$ | Half-life [h] (95% CI) | log10$PRR_{48}(95\%CI)$ |
|---|---|---|---|---|---|---|---|---|
| Ruxolitinib | | | | | | | | |
| R001 | 0 | 95.20% | <0.001 | -0.066 | 0.004 | 1513 (661-3464) | 4.54 (4.08-5.12) | 3.18 (2.82-3.54) |
| R003 | 0 | 82.50% | <0.001 | -0.064 | 0.007 | 1141 (233-5582) | 4.73 (3.86-6.10) | 3.06 (2.37-3.75) |
| R004 | 0 | 92.10% | <0.001 | -0.077 | 0.006 | 5148 (1318-20117) | 3.89 (3.36-4.63) | 3.71 (3.12-4.30) |
| R007 | 3 | 99.60% | <0.001 | -0.15 | 0.004 | 15730310 (7056400-35066413) | 2.01 (1.92-2.11) | 7.20 (6.85-7.54) |
| *R010^* | *1* | *90.10%* | *0.004* | *-0.119* | *0.02* | *506823 (7170-35823885)* | *2.53 (1.91-3.75)* | *5.70 (3.86-7.55)* |
| R012 | 4 | 99.60% | <0.001 | -0.072 | 0.002 | 2872 (1886-4372) | 4.18 (3.97-4.41) | 3.46 (3.28-3.64) |
| *R013^* | *2* | *97.90%* | *0.011* | *-0.188* | *0.019* | *1013372933 (15195707-67579921164)* | *1.60 (1.33-2.01)* | *9.01 (7.18-10.83)* |
| R014 | 1 | 98.10% | <0.001 | -0.069 | 0.004 | 2076 (879-4908) | 4.36 (3.91-4.91) | 3.32 (2.94-3.69) |
| R017 | 2 | 99.40% | <0.001 | -0.069 | 0.002 | 1945 (1198-3156) | 4.39 (4.13-4.69) | 3.29 (3.08-3.50) |
| R018 | 0 | 90.20% | <0.001 | -0.069 | 0.009 | 1992 (311-12756) | 4.38 (3.52-5.80) | 3.30 (2.49-4.11) |
| Placebo | | | | | | | | |
| R002 | 0 | 96.50% | <0.001 | -0.085 | 0.005 | 12646 (4594-34810) | 3.52 (3.18-3.95) | 4.10 (3.66-4.54) |
| R005 | 3 | 94.30% | <0.001 | -0.073 | 0.005 | 3319 (1128-9764) | 4.10 (3.62-4.73) | 3.52 (3.05-3.99) |
| R006 | 4 | 97.70% | <0.001 | -0.099 | 0.005 | 59411 (19674-179406) | 3.03 (2.75-3.37) | 4.77 (4.29-5.25) |
| R008 | 1 | 96.70% | <0.001 | -0.093 | 0.007 | 29890 (6457-138369) | 3.23 (2.81-3.79) | 4.48 (3.81-5.14) |
| R009 | 1 | 91.50% | <0.001 | -0.071 | 0.009 | 2647 (386-18145) | 4.22 (3.39-5.59) | 3.42 (2.59-4.26) |
| R011 | 0 | 94.10% | <0.001 | -0.067 | 0.006 | 1650 (456-5977) | 4.49 (3.83-5.44) | 3.22 (2.66-3.78) |
| R015 | 1 | 98.80% | <0.001 | -0.06 | 0.002 | 757 (457-1254) | 5.02 (4.66-5.43) | 2.88 (2.66-3.10) |
| R016 | 1 | 96.60% | <0.001 | -0.067 | 0.005 | 1580 (523-4770) | 4.52 (3.93-5.31) | 3.20 (2.72-3.68) |
| R017 | 2 | 99.40% | <0.001 | -0.069 | 0.002 | 1945 (1198-3156) | 4.39 (4.13-4.69) | 3.29 (3.08-3.50) |
| R019 | 3 | 99.80% | <0.001 | -0.067 | 0.001 | 1633 (1222-2183) | 4.50 (4.33-4.68) | 3.21 (3.09-3.34) |
| R020 | 2 | 99.70% | <0.001 | -0.094 | 0.002 | 33400 (19791-56367) | 3.19 (3.04-3.36) | 4.52 (4.30-4.75) |

$PRR_{48}$: Parasite reduction ratio over a 48-hour period. ^Subjects in italics had non-significant regression fits and were not included in cohort-specific $PRR_{48}$ estimate.

**Table S9. pCyTOF whole blood staining panel**

| Phospho-CyTOF | | | | |
|---|---|---|---|---|
| **Metal label** | **Specificity** | **Clone** | **Supplier** | **Cat#** |
| **89Y** | CD45 | HI30 | Standard BioTools | 3089003B |
| **141Pr** | CD7 | CD7-6B7 | Biolegend | 343111 |
| **142Nd** | CD19 | HIB19 | Standard BioTools | 3142001B |
| **143Nd** | CD45RA | HI100 | Standard BioTools | 3143006B |
| **144Nd** | pPLCg2 | K86-689.37 | Standard BioTools | 3144015A |
| **145Nd** | CD4 | RPA-T4 | Standard BioTools | 3145001B |
| **146Nd** | IgD | IA6-2 | Standard BioTools | 3146005B |
| **147Sm** | CD20 | 2H7 | Standard BioTools | 3147007B |
| **148Nd** | IgA | Polyclonal | Standard BioTools | 3148007B |
| **149Sm** | CD25 | 2A3 | Standard BioTools | 3149010B |
| **150Nd** | pStat5 | 47 | Standard BioTools | 3150005A |
| **151Eu** | CD123 | 6H6 | Standard BioTools | 3151001B |
| **152Sm** | Akt | D9E | Standard BioTools | 3152005A |
| **153Eu** | pStat1 | 4a | Standard BioTools | 3153005A |
| **154Sm** | CD127 | R34-34 | Novus Biological | DDX0700P-100 |
| **155Gd** | CD27 | L128 | Standard BioTools | 3155001B |
| **156Gd** | pP38 | D3F9 | Standard BioTools | 3156002A |
| **157Gd** | CD24 | ML-5 | Biolegend | 311102 |
| **158Gd** | pStat3 | 4 | Standard BioTools | 3158005A |
| **159Tb** | CD11c | Bu15 | Standard BioTools | 3159001B |
| **160Gd** | CD14 | M5E2 | Standard BioTools | 3160001B |
| **162Dy** | CD66b | 80H3 | Standard BioTools | 3162023B |
| **163Dy** | CD56 | NCAM16.2 | Standard BioTools | 3163007B |
| **164Dy** | IkBalpha | L35A5 | Standard BioTools | 3164004A |
| **165Ho** | CD16 | B73.1 | Standard BioTools | 3167001B |
| **167Er** | CD38 | HIT2 | Standard BioTools | 3168002B |
| **168Er** | CD8 | SK1 | Standard BioTools | 3169010B |
| **169Tm** | CD33 | WM53 | Standard BioTools | 3170001B |
| **170Er** | CD3 | UCHT1 | Standard BioTools | 3167001B |
| **171Yb** | pERK 1/2 | D13.14.4E | Standard BioTools | 3171010A |
| **172Yb** | Ki67 | B56 | Standard BioTools | 3172024B |
| **173Yb** | CD21 | Bu32 | Biolegend | 354902 |
| **174Yb** | HLA-DR | L243 | Standard BioTools | 3174001B |
| **175Lu** | pS6 | N7548 | Standard BioTools | 3175009A |
| **176Yb** | CREB | 87G3 | Standard BioTools | 3176005A |
| **209Bi** | CD11b | ICRF44 | Standard BioTools | 3209003B |

**Table S10. Segmented regression sine-wave mixed models of parasite growth (N=300, Subjects =20)**

| | Treatment Group Interaction Model | | Treatment Group Main effects Model | |
|---|---|---|---|---|
| **Parameter** | **Estimate (95% CI)** | **p-value** | **Estimate (95% CI)** | **p-value** |
| a | -2.06 (-2.41– -1.71) | <0.001 | -2.06 (-2.41– -1.71) | <0.001 |
| m (time) | 0.76 (0.71 – 0.81) | <0.001 | 0.76 (0.71 – 0.81) | <0.001 |
| g (group) | 0.37 (-0.32 – 1.06) | 0.30 | 0.30 (-0.24 – 0.85) | 0.28 |
| p (phase) | -4.27 (-4.80 – -3.75) | <0.001 | -4.24 (-4.71 – -3.76) | <0.001 |
| pt (phase x time) | -0.07 (-0.15 – 0.01) | 0.071 | -0.08 (-0.14 – -0.02) | 0.009 |
| pgt (phase x group x time) | -0.01 (-0.11 – 0.08) | 0.76 | - | - |
| $c$ | -0.56 (-0.63 – -0.50) | <0.001 | -0.56 (-0.63 – -0.50) | <0.001 |
| $Period\ (days)$ | 1.68 (1.65 – 1.70) | <0.001 | 1.68 (1.65 – 1.70) | <0.001 |
| $k$ | 3.78 (3.36 – 4.20) | <0.001 | 3.78 (3.36 – 4.19) | <0.001 |
| Random effects | | | | |
| Intercept: a | 0.40 (0.26 - 0.61) | | 0.40 (0.26 - 0.61) | |
| Intercept: p | 0.41 (0.21 – 0.80) | | 0.41 (0.22 – 0.78) | |
| Slope: m | 0.03 (0.01 - 0.10) | | 0.03 (0.01 - 0.10) | |
| Slope: pt | 0.04 (0.01 - 0.13) | | 0.04 (0.01 - 0.13) | |
| Residual | 0.36 (0.33 - 0.39) | | 0.36 (0.33 - 0.39) | |
| Model AIC | 396.9 | | 370.9 | |
| Deviance (-2xLog-likelihood) | 368.9 | | 364.7 | |

^growth rate for reinfection phase (m + pt): 0.68 (95% CI 0.63 - 0.73)

### Supplementary Figures

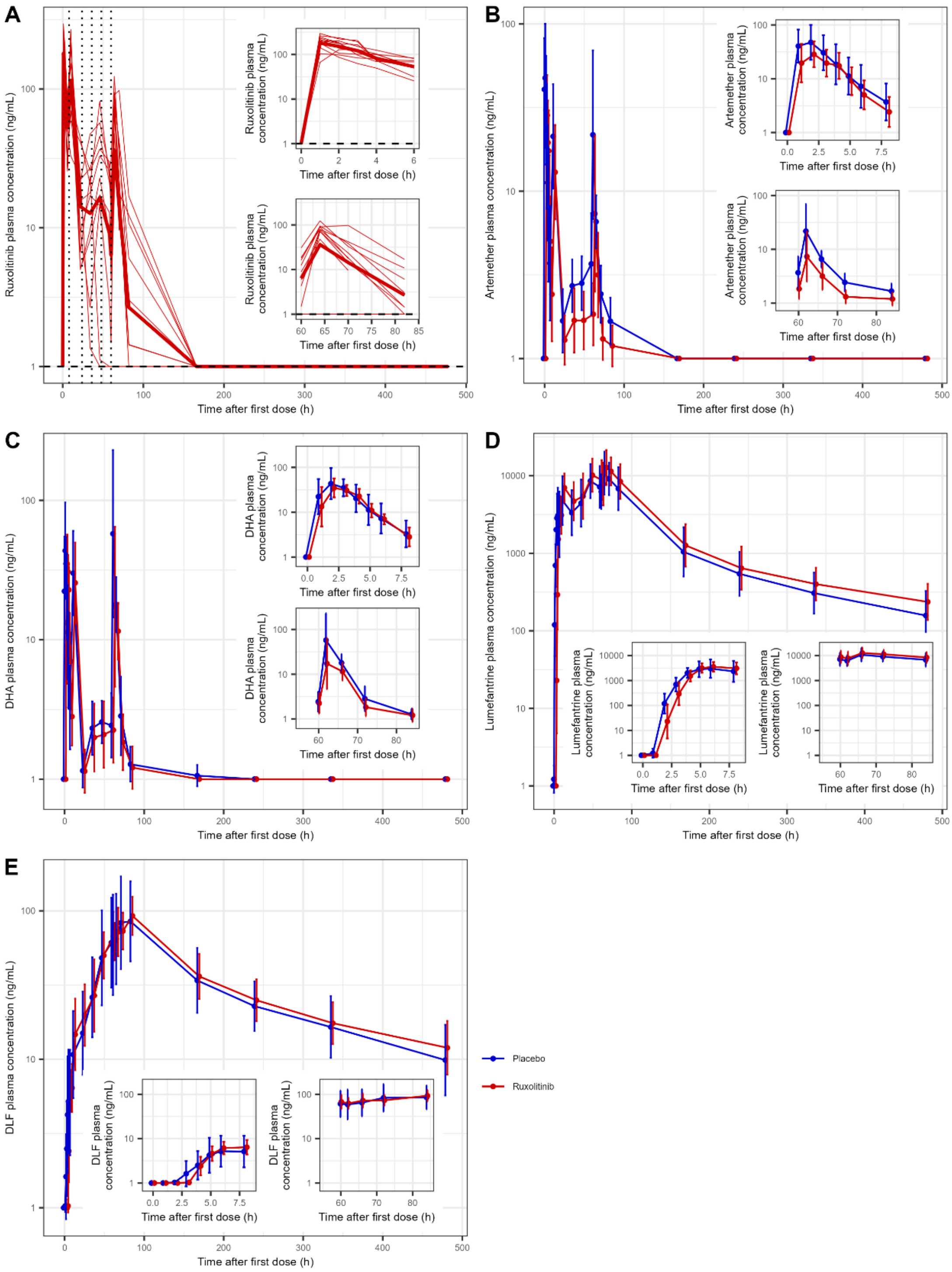


**Figure S1. Impact of ruxolitinib treatment on artemether, lumefantrine, and metabolite concentrations.**

Individual participant ruxolitinib plasma concentrations over time on a log10 scale **(A).** Geometric mean plasma concentrations of artemether **(B)**, lumefantrine **(C)**, dihydroartemisinin (DHA) **(D)** and desbutyl-lumefantrine (DLF) (E) on a log10 scale. Points represent geometric mean, and bars represent standard deviation.

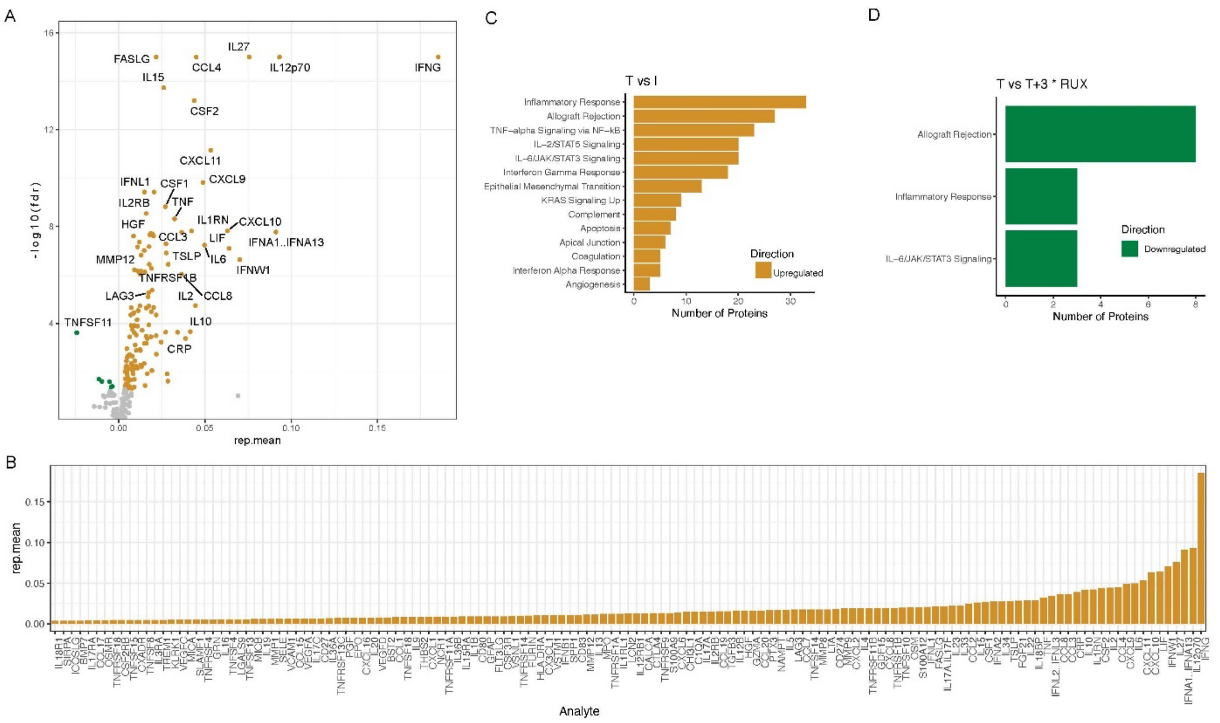


**Figure S2. NULisa analysis of inflammatory responses.**

A) Volcano plot of coefficient (rep.mean) and FDR (-log10) of change of analyte at T compared to I prior to randomisation. Coefficient and FDR values are from linear mixed effect model analysis. Analytes that are upregulated are in gold, and down regulated in green. B) Significantly upregulated analytes for change at T compared to I, ordered by coefficient values. C/D) Overrepresentation pathway analysis of analytes with a significant (FDR<0.05) change in C) at T compared to I prior to randomisation, and D) of interaction term between ruxolitinib treatment group and change of analyte at T+3 compared to T. Hallmark gene sets was used for the pathway analysis. Significant pathways (FDR<0.05) are shown, with upregulated pathways in gold and downregulated pathways in green.
