## Supplementary material for "Adjunctive ruxolitinib attenuates inflammation and enhances antiparasitic immunity in human volunteers experimentally infected with *Plasmodium falciparum*": Clinical Trial Protocol

**A randomised, double blind, placebo controlled trial to evaluate the safety, tolerability and anti-parasitic immunity boosting activity of ruxolitinib when co-administered with artemether-lumefantrine in healthy volunteers with *Plasmodium falciparum* Induced Blood Stage Malaria**

**Protocol Identifying Number: CTM2003**

**Principal Investigator: Associate Professor Bridget Barber**

**Trial Sponsor: QIMR Berghofer Medical Research Institute**

**Version Number: Version 5.0**

**04 Jan 2023**

CONFIDENTIALITY STATEMENT

This document contains information that is privileged or confidential. As such, it may not be disclosed unless specific prior permission is granted in writing by QIMRB Medical Research Institite or such disclosure is required by federal or other laws or regulations. Persons to whom any of this information is to be disclosed must first be informed that the information is confidential. These restrictions on disclosure will apply equally to all future information supplied, which is indicated as privileged or confidential.

### STATEMENT OF COMPLIANCE

#### Investigator declaration

I have read the protocol and agree that it contains all necessary details for carrying out the trial as described. I will conduct this protocol as outlined herein and will make a reasonable effort to complete the trial within the time designated.

I agree to personally conduct or supervise the described trial.

The trial will be conducted in accordance with the following:

- World Medical Association Declaration of Helsinki – Ethical Principles for Medical Research Involving Human Volunteers (Fortaleza, Brazil 2013)
- NHMRC National Statement on Ethical Conduct in Human Research (2007, updated 2018)
- Integrated Addendum to ICH E6 (R1): Guideline for Good Clinical Practice E6 (R2) (November 2016) – with introductory comments of the Australian Therapeutic Goods Administration
- Most current Clinical Trial Protocol approved by the relevant Human Research Ethics Committee(s)

I agree to inform all volunteers that the trial interventions are being used for investigational purposes and I will ensure that the requirements related to obtaining informed consent are in accordance with International Council for Harmonisation (ICH) Guidelines for Good Clinical Practice (GCP) Section 4.8 and local requirements.

I agree to report adverse events that occur in the course of the trial to the Sponsor in accordance with ICH Guidelines for GCP Section 4.11 and local requirements.

I have read and understand the information in the Investigator's Brochures, including the potential risks and side effects of the trial drug.

I agree to promptly report to the approving Human Research Ethics Committee(s) all changes in the research activity and all unanticipated problems involving risk to volunteers. I will not make any changes to the conduct of the trial without HREC and Sponsor approval, except when necessary to eliminate apparent immediate harm to volunteers.

I agree to maintain adequate and accurate records and make those records available in accordance with ICH Guidelines for GCP Section 4.11 and local requirements.

I agree to ensure that all associates, colleagues, and employees assisting in the conduct of the trial are informed about their obligations in meeting the above commitments.

I understand that the trial may be terminated or enrolment suspended at any time by the Sponsor, with or without cause, or by me if it becomes necessary to protect the best interest of the volunteers.

_______________________________ Date: ______________________________

A/Prof. Bridget Barber, Principal Investigator

#### Signatories

The undersigned parties agree that the protocol was written in accordance with the World Medical Association Declaration of Helsinki — Ethical Principles for Medical Research Involving Human Volunteers (Fortaleza, Brazil 2013), the NHMRC National Statement on Ethical Conduct in Human Research (2007, updated 2018), and the Integrated Addendum to ICH E6 (R1): Guideline for Good Clinical Practice E6 (R2) (November 2016) — with introductory comments of the Australian Therapeutic Goods Administration.

**This clinical trial protocol has been prepared by the following QIMR Berghofer personnel.**

| Name | Signature | Date |
|---|---|---|
| **Rebecca Webster PhD**<br>Clinical Operations Manager<br>QIMR Berghofer Medical Research Institute | | |

**This clinical trial protocol has been reviewed and approved by the Sponsor.**

| Name | Signature | Date |
|---|---|---|
| Professor Grant Ramm PhD<br>Deputy Director<br>QIMR Berghofer Medical Research Institute | | |

### 1 PROTOCOL SUMMARY

#### 1.1 SYNOPSIS

**Title:**

A randomised, double blind, placebo controlled trial to evaluate the safety, tolerability and anti-parasitic immunity boosting activity of ruxolitinib when co-administered with artemether-lumefantrine in healthy volunteers with *Plasmodium falciparum* Induced Blood Stage Malaria.

**Trial description:**

This is a, randomised, double-blind, placebo-controlled, phase 1b trial to assess the safety, tolerability, pharmacokinetic (PK), and pharmacodynamic (PD; malaria parasitaemia 18S qPCR, pSTAT3, and immune responses) of artemether-lumefantrine (AL)+ Ruxolitinib (Rux) in healthy adults with *P. falciparum* IBSM.

Twenty-six malaria-naïve, healthy males or females, aged between 18-55 years old, who meet all of the inclusion criteria and none of the exclusion criteria, are planned to be enrolled. Volunteers will be randomised in a 1:1 ratio to receive oral twice daily doses of AL+Rux or AL+placebo on Days 8, 9 and 10. A sentinel dosing strategy will be used whereby two volunteers (one randomised to AL+Rux and one randomised to AL+placebo) will be dosed initially. The Safety Data Review Team will review safety and tolerability data up to and including Day 16 before dosing of the remaining 24 volunteers.

As part of the informed consent process, volunteers will be asked if they agree to be contacted at approximately 3, 6 and 12 months after their second inoculation for blood sampling to investigate anti-parasitic immune response longevity.

**First Inoculation:**

All volunteers enrolled will receive a first inoculation on Day 0 with ~2,800 viable parasites of *P. falciparum*-infected human erythrocytes administered intravenously. Blood sampling will occur for baseline PD parameters (malaria parasitaemia 18S qPCR, pSTAT3 and immune responses) on Day 0 prior to inoculation. Volunteers will be followed up daily via phone call or text message on Days 1 to 3 post inoculation to solicit the occurrence of any adverse events (AEs).

Volunteers will then attend the clinical trial unit once daily from Day 4 until presence of asexual parasites is established by 18S qPCR. Once malaria 18S qPCR becomes positive, and until AL+Rux or AL+placebo administration, volunteers will attend the clinical trial unit twice-daily, separated by approximately 12 h, for clinical evaluation and blood sampling.

Volunteers will be admitted to the clinical trial unit on Day 8 when parasitaemia for the majority of volunteers is expected to be above 5,000 parasites/mL. Individual volunteers will be admitted to the clinical trial unit for earlier treatment if:

- they experience a serious adverse event (SAE) related to the malaria challenge agent, or
- they have a grade 3 AE graded in accordance with the Common Terminology Criteria for Adverse Events (CTCAE) deemed related to malaria and not self-resolved or relieved with concomitant medications, or
- the Investigator considers it necessary for volunteer safety

At the Investigator's discretion, in consultation with the medical monitor and Sponsor, administration of IMP to any or all of the participants may be delayed if parasitaemia levels on Days 6 and 7 are lower than expected. In this event, all subsequent trial time points will be adjusted accordingly.

Volunteers will be confined in the clinical unit for at least 72 h for twice daily dosing of AL+Rux or AL+placebo. During confinement blood sampling for safety, PK and PD (pSTAT3, malaria 18S qPCR and immune response) assessments will occur.

If adequate tolerance to dosing and clinical response is observed, volunteers will be discharged from the clinical trial unit at 72 hours, and will be followed up as outpatients for safety, and to monitor malaria parasitaemia via 18S qPCR. Volunteers will have safety, PK, malaria 18S qPCR, pSTAT3, and immune response assessment blood sampling up to Day 28±2.

**Second Inoculation:**

At Day 85±7 volunteers will have a repeat eligibility visit and the SDRT will assess safety, tolerability and RBC alloantibody status from all volunteers enrolled up to and including Day 85±7.

At Day 90±7, contingent upon SDRT decision, volunteers enrolled who remain eligible (meet all of the inclusion criteria and none of the exclusion criteria), and who do not demonstrate allo-immunisation, will be administered intravenously a second (homologous) inoculation of ~2,800 viable parasites of *P. falciparum*-infected human erythrocytes.

Volunteers will be followed up daily via phone call or text message on Days 91±7 to 93±7 to solicit any AEs.

Volunteers will then attend the clinical trial unit once daily from Day 94±7 until presence of asexual parasites is established by malaria 18S qPCR. Once malaria 18S qPCR becomes positive, and until AL administration, volunteers may be asked to attend the clinical trial unit up to twice-daily, separated by approximately 12 h, for clinical evaluation and blood sampling. In the event that parasitemia remains low and stable, clinic visits for blood sampling may be reduced to a minimum of 3 times per week, at the discretion of the Principal Investigator. Blood collection for immune response assessments may occur approximately three times per week from Day 94±7 until AL administration.

Volunteers will be administered the first dose of AL treatment at the clinical trial unit on an individualised basis, when:

- qPCR parasitaemia reaches ≥50,000 parasites/mL, or
- they have a malaria clinical score >6, and presence of parasitaemia, or
- they experience an SAE related to the malaria challenge agent, or
- they have a CTCAE grade 3 AE deemed related to malaria and not self-resolved or relieved with concomitant medications, or
- the Investigator considers it necessary for volunteer safety.
- **If none of the above criteria for AL administration are reached by Day 118±7 (28 days after second inoculation) then compulsory AL administration must occur.**

Volunteers will be followed up daily by via phone call or text message for 2 days to ensure adherence to AL twice daily dosing regimen.

Volunteers will have safety, tolerability, and PD (malaria 18S qPCR and immune response) assessments performed up to 28±3 days after AL treatment initiation.

Immune response assessment blood sampling may occur at the Investigator's discretion:

- within 3 h prior to the second inoculation on Day 90±7
- approximately three times per week from Day 94±7 until AL administration
- within 3 h prior to AL administration (volunteer parasitaemia reaches ≥50,000 parasites/mL or volunteer malaria clinical score reaches >6 and presence of parasitaemia), and
- 3±2, 7±3, 20±3, and 28±3 days after AL treatment initiation.

Time-points for immune response assessments may change based on data analysis from previous cohorts at the Investigator's discretion (refer to Lab Manual for each cohort).

Malaria 18S qPCR blood sampling will occur:

- pre-AL administration (volunteer parasitaemia reaches ≥50,000 parasites/mL or volunteer malaria clinical score reaches > 6 and presence of parasitaemia), and
- 3±2 and 7±3 days post AL administration or until two consecutive negative malaria 18S qPCR results are obtained, or
- at Investigator discretion

**Objectives and Endpoints:**

| **Primary objective** | **Primary endpoint** |
|---|---|
| 1. To assess the safety and tolerability of 3-day twice daily dosing of AL+Rux with AL+placebo in the context of *P. falciparum* IBSM. | 1. Incidence, severity, and relationship of observed and self-reported AEs by treatment regimen. |
| **Secondary objectives** | **Secondary endpoints** |
| 1. To compare the anti-parasitic immune response of AL+Rux with AL+placebo in volunteers with *P. falciparum* IBSM. | 1. Anti-parasitic immune response by treatment regimen and challenge phase:<br>• Parasite-specific IFNγ and IL-10 levels<br>• Other host immune responses to infection (**appendix 3**) |
| 2. To compare the effect of AL+Rux with AL+placebo on pSTAT3 inhibition. | 2. pSTAT3 inhibition will be assessed *ex-vivo* on whole blood cells. |
| 3. To characterise the PK profile of artemether and its major metabolite dihydroartemisinin [DHA], lumefantrine, and Rux. | 3. PK parameters of artemether, DHA, lumefantrine, and Rux using non-compartmental methods: $AUC_{last}$, $AUC_{0-\infty}$, $C_{max}$ (first and last dose), $t_{max}$ (first and last dose), elimination $t_{1/2}$, $t_{lag}$, $C_{168h}$ (for lumefantrine only), CL/F, Vz/F and λz in all volunteers. |
| 4. To compare the antimalarial effect of 3-day twice daily dosing of AL+Rux with AL+placebo in volunteers with *P. falciparum* IBSM. | 4. Antimalarial activity assessed by the following parameters:<br>• Parasite clearance half-life<br>• Parasite reduction ratio<br>• Percentage of volunteers with recrudescence of parasitaemia. |
| 5. To compare the effect of AL+Rux with AL+placebo on parasite growth following a second homologous *P. falciparum* IBSM infection. | 5. Parasite growth will be characterised by:<br>• 'time to parasitaemia': the first time-point that parasites are detected<br>• parasite multiplication rate (PMR) |
| **Exploratory objectives** | **Exploratory endpoints** |
| 1. To investigate the longevity of anti-parasitic immune responses in volunteers. | 1. Anti-parasitic immune response up to 16 months post-IMP administration by treatment regimen:<br>• Parasite-specific IFNγ and IL-10 responses.<br>• Other host immune responses to infection (**appendix 3**) |

| |
|---|
| **Population:**<br><br>Up to 26 volunteers are planned to be enrolled in this trial. Volunteers will be malaria-naïve, healthy males or (non-lactating, non-pregnant) females who agree to use a strict double method of contraception for the duration of the trial. Volunteers will be aged between 18–55 years old and will meet all of the inclusion and none of the exclusion criteria. |
| **Clinical phase:** 1b |
| **Number of sites enrolling volunteers:**<br>The trial is planned to be performed at the University of Sunshine Coast Clinical Trials Centre.<br>Additional sites in Australia may be added if necessary. |
| **Duration of trial:**<br><br>It is estimated that the clinical portion (from site initiation visit [SIV] to last volunteer last visit [LSLV]) of the trial will be completed within 24 months. |
| **Duration of volunteer participation:**<br><br>Approximately 5 months. This includes a screening period of up to 28 days, and approximately 4 months of 'on study' time. For volunteers that consent to be contacted at 3, 6 and 12 months post their second inoculation visit the estimated time-frame is 17 months. |

#### 1.2 SCHEMA

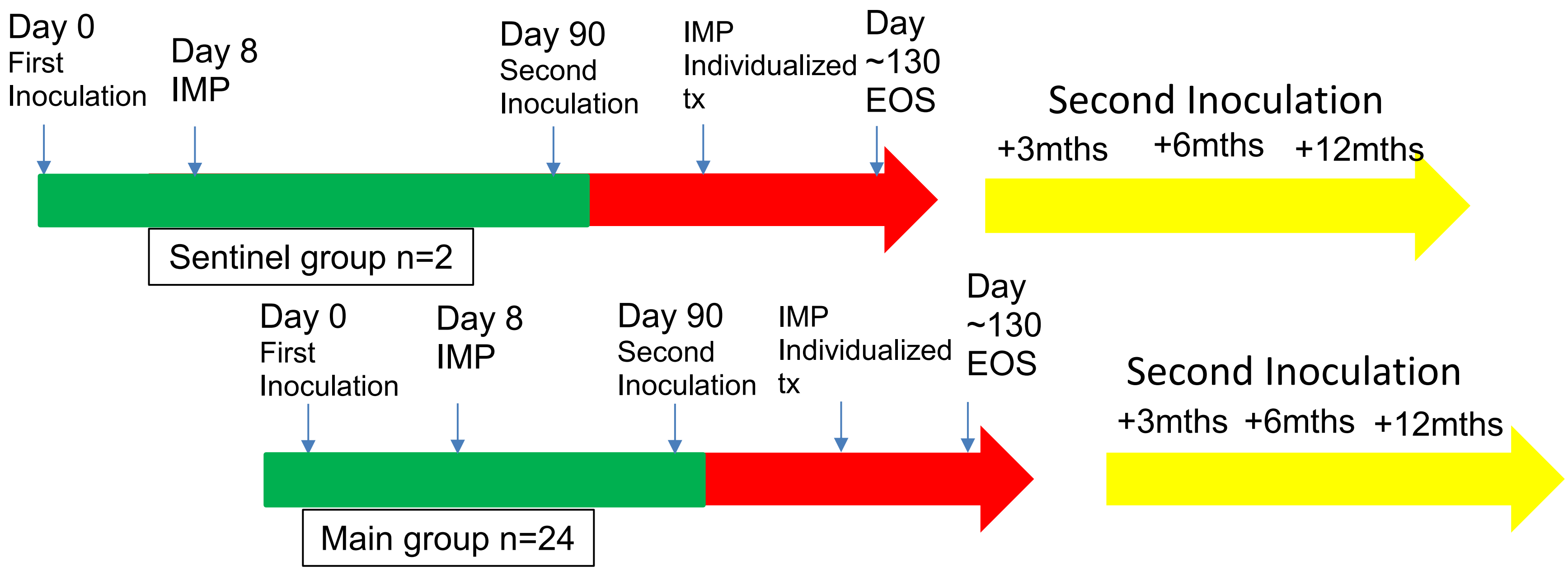

**Colour Key & abbreviations**

Green = First inoculation
Red = Second inoculation
Yellow = Optional sample collection
EOS = End of Study
IMP = Investigational medicinal product

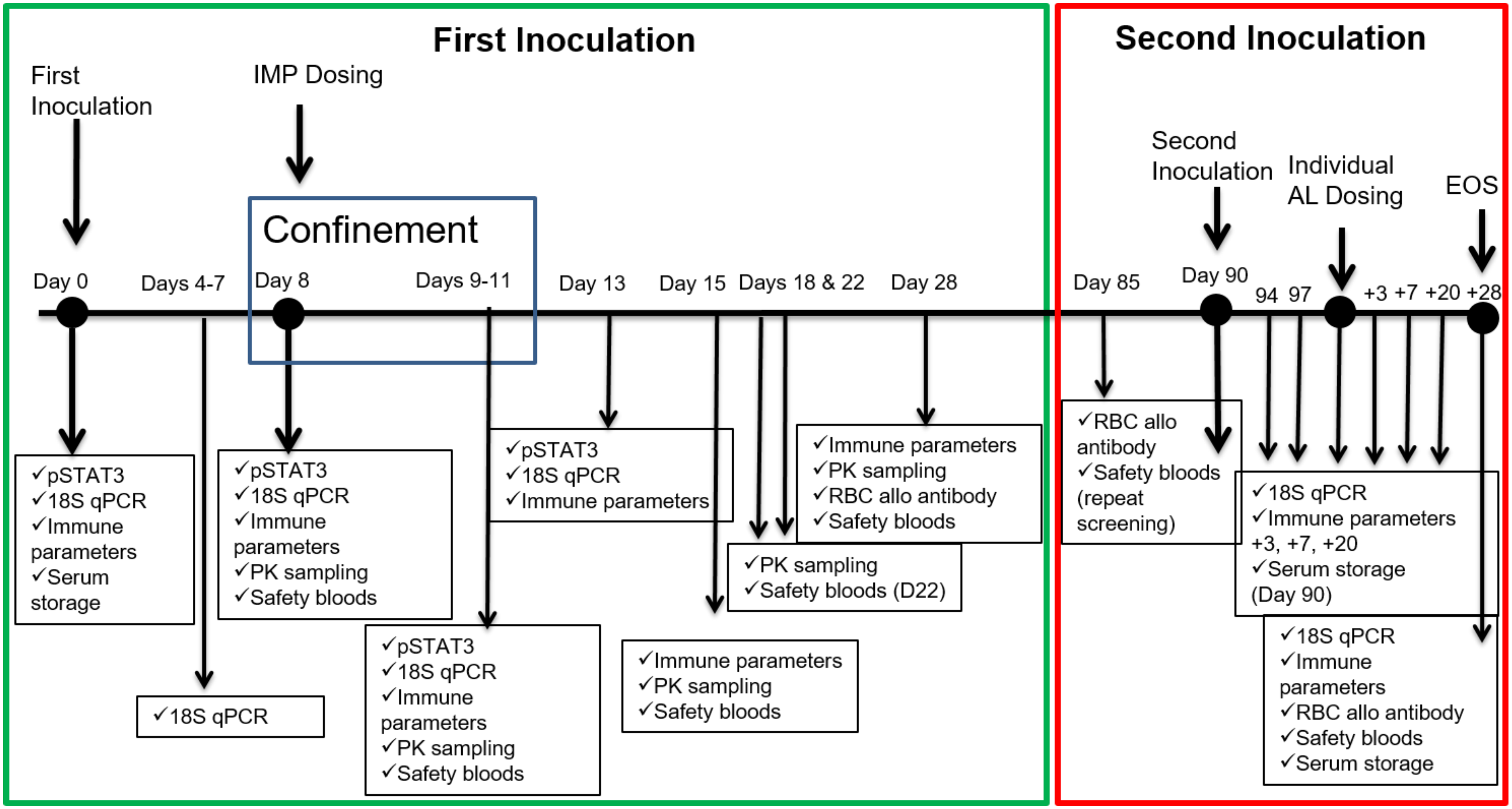
First Inoculation
Second Inoculation
First Inoculation
IMP Dosing
Confinement
Day 0
Days 4-7
Day 8
Days 9-11
Day 13
Day 15
Days 18 & 22
Day 28
Day 85
Second Inoculation
Day 90
Individual AL Dosing
EOS
94 97
+3 +7 +20 +28
✓pSTAT3 ✓18S qPCR ✓Immune parameters ✓Serum storage
✓18S qPCR
✓pSTAT3 ✓18S qPCR ✓Immune parameters ✓PK sampling ✓Safety bloods
✓pSTAT3 ✓18S qPCR ✓Immune parameters ✓PK sampling ✓Safety bloods
✓pSTAT3 ✓18S qPCR ✓Immune parameters
✓Immune parameters ✓PK sampling ✓Safety bloods
✓PK sampling ✓Safety bloods (D22)
✓Immune parameters ✓PK sampling ✓RBC allo antibody ✓Safety bloods
✓RBC allo antibody ✓Safety bloods (repeat screening)
✓18S qPCR ✓Immune parameters +3, +7, +20 ✓Serum storage (Day 90)
✓18S qPCR ✓Immune parameters ✓RBC allo antibody ✓Safety bloods ✓Serum storage

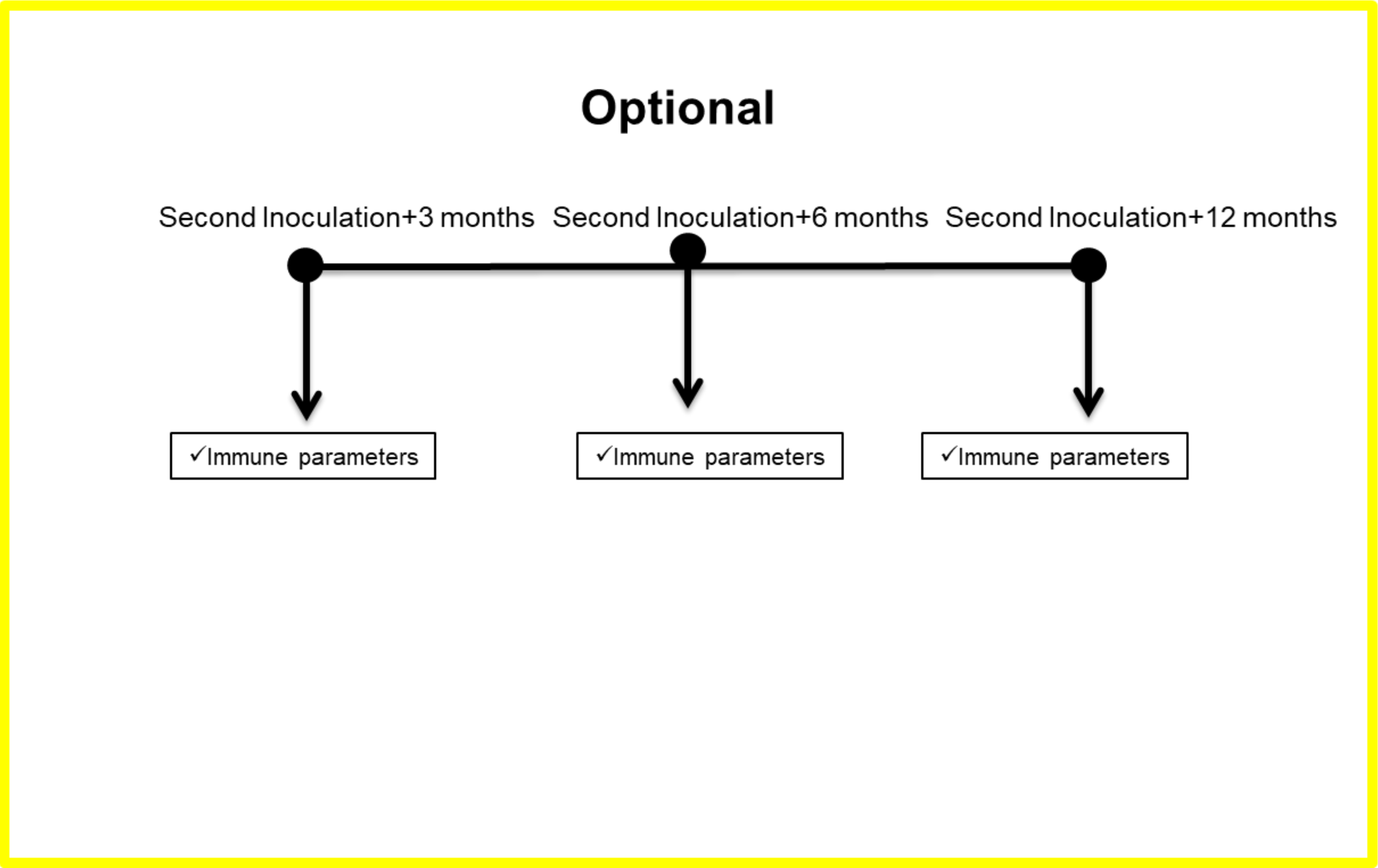
Optional
Second Inoculation+3 months
Second Inoculation+6 months
Second Inoculation+12 months
✓Immune parameters
✓Immune parameters
✓Immune parameters

#### 1.3 SCHEDULE OF ACTIVITIES DETAILS

##### 1.3.1 FIRST INOCULATION

###### 1.3.1.1 SCREENING: DAYS -28 TO DAY -1

A screening visit will be scheduled after an initial phone interview conducted by clinical trial unit staff has occurred to review background information. For the screening visit, potential volunteers will be asked to come to the clinical trial unit after an overnight fast of ≥8 h. During this initial screening visit, an Investigator will discuss the details of the trial with the potential volunteer, and the volunteer will read the Participant Information Sheet and be encouraged to ask questions. The potential volunteer will be fully informed of the nature of the trial at this time.

Volunteers willing to be considered for inclusion may sign the Informed Consent Form during the screening visit or may return to the clinical trial unit after further consideration of the trial and the Informed Consent Form. The volunteer will be given a copy of the Participant Information Sheet and signed Informed Consent Form for their records.

The signed and dated originals will be held on file by the clinical trial unit. Participation consent must be obtained from all volunteers prior to screening tests. Once the potential volunteer has provided written consent to participate, the pre-trial screening may be initiated.

Volunteers who complete all screening procedures and satisfy all entry criteria will be considered eligible to participate in this trial. For laboratory parameters, a repeat may be requested to exclude laboratory error and/or to confirm eligibility. Re-screening will not be allowed unless the Investigator, in agreement with the independent medical monitor, considers the cause of the initial pre-screening failure to be of an acute and completely reversible nature.

If screening laboratory results are abnormal (e.g., HIV testing), the volunteer will be referred for appropriate counselling. If any clinically significant abnormalities are detected during screening, the volunteer will be referred to a general practitioner or medical specialist for follow-up tests as appropriate.

**The screening will be conducted within 4 weeks prior to first inoculation**

1. Assign screening number to each volunteer.
2. Elicit a complete medical history and use of medications.
3. Elicit a social history including recreational drug, alcohol, and tobacco use.
4. Elicit demographic data.
5. Record body height and weight and calculate BMI.
6. Perform alcohol breath test.
7. Perform urine drug screen.
8. Perform full physical examination.
9. Ask volunteer to complete the Beck Depression Inventory.
10. Record vital signs (supine and standing blood pressure and heart rate).

11. Obtain a 12-lead ECG.
12. Collect urine for urinalysis.
13. Collect blood samples for
    a. haematology (including G6PD)
    b. biochemistry (including lipids, no CRP)
    c. serology (viral hepatitis B and C, HIV)
    d. coagulation profile
    e. QuantiFERON-TB Gold assay (tuberculosis testing)
    f. RBC alloantibodies testing
14. Perform serum β-hCG pregnancy test for all female subjects, and FSH test for post-menopausal females.
15. Assess the cardiovascular disease risk as per exclusion criterion.
16. Verify volunteer meets inclusion/exclusion criteria.
17. Educate volunteers on signs and symptoms of malaria.

###### 1.3.1.2 ELIGIBILITY VISIT: DAY -3 TO DAY -1

Volunteers will report to the clinical trial unit after an overnight fast of ≥8 h between Day -3 to Day -1 for the following baseline assessments, unless screening laboratory assessments were conducted within this period, in which case repeat sampling will not be required.

This visit will include:

1. Collect urine for urinalysis
2. Collect blood samples for
    a. haematology
    b. biochemistry (including iron studies)
    c. urine β-hCG pregnancy test for WOCBP

The timing of these assessments is to ensure that results are available for review by the Investigator prior to inoculation on inoculation day. Volunteers with clinically significant laboratory findings at this stage will not be eligible for inoculation.

###### 1.3.1.3 FIRST MALARIA INOCULATION: DAY 0

Every volunteer, including reserve volunteers will report to the clinical trial unit on the morning of Day 0. The Investigator will review the volunteers' screening and eligibility confirmation visit results prior to their enrolment into the trial and subsequent inoculation. The Investigator will emphasise the requirement to return to the clinical trial unit for antimalarial drug treatment after the malaria inoculation.

On admission to the clinical trial unit, volunteers will be required to undertake further procedures to determine whether they remain eligible to be enrolled. A reserve volunteer may be asked to replace a volunteer who no longer meets eligibility. The reserve volunteers will be compensated for the trial visit even if not inoculated, as described in the Participant Information Sheet and Informed Consent Form.

<u>The procedures that will be undertaken prior to inoculation on Day 0 include:</u>

1. Verify that all applicable eligibility criteria have been met.
2. Elicit information regarding any new medical conditions, illnesses and medication use since screening.
3. Perform alcohol breath test and urine drug screen.
4. Perform urine β-hCG pregnancy test for WOCBP.
5. Conduct abbreviated physical examination.
6. Record vital signs.
7. Obtain 12-lead ECG (baseline value).
8. Cannulate volunteers with an indwelling IV cannula for the malaria inoculum, and record which arm is used.
9. Collect blood samples for
    a. serum retention sample
    b. malaria 18S qPCR (baseline parasitaemia sample)
    c. pSTAT3 assay
    d. immune response assessments

<u>Administration of the malaria challenge agent:</u>

1. Administer the malaria challenge agent of ~2,800 viable *P. falciparum* 3D7 infected human RBCs intravenously in the morning.
2. Observe volunteer for a minimum of 60 min after inoculation to evaluate for immediate adverse reactions.
3. Re-educate volunteers of signs and symptoms of malaria.
4. Emphasise to volunteers the importance of returning on the nominated day (approximately Day 8), or as advised by the clinical trial unit staff, for antimalarial treatment.
5. Provide volunteers with diary cards to record symptoms and concomitant medications during the trial, and thermometers to record any temperature readings during the trial in the event of symptoms of fever.
6. Record vital signs prior to leaving the clinical trial unit (approximately 60 min after inoculation).
7. Record malaria clinical score prior to leaving the clinical trial unit (malaria clinical score baseline).

8. Record AEs and concomitant medications.

###### 1.3.1.4 MALARIA MONITORING VIA PHONE: DAY 1 TO DAY 3

During this period, the volunteers are expected to be asymptomatic. Clinical trial unit staff will contact volunteers via a daily phone call or text message to monitor volunteer wellbeing and to solicit any AEs and concomitant medications.

###### 1.3.1.5 MALARIA MONITORING OUTPATIENT VISITS: DAY 4 TO DAY 7

Follow-up from Day 4 to Day 7 will be undertaken through up to twice-daily (am and pm) visits to the clinical trial unit separated by approximately 12 h (e.g., 06.00–11.00 and 18.00–23.00).

The following procedures will occur during these visits:

1. Perform symptom-directed physical examination if clinically indicated at the Investigator's discretion.
2. Record vital signs.
3. Collect blood sample for malaria 18S qPCR.
4. Record malaria clinical score.
5. Check volunteer diary cards.
6. Record AEs and use of concomitant medications.

###### 1.3.1.6 OBSERVATION AND ANTIMALARIAL TREATMENT: DAY 8 TO DAY 11

Volunteers will be admitted to the clinical trial unit (after fasting for ≥8 h) for at least 72 h on the morning of Day 8 when parasitaemia for the majority of volunteers is expected to be more than 5,000 parasites/mL. Individual volunteers will be admitted to the clinical trial unit for earlier treatment if:

- they experience an SAE related to the malaria challenge agent, or
- they have a grade 3 AE graded in accordance with CTCAE deemed related to malaria and not self-resolved or relieved with concomitant medications, or
- the Investigator considers it necessary for volunteer safety.

At the Investigator's discretion, in consultation with the medical monitor and Sponsor, administration of IMP to any or all of the participants may be delayed if parasitaemia levels on Days 6 and 7 are lower than expected. In this event, all subsequent trial time points will be adjusted accordingly.

***Admission***

The following procedures will occur at admission to the clinical trial unit:

1. Perform abbreviated physical examination.
2. Perform alcohol breath test.
3. Obtain 12-lead ECG.
4. Record vital signs.
5. Collect urine for:
    a. urine drug screen
    b. urinalysis
    c. urine β-hCG pregnancy test for WOCBP
6. Cannulate volunteers with an indwelling IV cannula.
7. Collect blood samples for:
    a. haematology
    b. biochemistry (including lipids and iron studies)
    c. immune response assessments at 0 h (pre AL administration)
    d. malaria 18S qPCR at 0 h (pre AL administration)
    e. pSTAT3 assay at 0 h (pre AL administration)
    f. PK at 0 h (pre AL administration)
8. Record malaria clinical score.
9. Check volunteer diary cards.
10. Record AEs and use of concomitant medications.
11. Volunteers may be asked to complete a questionnaire about their experience during the conduct of the clinical trial.

***IMP administration***

The following procedures will occur when volunteers are confined in the clinical trial unit:

1. Administer AL under direct observation at 0, 8, 24, 36, 48 and 60 h on Days 8, 9, and 10.
2. Administer Rux or placebo (according to randomisation schedule) **2 h after administration of AL** at 2, 10, 26, 38, 50 and 62 h on Days 8, 9, and 10.
3. Follow volunteers for at least 72 h to monitor safety and tolerability of the treatment and adequate clinical response.
4. Collect blood samples for:
    a. haematology and biochemistry daily during confinement [twice daily on Day 8, baseline (number 7 above) and after the second dose of Rux or placebo]
    b. immune response assessments at 24, 48 and 72 h on Days 9, 10 and 11 (Time-points for immune response assessments may change based on data analysis from previous cohorts at the Investigator's discretion (refer to Lab Manual for each cohort)).

c. malaria 18S qPCR at 4, **8**, 12, 16, **24**, **36**, **48**, **60** and 72 h on Days 8, 9, 10 and 11
d. pSTAT3 assay at 28 h on Day 9
e. PK at 1, **2**, 3, 4, 5, 6, **8**, 12, **24**, **36**, **48**, **60**, **62,** 66 and 72 h on Days 8, 9, 10 and 11.
   - When PK and PD (immune response assessments, malaria 18S qPCR, pSTAT3 assay) sampling timepoints coincide with IMP administration **(in bold)** PK and PD sampling **MUST** occur **prior** to IMP administration.

5. Perform symptom-directed physical examination if clinically indicated at the Investigator's discretion.
6. Record vital signs 3 times a day while confined.
7. Obtain 12-lead ECG once daily while confined.
8. Record malaria clinical score 3 times a day while confined.
9. Record AEs and use of concomitant medications.

***Discharge***

Volunteers will be allowed to leave the clinical trial unit at the earliest 72 h after AL dosing at the Investigator's discretion. Before exit from confinement, the clinical trial unit nursing staff must advise the Investigator if any volunteer requires symptom-directed physical examination prior to discharge. Volunteers may be asked to complete a questionnaire about their experience during confinement of the clinical trial.

The following procedures will occur prior to discharge from the clinical trial unit:

1. Perform symptom-directed physical examination if clinically indicated at the Investigator's discretion.
2. Obtain 12-lead ECG.
3. Record vital signs.
4. Record malaria clinical score.
5. Record AEs and use of concomitant medications.
6. Collect blood samples for:
   a. haematology
   b. biochemistry
   c. immune response assessments

###### 1.3.1.7 OUTPATIENT MONITORING: DAY 11PM TO DAY 28

Out patient follow-up visits will occur during Days 11pm-28. Follow-up at either am (approximately 08.00) or am and pm (if necessary, approximately 12 h apart). Volunteers are required to fast for ≥8 h prior to visit Days 15 and 28.

The following procedures will take place during these visits:

1. Perform symptom-directed physical examination.
2. Record vital signs.
3. Record malaria clinical score if vital signs are abnormal.
4. Check volunteer diary cards.

5. Record AEs and use of concomitant medications.

**Safety samples**

6. Haematology Days 15, 22 and 28.
7. Biochemistry Days 15, 22 and 28 (including lipids and iron studies on Days 15 and 28).
8. RBC alloantibodies Day 28.
9. Urinalysis Day 28.

**PD blood collection**

10. If a minimum of one negative 18S qPCR is not detected by exit from confinement, then collect blood for malaria 18S qPCR at a minimum 3 times per week until a minimum of one negative 18S qPCR is detected.
11. Collect blood for 18S qPCR on Day 28.
12. Collect blood for parasite lifecycle stage qRT-PCR (if required).
13. Collect blood for pSTAT3 assay on Day 13.
14. Collect blood for immune response assessments on Days 13, 15 and 28.

**PK blood collection**

15. Collect blood for PK on Days 11 pm, 15, 18, 22 and 28.

###### 1.3.1.8 MONITORING VIA PHONE: DAY 28 TO DAY 85±7

During this period, clinical trial unit staff will contact volunteers every two weeks via a phone call or text message to monitor volunteer wellbeing. Volunteers will be given a diary and asked to record any signs or symptoms they experience and any concomitant medications they take.

##### 1.3.2 SECOND INOCULATION

Contingent upon SDRT decision, volunteers enrolled who remain eligible and do not demonstrate allo-immunisation will be intravenously administered a second (homologous) inoculation of ~2,800 viable parasites of *P. falciparum*-infected human erythrocytes.

###### 1.3.2.1 CONTINUED ELIGIBILITY VISIT: DAY 85±7

For the continued eligibility visit, volunteers will be asked to come to the clinical trial unit after an overnight fast of ≥8 h.

The continued eligibility visit will include all activities listed below:

1. Ask volunteer if they have any new AEs and/or use of medications since last involvement in trial, and review volunteer’s diary card.
2. Ask volunteer about recreational drug, alcohol, and tobacco use since last involvement in trial.
3. Record body weight.
4. Perform alcohol breath test.
5. Perform urine drug screen.
6. Perform full physical examination.

7. Ask volunteer to complete the Beck Depression Inventory.
8. Record vital signs (supine and standing blood pressure and heart rate).
9. Obtain a 12-lead ECG.
10. Collect urine for urinalysis.
11. Collect blood samples for
    a. haematology
    b. biochemistry (including lipids and iron studies)
    c. serology (viral hepatitis B and C, HIV)
    d. RBC alloantibodies testing
    e. serum β-hCG pregnancy test for WOCBP
12. Assess the cardiovascular disease risk as per exclusion criterion
13. Verify volunteer meets eligibility criteria.
14. Re-educate volunteers of signs and symptoms of malaria.

###### 1.3.2.2 SECOND MALARIA INOCULATION: DAY 90±7

Each volunteer will report to the clinical trial unit on the morning of Day 90±7. Depending on the number of volunteers that remain eligible for the second malaria inoculation, up to 4 inoculation days may be required.

The Investigator will review the volunteers' screening and eligibility confirmation visit results prior to their subsequent inoculation. The Investigator will emphasise the requirement to return to the clinical trial unit for malaria monitoring and antimalarial treatment after the inoculation.

On admission to the clinical trial unit, volunteers will be required to undertake further procedures to determine whether they remain eligible for repeat inoculation.

<u>The procedures that will be undertaken prior to inoculation on Day 90±7 include:</u>

1. Verify that all applicable eligibility criteria have been met.
2. Elicit information regarding any new medical conditions, illnesses and medication use since screening.
3. Perform alcohol breath test and urine drug screen.
4. Perform urine β-hCG pregnancy test for WOCBP.
5. Conduct abbreviated physical examination.
6. Record vital signs.
7. Obtain 12-lead ECG.
8. Cannulate volunteers with an indwelling IV cannula for the malaria challenge agent, and record which arm is used.
9. Collect blood samples for
    a. malaria 18S qPCR (baseline parasitaemia sample for second inoculation)
    b. immune response assessments (within 3 h prior to inoculation)
    c. safety serum retention sample

Administration of the malaria challenge agent:

1. Administer the malaria challenge agent of ~2,800 viable *P. falciparum* 3D7 infected human RBCs intravenously in the morning.
2. Observe volunteer for a minimum of 60 min after inoculation to evaluate for immediate adverse reactions.
3. Re-educate volunteers of the signs and symptoms of malaria.
4. Emphasise to volunteers the importance of returning for antimalarial monitoring and antimalarial treatment when directed.
5. Ensure volunteers still have diary cards and thermometers to record any temperature readings during the trial in the event of symptoms of fever. Volunteers will also record symptoms and concomitant medications on the diary cards during the trial.
6. Record vital signs prior to leaving the clinical trial unit (approximately 60 min after inoculation).
7. Record malaria clinical score prior to leaving the clinical trial unit (malaria clinical score baseline of second inoculation).
8. Record AEs and concomitant medications.

###### 1.3.2.3 MALARIA MONITORING VIA PHONE: DAYS 91±7 TO 93±7

During this period, the volunteer is expected to be asymptomatic. Clinical trial unit staff will contact volunteer via a daily phone call or text message to monitor volunteer wellbeing and to solicit any AEs and concomitant medications.

###### 1.3.2.4 MALARIA MONITORING: DAY 94±7 TO AL ADMINISTRATION: DAY TO BE INDIVIDUALISED, BASED ON TREATMENT CRITERIA

Volunteers will come to the clinical trial unit once daily from Day 94±7 until presence of asexual parasites is established by malaria 18S qPCR. Once malaria 18S qPCR becomes positive, and until antimalarial drug (AL) treatment, volunteers will come to the clinical trial unit up to twice-daily, separated by approximately 12 h.

The following procedures will occur during these visits:

1. Perform symptom-directed physical examination if clinically indicated at the Investigator's discretion.
2. Record vital signs.
3. Collect blood samples for:
    a. malaria 18S qPCR
    b. immune response assessments approximately three times per week until antimalarial (AL) treatment
    c. haematology and biochemistry at Investigator discretion
4. Record malaria clinical score.
5. Check volunteer diary cards.
6. Record AEs and use of concomitant medications.

###### 1.3.2.5 AL ADMINISTRATION: DAY TO BE INDIVIDUALISED, BASED ON TREATMENT CRITERIA

Volunteers will be administered AL at the clinical trial unit on an individualised basis, when:

- qPCR parasitaemia reaches ≥50,000 parasites/mL, or
- they have a malaria clinical score >6 and presence of parasitaemia, or
- they experience an SAE related to the malaria challenge agent, or
- they have a CTCAE grade 3 AE deemed related to malaria and not self-resolved or relieved with concomitant medications, or
- at Investigator discretion
- **If none the above criteria for AL administration are reached by Day 118±7 (28 days post second inoculation) then compulsory AL administration must occur.**

The following procedures will occur at the clinical trial unit at time of AL administration:

1. Perform abbreviated physical examination.
2. Obtain 12-lead ECG.
3. Record vital signs.
4. Collect blood samples for:
    a. haematology
    b. biochemistry (including iron studies),
    c. immune response assessments (within 3 h prior to AL administration)
    d. malaria 18S qPCR (within 3 h prior to AL administration)
5. Record malaria clinical score.
6. Record AEs and use of concomitant medications.

***Antimalarial treatment***

Administer AL as instructed by the manufacturer under direct observation at 0 h. Volunteers will not be confined, but will be followed up via a daily phone call or text message for the following 2 days to ensure adherence to AL twice daily dosing regimen.

###### 1.3.2.6 POST AL ADMINISTRATION: DAYS AL+3±2, AL+7±3, AL+20±3

For all volunteers a follow-up visit will occur 3 days (AL+3±2), 7 (AL+7±3) and 20 (AL+20±3) after initiation of AL. The following procedures will be performed at each visit, except where otherwise specified below:

1. Perform symptom-directed physical examination.
2. Record vital signs.
3. Record malaria clinical score.
4. Record AEs and use of concomitant medications
5. Collect blood samples for:
    a. haematology
    b. biochemistry (including iron studies [Days AL+7±3 & AL+20±3])

c. immune response assessments (Days AL+3±2, AL+7±3, & AL+20±3). Time-points for immune response assessments may change based on data analysis from previous cohorts at the Investigator's discretion (refer to Lab Manual for each cohort).
d. malaria 18S qPCR (Days AL+3±2 & AL+7±3 or until two consecutive negative malaria 18S qPCR results are obtained, or at Investigator discretion)
e. malaria parasite lifecycle stage qRT-PCR (if required, at Investigators decision)

###### 1.3.2.7 EOS: DAY AL+28±3 OR EARLY TERMINATION

For the EOS (or early termination) visit, volunteers will be asked to come to the clinical trial unit after an overnight fast of ≥8 h. The following procedures will be performed:

1. Full physical examination.
2. Record vital signs.
3. Record malaria clinical score.
4. Collect urine for urinalysis.
5. Perform urine β-hCG pregnancy test for WOCBP
6. Obtain 12-lead ECG.
7. Record AEs and use of concomitant medications.
8. Collect blood samples for:
    a. haematology
    b. biochemistry (including iron studies)
    c. serology
    d. immune response assessments. Time-points for immune response assessments may change based on data analysis from previous cohorts at the Investigator's discretion (refer to Lab Manual for each cohort).
    e. malaria 18S qPCR
    f. RBC alloantibodies
    g. safety serum retention sample
9. Volunteers may be asked to complete a questionnaire about their experience during the conduct of the clinical trial.

##### 1.3.3 ADDITIONAL BLOOD SAMPLING: 3, 6 & 12 MONTHS POST SECOND INOCULATION VISIT:

If consented separately (optional consent form) volunteers will be contacted at approximately 3, 6 and 12 months after their second inoculation visit and asked to return for blood collection for immune response assessments.

###### 1.3.3.1 EARLY TERMINATION VISIT

**Volunteers are informed of the essential requirement to complete the antimalarial drug treatment for their safety, via the Participant Information Sheet**. If withdrawal occurs at any stage of the trial, the volunteer will be asked to complete an EOS visit evaluation. Participation in an early termination evaluation by each volunteer is voluntary. Procedures during the early termination visit are the same as for the Day AL+28±3.

###### 1.3.3.2 UNSCHEDULED VISITS

Unscheduled visits for safety monitoring or malaria 18S qPCR may be required at the Investigator's discretion based on laboratory results, parasitaemia, or clinical symptoms. The volunteer will be contacted to arrange these visits. Where possible, visits will be arranged at a time that is both convenient for the volunteer and meets any clinical urgency as determined by the Investigator. Unscheduled visits will be documented in the source documents and eCRF.

#### 1.4 SCHEDULE OF ACTIVITIES TABLES

###### 1.4.1.1 SCHEDULE OF ACTIVITIES TABLES

The two Schedule of Activities Tables (first inoculation and second inoculation) summarise the activities and procedures to be conducted as per this protocol during screening, confinement, and post-confinement. Section 6 provides detailed information about the procedures.

##### 1.4.1.2 SCHEDULE OF ACTIVITIES TABLE — FIRST INOCULATION

Table indicative only. Refer to Section 1.3.1 for specific details of trial assessments.

| **Activity/Procedure** | **Screening D-28 to D-1** | **Eligibility visit D-3 to D-1** | **Malaria inoculation (first) D0** | **Post-inoculation phone contact D1 to D3** | **Malaria monitoring D4 to D7** | **AL and Rux treatment and clinical trial unit confinement D8 to D11** | **Out patient monitoring D11PM to D28** | **Phone contact D29 to D85±7** |
|---|---|---|---|---|---|---|---|---|
| Overnight fast of ≥8 h required for visit | X | | | | | X (admission) | X (D15 & D28) | |
| Informed consent & BDI | X | | | | | | | |
| Medical history, eligibility & prior medications | X | | X | | | | | |
| Drug screen & alcohol test | X | | X | | | X (admission) | | |
| Full physical examination | X | | | | | | | |
| Abbreviated physical examination | | | X | | | X (admission) | | |
| Symptom-directed physical examination | | | | | X | X (if clinically indicated during confinement & prior to discharge) | X (if clinically indicated) | |
| Vital sign assessment | X | | X (pre & post malaria inoculation) | | X | X (admission, discharge & 3 times daily while confined) | X | |
| ECG | X | | X | | | X (admission, discharge & once daily while confined) | | |
| Urinalysis | X | X | | | | X (admission) | X (D28) | |
| Intravenous cannulation | | | X | | | X | | |
| Haematology | X | X | | | | X | X (D15, 22 & D28) | |
| G6PD deficiency | X | | | | | | | |

| **Activity/Procedure** | **Screening D-28 to D-1** | **Eligibility visit D-3 to D-1** | **Malaria inoculation (first) D0** | **Post-inoculation phone contact D1 to D3** | **Malaria monitoring D4 to D7** | **AL and Rux treatment and clinical trial unit confinement D8 to D11** | **Out patient monitoring D11PM to D28** | **Phone contact D29 to D85±7** |
|---|---|---|---|---|---|---|---|---|
| Biochemistry (may include CRP, serum β-hCG, FSH [female, post-menopausal volunteers only], iron studies and lipids) | X | X | | | | X | X (D15, D22 & D28 | |
| Coagulation profile | X | | | | | | | |
| Serology (viral hepatitis B & C, HIV) | X | | | | | | | |
| QuantiFERON-TB Gold assay | X | | | | | | | |
| Urine β-hCG pregnancy test (WOCBP only) | | X | X | | | X (admission) | | |
| RBC alloantibodies | X | | | | | | X (D28) | |
| Safety serum retention sample | | | X | | | | | |
| AEs & concomitant medications | | | X (post malaria inoculation) | X | X | X (admission, discharge & while confined) | X | X |
| Malaria clinical score | | | X (post malaria inoculation) | | X | X (admission, discharge & 3 times daily while confined) | X | |
| Malaria 18S qPCR blood sampling | | | X | | X | X (times specified in text) | X (times specified in text) | |
| Parasite lifecycle stage qRT-PCR blood sampling | | | | | | | X (at Investigators discretion based on evidence of gametocytes) | |
| Rux and AL concentration blood sampling | | | | | | X (times specified in text) | X (D11 pm, D15, D18, D22 & D28) | |

| Activity/Procedure | Screening D-28 to D-1 | Eligibility visit D-3 to D-1 | Malaria inoculation (first) D0 | Post-inoculation phone contact D1 to D3 | Malaria monitoring D4 to D7 | AL and Rux treatment and clinical trial unit confinement D8 to D11 | Out patient monitoring D11PM to D28 | Phone contact D29 to D85±7 |
|---|---|---|---|---|---|---|---|---|
| pSTAT3 assay | | | X | | | X (0, 28 h on D8 & D9) | X (D13) | |
| Immune response assessments[a] | | | X | | | X (0, 24, 48 & 72 h on D8, D9, D10 & D11) | X (D13, D15 & D28) | |
| AL and Rux/placebo administration | | | | | | X (times specified in text) | | |
| Primacin™ treatment (if gametocytaemic as determined by qRT-PCR) | | | | | | | X | |

AE: adverse event; AL: artemether-lumefantrine; BDI: Beck Depression Inventory; D: Day relative to inoculation on Day 0; ECG: electrocardiogram; G6PD: glucose-6-phosphate dehydrogenase; PK: pharmacokinetic; TB: tuberculosis; WOCBP: women of childbearing potential.

[a] Time-points for immune response assessments may change based on data analysis from previous cohorts at the Investigator's discretion (refer to Lab Manual for each cohort).

##### 1.4.1.3 SCHEDULE OF ACTIVITIES TABLE — SECOND INOCULATION

Table indicative only. Refer to Section 1.3.2 for specific details of trial assessments.

| Activity/Procedure | Continued eligibility visit D85±7 | Malaria inoculation (second) D90±7 | Post-inoculation phone contact D91±7 to D93±7 | Outpatient monitoring D94±7 to parasitaemia threshold reached (50,000 parasites/mL) | AL treatment D-unknown (based on parasitaemia threshold) | Post AL administration D-AL+3±2, AL+7±3, AL+20±3 | EOS visit D-AL+28±3 |
|---|---|---|---|---|---|---|---|
| Overnight fast of ≥8 h required for visit | X | | | | | | X |
| Informed consent & BDI | X | | | | | | |
| Medical history, eligibility & prior medications | X | X | | | | | |
| Drug screen & alcohol test | X | X | | | | | |
| Full physical examination | X | | | | | | X |
| Abbreviated physical examination | | X | | | X | | |
| Symptom-directed physical examination | | | | X | | X | |
| Vital sign assessment | X | X (pre & post malaria inoculation) | | X | X | X | X |
| ECG | X | X | | | X | | X |
| Urinalysis | X | | | | | | X |
| Intravenous cannulation | | X | | | | | |
| Haematology | X | | | | X | X | X |
| Biochemistry (may include CRP, serum β-hCG [WOCBP only], FSH [female volunteers only], iron studies and lipids) | X | | | | X | X | X |
| Serology (viral hepatitis B & C & HIV) | X | | | | | | X |

| Activity/Procedure | Continued eligibility visit D85±7 | Malaria inoculation (second) D90±7 | Post-inoculation phone contact D91±7 to D93±7 | Outpatient monitoring D94±7 to parasitaemia threshold reached (50,000 parasites/mL) | AL treatment D-unknown (based on parasitaemia threshold) | Post AL administration D-AL+3±2, AL+7±3, AL+20±3 | EOS visit D-AL+28±3 |
|---|---|---|---|---|---|---|---|
| Urine β-hCG pregnancy test [WOCBP only] | | X | | | | | X |
| RBC alloantibodies | X | | | | | | X |
| Safety serum retention sample | | X | | | | | X |
| AEs & concomitant medications | | X (post malaria inoculation) | X | X | X | X | X |
| Malaria clinical score | | X (post malaria inoculation) | | X | X | X | X |
| Malaria 18S qPCR blood sampling | | X | | X (once daily until parasitaemia positive. Then up to twice daily until AL treatment) | X (within 3 h prior to AL treatment) | X (Days AL+3±2 & AL+7±3 or until two consecutive negative malaria 18S qPCR results are obtained, or at investigators discretion) | X |
| Parasite lifecycle stage qRT-PCR blood sampling | | | | | | X (at Investigators discretion based on evidence of gametocytes) | |
| Immune response assessments[a] | | X (within 3 h prior to inoculation) | | X (3x per week until antimalarial AL treatment) | X (within 3 h prior to AL treatment) | X (Days AL+3±2, AL+7±3 & AL+20±3) | X |
| Primacin™ treatment (if gametocytaemic as determined by qRT-PCR) | | | | | | X | |

AE: adverse event; AL: artemether-lumefantrine; BDI: Beck Depression Inventory; D: Day relative to first inoculation on Day 0; ECG: electrocardiogram; PK: pharmacokinetic; WOCBP: women of childbearing potential.

[a] Time-points for immune response assessments may change based on data analysis from previous cohorts at the Investigator's discretion (refer to Lab Manual for each cohort).

### 2 INTRODUCTION

#### 2.1 TRIAL RATIONALE

The World Health Organization (WHO) estimated 228 million cases of malaria and 405,000 deaths from malaria occurred worldwide in 2018. The WHO African Region carries the highest burden, with 213 million cases (93%). Nearly all of the cases (99.7%) in sub-Saharan Africa were caused by the parasite *Plasmodium falciparum* [1]. Current approaches to malaria control are failing, and emerging evidence suggests that both developing and established immunoregulatory pathways impede the acquisition of natural, drug-mediated, or vaccine-induced immunity against malaria. Type I interferons have been identified as important immune regulators in malaria, and are promising targets for improving anti-parasitic immunity.

We have identified a registered drug — Jakavi® (ruxolitinib) — that has shown immune-enhancing ability when combined with a drug targeted at the pathogen of interest. We wish to test whether co-administration of ruxolitinib and the antimalarial drug Riamet® (artemether-lumefantrine; AL) can enhance immune response in malaria-naïve humans who have an induced *P. falciparum* infection.

This trial is designed to assess the safety, tolerability, PK, PD (malaria 18S qPCR, pSTAT3, and immune responses) of AL+Rux in healthy adults with *P. falciparum* induced blood stage malaria (IBSM). This will be a randomised placebo-controlled trial that uses the IBSM model: healthy volunteers will be inoculated with blood stage *P. falciparum* parasites and randomised to receive co-administered AL and Rux or AL and placebo. Volunteers will be re-inoculated (with a homologous infection) 90±7 days after their first inoculation. Safety, tolerability, PK, antimalarial drug activity, and immune response will be assessed.

Volunteers, if they agree will be contacted approximately 3, 6 and 12 months after their EOS visit to provide blood samples to investigate anti-parasitic immune response longevity.

#### 2.2 BACKGROUND

Malaria remains a significant health problem in tropical and sub-tropical regions of the world. Substantial progress in disease control has been made through vector management, use of insecticide-treated bed nets, and improved diagnosis and drug treatment [1]. However, malaria remains responsible for more deaths than other parasitic diseases (WHO website - https://www.who.int/). Critically, there is no licensed vaccine, and the emergence of artemisinin-resistant parasites in the Greater Mekong Subregion represents a serious threat to malaria treatment and control efforts. A major impediment to malaria eradication is our poor understanding of host immunity against the *Plasmodium* species that cause malaria in humans. This shortcoming in knowledge is especially evident given the poor performances of malaria vaccine candidates in malaria endemic areas and a lack of consensus on how to generate long-lasting immunity in such endemic settings.

Studies performed more than 50 years ago showed anti-parasitic antibodies can protect against clinical malaria and control parasite growth [2]. However, these responses take years to develop in malaria endemic areas, after repeated exposure to malaria parasites [3]. Recent reports also highlight a requirement for antibodies with specific functional properties to mediate immunity [4]. The development of long-lived, anti-parasitic memory B and plasma cells that produce these antibodies requires $CD4^+$ T cell help [5, 6]. Specialised $CD4^+$ T follicular helper (Tfh) cells are critical in this process by providing key cognate and soluble signals to B cells [7, 8]. Interferon gamma (IFNγ) production by $Tbet^+$ $CD4^+$ T (Th1) cells is also important for generating specific antibody isotypes and activating phagocytic cells for increased antibody-mediated and complement-mediated removal of parasitised red blood cells and generating microbiocidal molecules, such as reactive oxygen and nitrogen intermediates [9, 10]. However, these anti-parasitic responses are often absent, sub-optimal, or dysfunctional in individuals living in malaria endemic areas [11-13]. The reasons are still unclear, but emerging evidence indicates parasite-induced development of atypical B cells, sub-optimal Tfh cells, and Th1 cell responses characterised by autologous interleukin 10 (IL-10) production (type 1 regulatory [Tr1] cell responses), as well as the development of immunoregulatory networks [14-19], all contribute to these outcomes. Unless these factors are addressed, malaria eradication will not be possible.

#### 2.3 RISK/BENEFIT ASSESSMENT

##### 2.3.1 KNOWN POTENTIAL RISKS

We identified potential risks by reviewing clinical studies conducted to date that used the IBSM model, CMI sheets for Riamet® (AL) and Jakavi® (Rux), the Investigator's Brochure for the 3D7 *P. falciparum* challenge agent [20], and published literature.

**IBSM model risks**

Risks pertaining to the IBSM model are development of blood borne infections, reaction to the blood sample, severity of malaria infection, development of liver function abnormalities, and occurrence of cardiac adverse events (AEs).

**Risk management of blood borne infections**

The *P. falciparum* 3D7 challenge agent will be used to induce blood stage malaria in this trial. Although the challenge agent contains a very small amount of blood, risk of a transfusion-transmissible infection in this trial is extremely low. Firstly, donors were screened and tested negative for the presence of active blood borne infections. Secondly, the Australian Red Cross Blood Service (Blood Service) removed white blood cells from the donor blood to lower the risk of a transfusion-transmissible infection. Thirdly, the volume of blood used in the IBSM model for transmitting malaria is many thousands of times smaller than in a transfused unit (i.e., a relatively lower risk of infection). As part of the safety monitoring, all volunteers will have serum stored for testing of blood borne infections before and after the trial (Section 1.3). To date, no blood borne infections have been reported in any of the 397 volunteers who have received this *P. falciparum* 3D7 challenge agent in IBSM clinical trials.

**Risk management of reaction to the blood sample**

The risk of developing red blood cell (RBC) alloantibodies and/or experiencing an acute haemolytic reaction in this trial is considered extremely low because the donor blood used to produce the challenge agent was blood group O (RhD) negative. People with this blood group are generally considered "universal donors". However, it is possible that volunteers could suffer a transfusion reaction after they receive the challenge agent, or could develop alloantibodies to the donor RBCs that may make blood transfusion more difficult in the future. To date, one volunteer has developed an antibody response to a minor Rh antigen (anti-E antibody) following inoculation with *P. falciparum* 3D7 [20]. However, there was no laboratory evidence to indicate that the specific Rh phenotype of the donor RBCs in the challenge agent stimulated production of this anti-E alloantibody.

Volunteers will be monitored for signs and symptoms in the period immediately after administration of the challenge agent to further assess the risk of the challenge agent causing a transfusion reaction. All volunteers will be tested for RBC alloantibodies at screening, on Day 29, at the continued eligibility visit before the second inoculation on Day 85±7, and at the end of the trial, as part of safety monitoring (Section 1.3).

Women of childbearing potential (WOCBP) have a small additional risk of developing RBC alloantibodies, which could cause problems during pregnancy. WOCBP who have participated in several IBSM trials with *P. falciparum* isolate 3D7 have had no known issues to date. Including WOCBP in the trial enhances the generalisability of the trial results.

**Risk management of severity of malaria infection**

The number of viable blood stage parasites that will be used to infect the volunteers in this trial (~2,800) is substantially lower than the parasitaemia induced from the bite of a single malaria-infected mosquito (~30,000 parasites are released into the blood when they break out of a single infected liver cell [21]. In this trial, parasite growth and malaria symptoms will be closely monitored in volunteers following administration of the challenge agent. The threshold for commencement of antimalarial drug treatment defined for Day 8 has been selected because it is before the time-point at which clinical symptoms of malaria are likely to occur.

Volunteers will be admitted to the clinical trial unit for earlier treatment if they experience an SAE, or if they have a CTCAE grade 3 AE deemed related to malaria and not self-resolved or relieved with concomitant medications, or at Investigators discretion.

For the second inoculation, volunteers will be administered AL at the clinical trial unit on an individualised basis, when:

- qPCR parasitaemia reaches ≥50,000 parasites/mL, or
- they have a malaria clinical score >6 (appendix 3), and presence of parasitaemia,
- they experience an SAE related to the malaria challenge agent, or
- they have a CTCAE grade 3 AE deemed possibly related to malaria and not self-resolved or relieved with concomitant medications, or
- at Investigator discretion.

If none of the above criteria for AL administration are reached by Day 118±7, then compulsory AL administration must occur.

**Risk management of liver function abnormalities**

Transient, asymptomatic liver function test (LFT) abnormalities, including rare cases of alanine aminotransferase (ALT) and/or aspartate aminotransferase (AST) elevations >10 fold the upper limit of normal (×ULN), have been reported in several volunteers in IBSM studies [20, 22]. However, no changes in bilirubin were reported except in one volunteer with unappreciated pre-existing liver disease [20, 22]. The LFT abnormalities did not require treatment, and resolved by the end of the studies. A few cases of the LFT elevations were considered serious AEs (SAEs) by two Pharma sponsors due to internal processes for SAE notifications. An independent review involving drug-induced liver injury experts found these liver function abnormalities are most likely a direct consequence of the malaria infection rather than a direct drug-induced liver injury caused by an investigational antimalarial drug. As a precaution, all volunteers in this trial will undergo regular safety monitoring to assess for asymptomatic liver function abnormalities. Volunteers will be required to minimise intake of possibly hepatotoxic substances, such as alcohol and paracetamol, during the trial (see Section 3.2). Drugs of abuse are not permitted under any circumstance.

**Risk management of cardiac adverse events**

To our knowledge, three cardiac SAEs have been reported in healthy volunteers in the Netherlands participating in malaria challenge studies using sporozoites (i.e., direct feeds by infected mosquitoes rather than IBSM infection). No cardiac SAEs caused by the challenge agents have been reported in IBSM studies. However, in a recent trial, 2 volunteers (1 infected with the *P. falciparum* 3D7 challenge agent and 1 infected with another malaria challenge agent strain *P. falciparum* K13) developed ventricular extra systoles that were classified as moderate AEs possibly related to malaria. As a precaution, people at significant risk of cardiovascular disease will be excluded from participating in IBSM studies, and regular safety monitoring, including physical examination and ECG recordings, will take place for all volunteers. Follow-up with a cardiologist is also available if any cardiac AEs are seen during the IBSM studies.

#### Risk management of trial interventions

Individual risks pertaining to Riamet® and Jakavi® have been well researched, and we have conducted modelling simulations to anticipate drug-drug interactions. We have also developed trial-specific toxicity rules related to the IMPs and malaria challenge agent.

**Risk management of using artemether-lumefantrine**

Riamet® (artemether-lumefantrine; AL) is a registered drug for the treatment of malaria. The dose of AL that will be used in this trial is the standard adult dose. Detailed information about Riamet® is available in the CMI sheet.

**Risk management of using ruxolitinib and placebo**

Jakavi® (ruxolitinib; Rux) is a registered drug that has undergone extensive safety testing. The dosing regimen of Rux that will be used in this trial is 6 doses of 1 tablet (1 × 20 mg) administered orally twice daily over 3 consecutive days. Rux is registered for the treatment of intermediate or high-risk myelofibrosis in adults, and has also been safely and effectively used in children with type I interferonopathy [23].

The safety, tolerability, and PK of Rux has been extensively studied in healthy volunteers including males and females (including WOCBP) [24]. The oral dose PK, PD, safety, and tolerability of Rux were evaluated in healthy volunteers in 2 double-blind, randomised, and placebo-controlled studies conducted in the United States. The first study enrolled 23 volunteers and evaluated single ascending doses of 5 to 200 mg and the effect of food. From a safety perspective, 1 volunteer withdrew consent, 1 volunteer was lost to follow-up, and 3 volunteers withdrew prematurely because of AEs, including the 1 case of a serious adverse event of hyponatremia following dosing with 5 mg Rux in a volunteer with a history of low sodium levels, judged unrelated to study drug; 1 case of nonsustained ventricular tachycardia following placebo dosing; and 1 case of nonsustained ventricular tachycardia following dosing with 5 mg. All 5 volunteers were replaced. There was no dose dependency for the frequency of AEs as a whole or of any particular AE. The most frequent AEs were headache (13.0% overall incidence) occurring in volunteers receiving placebo, 5 mg, or 100 mg; diarrhoea (13.0% overall incidence), occurring in volunteers receiving placebo, 5 mg Rux, or 100 mg Rux; and blood sampling catheter site haemorrhage (13.0% overall incidence) occurring in volunteers receiving placebo, 50 mg Rux, or 100 mg Rux. The second study enrolled 71 volunteers and evaluated multiple ascending doses, including both once- and twice-daily dosing for 10 days. In this study, AEs at least possibly related to study medication and occurring in more than 1 Rux-treated volunteer were restricted to neutropenia, which occurred in 3 volunteers receiving 50 mg bid (2 were of grade 2 severity and 1 was grade 4). Three volunteers discontinued the study: 1 volunteer receiving 50 mg b.i.d was discontinued from the study because of grade 4 neutropenia. Another volunteer, who received placebo, was discontinued for mild rhabdomyolysis, assessed by the blinded investigator as probably related to study medication. A third volunteer withdrew consent after 4 days of dosing with 50 mg b.i.d INCB018424. Neutropenia of any severity grade was observed in 22.2% of placebo volunteers, 11.1% of volunteers at 50 mg qd, 66.6% of participants at 100 mg qd, 12.5% of participants at 15 mg b.i.d, 33.3% of volunteers at 25 mg bid, and 66.6% of volunteers at 50 mg b.i.d. These were generally transient and rapidly normalised following the last dose of study medication. In both the single- and multiple-dose studies, AEs were, in general, mild to moderate in intensity and resolved quickly. The 1 episode of grade 4 neutropenia in the volunteer who received 50 mg bid was considered a dose-limiting toxicity. In summary, Rux was generally safe and well tolerated, with 25 mg b.i.d and 100 mg qd established as the maximum tolerated doses in healthy volunteers. The safety, tolerability, and PK of Rux has further been further evaluated in healthy Japanese volunteers [25]. Forty volunteers were randomised to receive single (10–100 mg) and multiple (10 and 25 mg every 12 h) doses of Rux or placebo. From a safety perspective, no dose discontinuations or SAEs occurred. Dose adjustments were made in two volunteers treated with b.i.d doses because of decreased neutrophil counts on day 5. Both volunteers had dose reductions from 10 and 25 mg b.i.d to 10 and 25 mg single dose, respectively, on days 6 and 7. Volunteers were dosed according to schedule (b.i.d on days 8 and 9, single dose on day 10) for the remainder of the dosing period. Most AEs were observed in volunteers during treatment with repeated b.id doses, during which the observation period was longer, but there was no clear pattern of dose dependency. AEs occurred in 15% of the volunteers, including placebo-treated volunteers; all AEs were grade 1 or 2 and were classified as being related to study drug. Grade 2 neutropenia occurred in five (12.5%) volunteers: one treated at 100 mg single dose, three treated at 10 mg b.i.d, and one treated at 25 mg b.i.d. A decrease in neutrophil count was also observed at some point during the study in all volunteers receiving placebo. For all volunteers, neutrophil counts returned to normal within 24 h of the last dose. In summary, orally administered Rux was well tolerated in healthy Japanese volunteers with no apparent differences in the safety or PK of Rux between Japanese and non-Japanese volunteers.

Rux has no known contraindications. Detailed information about Jakavi® is available in the CMI sheet.

The dosing regimen of the placebo that will be used in this trial is 6 doses of 1 tablet (1 × 20 mg) administered orally twice daily over 3 consecutive days. There are no anticipated risks associated with the administration of the placebo.

**Risk Management of using AL and Rux in combination**

In the context of evaluating the combination of two registered medications (AL and Rux) and malaria infection, the unit is prepared for potential SAEs and will closely monitor for these. Candidate volunteers with a risk of cardiovascular, neurological or psychiatric conditions or with hepatic/haematology abnormalities will be excluded from participation in the trial. More specifically, candidate volunteers with liver function test (LFT) results and/or an absolute neutrophil count (ANC) outside of the normal ranges, or other clinically significant haematology results, at screening or eligibility visits, will be excluded. Close clinical monitoring during the trial will include LFT measurements, haematology parameters with a focus on absolute neutrophil count (ANC), and ECG measurements. Volunteers will be confined in the clinical unit for at least 72 h post-first IMP administration and discharged only when clinically well. Furthermore, outpatient follow-up, which will include safety assessments, will occur until EOS visit.

There are toxicity rules that apply to the most significant risks of this trial.

**Toxicity rules**

When applying toxicity rules, AEs will be graded using the CTCAE grading system for standardised recording and reporting. For the purpose of this protocol, "toxicity" refers to the occurrence of any grade 3 or higher grade AE that is related to an IMP (AL, Rux, and Placebo) or the malaria inoculation.

**General toxicity rules:**

The general toxicity rules refer to all AEs, excluding those relating to the liver and haematology (other than neutropenia, which is an identified liability of Rux). There is a greater anticipation of liver and haematology AEs with malaria inoculation (mild/moderate ALT elevations, mild/moderate decrease in ANC, lymphocyte and platelet count) and the CTCAE grading system may not necessarily give clear enough guidance.

Therefore, trial-specific toxicity rules that apply to AEs relating to the liver and haematology have been created for this trial.

**Individual toxicity rules:**

- Rux administration for an individual volunteer will be stopped if the volunteer experiences any CTCAE grade 3 (or higher) AE, or an SAE irrespective of severity/CTCAE grade deemed related to IMP.

**Group toxicity rules:**

- Rux dosing will be suspended in all volunteers, or potentially only in an affected subgroup if either of the following occur: (1) One volunteer experiences a severe (CTCAE grade 3 or higher) AE deemed related to Rux, (2) One volunteer experiences an SAE (irrespective of severity/CTCAE grade) deemed related to Rux.

Dosing can only continue with an amendment to the protocol, which has been approved by the HREC.

**Individual toxicity rules:**

Dosing of Rux in a volunteer will be suspended if they experience any of the following:

LFT elevation

- ALT or AST value >3× ULN together with bilirubin increase >2× ULN.
- ALT or AST value >8× ULN.
- ALT or AST value >3× ULN, and symptomatic.

Haematology

- Haemoglobin decrease from baseline (eligibility visit) of 4g/dL (40g/L) or lower.
- Clinically significant drop in neutrophil count (grade 3 or higher).
- Platelet count drop of >50% from baseline or absolute value <100 000 $mm^3$ (Day -1 blood samples).

**Group toxicity rules:**

If one volunteer fulfils at least one of the individual toxicity rules, and this could be reasonably attributed to Rux, dosing for all volunteers in the trial will be at least temporarily suspended and an SDRT meeting will be arranged. At this meeting, a decision will be made regarding the continuation of dosing in the remaining volunteers.

If two or more volunteers fulfil any of the individual toxicity rules, then dosing will be suspended in all volunteers and can only continue if an SDRT meeting is arranged (and a decision made that dosing can continue) and an amendment is made to the protocol, which has been reviewed/approved by the HREC.

**Drug-drug interaction**

Results of physiologically based pharmacokinetic simulations conducted using Simcyp and available *in vitro* CYP inhibition data for Rux, indicate that the risk of drug-drug interaction from giving Rux (a CYP3A4 inhibitor) 2 h after administration of AL (both CYP3A4 substrates) is not expected to be clinically significant, and is lower than giving Rux before AL or concomitantly. Volunteers will be monitored and assessed in confinement for at least 72 h following administration of both drugs to ensure tolerance of the AL+Rux.

**Rescue medication risks**

Risks related to use of primaquine phosphate (Primacin®) and atovaquone/proguanil (Malarone®) are detailed in the prescribing information provided by manufacturers. Primacin® may cause severe haemolytic anaemia in volunteers with glucose-6-phosphate dehydrogenase (G6PD) deficiency. Volunteers will be tested for G6PD deficiency at screening to ensure the safety of Primacin®. Volunteer's G6PD status will determine how they are treated with Primacin®. A dosing table based on G6PD status will be provided in the Pharmacy Manual.

**Safety Data Review Team**

A SDRT will review predefined trial data to inform decision making on trial progression. The SDRT will assess if any of the individual or group toxicity rules are met and if the concerned AEs have a causal relationship with malaria infection, an IMP (AL, Rux, and Placebo) or rescue medication. If so, the SDRT will act in accordance with the predefined toxicity rules.

Any additional information, if required, will be included in the SDRT charter.

**General risk management**

This trial will be conducted at an early phase clinical trial unit by an experienced Principal Investigator and well trained medical and technical staff with ample experience in the conduct of human malaria challenges in healthy volunteers. The trial has been designed to include suitable volunteers and to monitor, treat and report potential expected adverse reactions as well as potential unexpected AEs. Additionally, a sentinel dosing strategy will be used.

In applying the above risk management strategies, the overall risk to volunteers in this trial is considered low.

The risk to volunteers in this trial will also be minimised as follows:

- Adherence to the inclusion/exclusion criteria to ensure only volunteers who are not at any perceived risk are enrolled.
- Close clinical and laboratory monitoring to ensure the safety and wellbeing of the volunteers
- The total volume of blood drawn from each volunteer enrolled into the trial will not exceed a standard unit of blood (approximately 470 mL) over any 30-day period.
- In the rare event that a volunteer requires hospitalisation at the request of the PI or representative, this will be done at the Royal Brisbane and Women's Hospital or Caboolture Hospital. Emergency procedures are in place at the clinical research unit for managing any unforeseen clinical emergencies that may arise.
- All volunteers will be prescribed curative therapy for malaria (Riamet® with the addition of Primacin™ if gametocytes are detected) for final parasite clearance during or at the end of the trial. Any volunteer inoculated with the *P. falciparum* 3D7 challenge agent who withdraws from the trial before two negative 18S malaria qPCR results can be obtained, will receive a compulsory course of Primacin® to ensure clearance of gametocytes.

##### 2.3.2 KNOWN POTENTIAL BENEFITS

There are no expected clinical benefits for the healthy volunteers who will participate in this trial. There may be benefits to others in the future if the results of this study lead to improved treatment outcomes in patients with malaria.

##### 2.3.3 ASSESSMENT OF POTENTIAL RISKS AND BENEFITS

On the basis of the safety provisions and risk management strategies outlined in Section 2.3.1, the overall risk to the volunteers in this trial is considered to be minimal and acceptable, and the potential of future improved treatment for malaria is considered to outweigh these potential risks.

#### 2.4 SCIENTIFIC RATIONALE FOR TRIAL DESIGN

Recently, type I interferons (IFNs) have emerged as important regulators of IL-10 production by Tr1 cells. Type I IFNs comprise a large family of cytokines that includes several types of IFNα and two types of IFNβ proteins which all signal through the common IFNα receptor (IFNAR) that consists of IFNAR1 and IFNAR2 chains which signal via STAT-1 and STAT-2 [26, 27] to mediate diverse functions during many infections [28]. Polymorphisms in the *IFNAR1* gene have been associated with increased risk of severe malaria in The Gambia [29, 30], while a whole-brain transcriptional analysis in genetically resistant and susceptible inbred mice infected with *P. berghei* ANKA identified type I IFN-dependent transcriptional program associated with the pathogenesis of severe malaria ECM [31]. Type I IFNs suppress $CD4^+$ T cell-dependent parasite control during experimental blood stage malaria by modulating the function of dendritic cells following *P. berghei* ANKA infection, rather than acting directly on Th1 cells [32, 33]. Another trial in mice infected with *P. yoelli* showed that type I IFNs directly promoted the expansion of Tr1 cells [22]. Significantly, in malaria volunteer infection studies (VIS), type I IFNs produced by several different cell sources were found to be important regulators of developing anti-parasitic immunity. Type I IFNs not only suppressed innate immune cell function and parasitic-specific $CD4^+$ T cell IFNγ production, but also promoted the development of parasitic-specific Tr1 cells [34]. Thus, type I IFNs are key immunomodulatory molecules in humans infected with *P. falciparum*, and targeting this pathway for clinical advantage is a promising strategy to overcome established or developing immunoregulatory networks to improve natural, drug-mediated, or vaccine-induced immunity against malaria.

We sought a drug that could block type 1 IFN signaling, and identified ruxolitinib (Jakavi®) as a suitable candidate for investigation. Ruxolitinib (Rux) is a licensed orally administered small molecule JAK1/2 inhibitor that has an established safety profile, including a relatively short elimination half-life (~3 h). The Janus kinase family of tyrosine kinases (JAKs) are intimately associated with cytokine receptors, including the type I IFN receptor. Following the binding of cytokines on these receptors, the JAKs become phosphorylated, and in turn phosphorylate the cytoplasmic domains of the cytokine receptor, creating binding sites for signal transducers and activators of transcription (STATs). The STATs are then phosphorylated by JAKs, resulting in the dissociation of STATs from the cytokine receptor and subsequent translocation to the cell nucleus where they drive transcription of target genes [35]. Following the binding of IL-6 to the IL-6 receptor, JAK1 and JAK2 are phosphorylated leading to the tyrosine phosphorylation of STAT3 (pSTAT3), and this robust response in monocytes and T cells is often used as a surrogate assay to measure the Rux PD effect (pSTAT3 inhibition) [36]. Rux is used to treat neoplastic diseases [37, 38], but has also been used to successfully treat children with a type I interferonopathy associated with gain of function mutations in *TMEM173* (encoding STING), responsible for high childhood morbidity and mortality [39]. In addition, Rux was found to reduce serum type I IFN levels and IFN-inducible gene scores in dermatomyositis patients [23]. Thus, there is a growing body of evidence that Rux may be used to block type I IFN signaling in humans in a range of diseases, including malaria.

Rux was also selected because of its potential for application in malaria endemic settings either in combination with anti-parasitic drugs and/or malaria vaccines. The short half-life of Rux will minimise risk of adverse outcomes in populations where co-infections with tuberculosis, human immunodeficiency virus (HIV) and other pathogens is relatively common.

#### 2.5 JUSTIFICATION FOR DOSE

The challenge agent dose of an estimated ~2,800 viable *P. falciparum* 3D7 parasite-infected RBCs in a volume of 2 mL was chosen based on our experience running IBSM studies using this challenge agent. The dose of AL will be the standard dosing regimen for adults. The 20 mg Rux twice daily dosing to be used in this study is the standard dose for patients with platelet count >200x$10^9$/L in the Australian Product Information (PI) for Jakavi®. A 3- day 20 mg b.i.d. dosing regimen was considered appropriate for healthy volunteers based on the reported safety of a higher dose of 25 mg b.i.d. over a 10-day period in healthy volunteers in a phase 1 safety trial. [24]. No effect of food was found in this trial, and, consequently, Rux will be administered without regard to meals. The relatively short duration of dosing (3 days) and our aim for an unequivocal assessment of immune boosting by Rux, rather than clinical endpoints, was balanced against minimising harm to volunteers and the chosen regimen was determined to be most suitable.

We consider co-administering AL and Rux to be appropriate because this is how Rux would likely be administered in field settings. The 3-day dosing of Rux in combination with AL is also supported by an immunological rationale for combining the therapies, which is thought to involve AL related parasite killing leading to consequent activation of type 1 IFN pathways, which would be disrupted by Rux. Review of healthy volunteer data emphasised that possible haematological effects (reduction in ANC) are monitorable, low grade, predictable in terms of timing and grade, directly linked to the effect of the drug. In particular, at the proposed regimen, a decrease in ANC is expected to be transient and due to margination rather than a direct toxic effect. Any impact of AL on Rux activity, including immune boosting, will be assessed by measuring pSTAT3 inhibition in blood samples taken from volunteers, as previously described in phase 1 safety studies [24].

#### 2.6 END OF TRIAL DEFINITION

A volunteer is considered to have completed the trial if they have completed all phases of the trial in which they are enrolled including the last visit or the last scheduled procedure shown in the Schedule of Activities (SoA), Section 1.4. The end of the trial is defined as completion of the last visit or procedure shown in the SoA in the trial globally.

### 3 TRIAL POPULATION

#### 3.1 INCLUSION CRITERIA

Individuals must fulfil all of the following criteria to be eligible for inclusion in this trial:

**Demography**

1. Male or female (non-pregnant, non-lactating) aged 18 to 55 years inclusive who will be contactable and available for the duration of the trial and up to two weeks following the EOS visit.
2. Total body weight greater than or equal to 50 kg, and a body mass index (BMI) within the range of 18 to 32 $kg/m^2$ (inclusive). BMI is an estimate of body weight adjusted for height. It is calculated by dividing the weight in kilograms by the square of the height in metres.

**Health status**

3. Certified as healthy by a comprehensive clinical assessment (detailed medical history and full physical examination).
4. Fully vaccinated (meaning first and second dose) against COVID-19 at least 14 days prior to IMP administration
5. Vital signs at screening (measured after 5 min in the supine position):
    - Systolic blood pressure (SBP) - 90–140 mmHg,
    - Diastolic blood pressure (DBP) - 40–90 mmHg,
    - Heart rate (HR) 40–100 bpm.
6. At screening, continued eligibility (Day 85±7), pre-inoculation (first and second) and pre IMP dosing: QTcF ≤450 msec (male volunteers); QTcF ≤470 msec (female volunteers); PR interval ≤210 msec for both males and females.
7. Heterosexual female volunteers of childbearing potential who have, or may have, male sexual partners during the course of the study should be using an insertable (implant or IUD), injectable, transdermal or combination oral contraceptive approved by the TGA combined with a barrier contraceptive from the time of informed consent until EOS. Abstinent female volunteers must agree to start a double method if they start a sexual relationship with a male during the trial. Female volunteers must not be planning *in vitro* fertilisation within the required contraception period.

    Women of non-childbearing potential who will not require contraception during the trial are defined as: surgically sterile (tubal ligation is not considered surgically sterile), post-menopausal (spontaneous amenorrhoea for ≥12 months, or spontaneous amenorrhoea for 6-12 months and follicle-stimulating hormone (FSH) ≥40 IU/mL; either should be together with the absence of oral contraceptive use for >12 months).

Male volunteers who have, or may have female sexual partners during the course of the study must agree to use a double method of contraception including condom plus diaphragm, or condom plus intrauterine device, or condom plus stable oral/transdermal/injectable hormonal contraceptive by the female partner, from the time of informed consent through to 90 days after the last dose of the IMP. Abstinent male volunteers must agree to start a double method if they begin a sexual relationship with a female during the trial, and through to 90 days after the last dose of the IMP. Male volunteers with female partners that are surgically sterile, or male volunteers who have undergone sterilisation and have had testing to confirm the success of the sterilisation, may also be included.

**Regulations**

8. Completion of the written informed consent process prior to undertaking any trial-related procedure.
9. Must be willing and able to communicate and participate in the whole trial.
10. Agreement to adhere to Lifestyle Considerations (see Section 3.3) throughout trial duration.

#### 3.2 EXCLUSION CRITERIA

Individuals fulfilling any of the following criteria are not eligible for inclusion in this trial:

**Medical history and clinical status**

1. Individual who lives alone, OR, does not satisfy the following criteria: Volunteers who live alone may be included on a case-by-case basis, following discussion with the Principal Investigator and with approval of the Medical Monitor. Volunteers who live alone must identify and provide contact details of a support person who is aware of the volunteer's participation in the study and is available to provide assistance if required (for example with contacting the volunteer in the event that study staff are unable to, or with transporting the volunteer to and from the study site if required). This criteria must be adhered to during the following time periods:
    i. First inoculation — at any stage from inoculation day until Day 15
    ii. Second inoculation — at any stage from inoculation Day until the end of the Riamet® treatment.
2. Known hypersensitivity to ruxolitinib, artesunate or any of its derivatives, artemether, lumefantrine or other artemisinin derivatives, proguanil/atovaquone, primaquine, or 4-aminoquinolines.
3. Haematology, biochemistry or urinalysis results at screening or at the eligibility visit, or at the Day 85±7 continued eligibility visit that are outside of Sponsor-approved clinically acceptable laboratory ranges, and are considered clinically significant by the Investigator or their delegate.
4. Participation in any investigational product trial within the 12 weeks preceding IMP administration.
5. Symptomatic postural hypotension at screening (confirmed on two consecutive readings), irrespective of the decrease in blood pressure, or asymptomatic postural hypotension defined as a

decrease in systolic blood pressure ≥20 mmHg within 2–3 min when changing from supine to standing position.

6. History or presence of diagnosed (by an allergist/immunologist) or treated (by a physician) food or known drug allergies (including but not limited to allergy to any of the antimalarial rescue medications), or any history of anaphylaxis or other severe allergic reactions including face, mouth, or throat swelling or any difficulty breathing. Volunteers with seasonal allergies/hay fever or allergy to animals or house dust mite that are untreated and asymptomatic at the time of dosing can be enrolled in the trial.
7. History of convulsion (including drug or vaccine-induced episodes). A medical history of a single febrile convulsion during childhood is not an exclusion criterion.
8. Presence of current or suspected serious chronic diseases such as cardiac or autoimmune disease, diabetes, progressive neurological disease, severe malnutrition, acute or progressive hepatic or renal disease, porphyria, psoriasis, rheumatoid arthritis, asthma (excluding childhood asthma, or mild asthma with preventative asthma medication required less than monthly), or epilepsy.
9. History of malignancy of any organ system (other than localised basal cell carcinoma of the skin or *in situ* cervical cancer), treated or untreated, within five years of screening, regardless of whether there is evidence of local recurrence or metastases.
10. Individuals with history of schizophrenia, bipolar disorder psychoses, disorders requiring lithium, attempted or planned suicide, or any other severe (disabling) chronic psychiatric diagnosis including generalised anxiety disorder.
11. Individuals who have been hospitalised within five years prior to enrolment for either a psychiatric illness or due to danger to self or others.
12. History of an episode of mild/moderate depression lasting more than 6 months that required pharmacological therapy and/or psychotherapy within the last 5 years; or any episode of major depression.

    The Beck Depression Inventory (BDI) will be used as a validated tool for the assessment of depression at screening. In addition to the conditions listed above, volunteers with a score of 20 or more on the BDI and/or a response of 1, 2 or 3 for item 9 of this inventory (related to suicidal ideation) will not be eligible for participation. These volunteers will be referred to a general practitioner or medical specialist as appropriate. Volunteers with a BDI score of 17 to 19 may be enrolled at the discretion of an Investigator if they do not have a history of the psychiatric conditions mentioned in this criterion and their mental state is not considered to pose additional risk to the health of the volunteer or to the execution of the trial and interpretation of the data gathered.
13. History of recurrent headache (e.g. tension-type, cluster, or migraine) with a frequency of ≥2 episodes per month on average and severe enough to require medical therapy, during the 2 years preceding screening.
14. Presence of clinically significant infectious disease or fever (e.g., sublingual temperature ≥38.5°C) within the five days prior to first and second inoculation.

15. Evidence of acute illness within the 4 weeks prior to screening that an Investigator deems may compromise volunteer safety.
16. Blood donation of any volume within one month before inclusion, or participation in any research trial involving blood sampling (more than 300 mL/unit of blood) within one month prior to IMP administration, or blood donation to Life Blood (Blood Service) or other blood bank during the 8 weeks prior to IMP administration.
17. History or presence of alcohol abuse (alcohol consumption more than 40 g/4 units/4 standard drinks per day), or drug habituation, or any prior intravenous usage of an illicit substance.
18. Any individual who has ever smoked >1 pack of cigarettes per day for >10 years, or who currently (within 14 days prior to inoculation) smokes >5 cigarettes/day.
19. Female who is breastfeeding.
20. Any COVID-19 vaccine within 14 days of IMP administration, any other vaccination within 28 days of IMP intake, and any vaccination (including for COVID-19) planned up to the final follow-up visit.
21. Any corticosteroids, anti-inflammatory drugs (excluding commonly used over-the-counter anti-inflammatory drugs such as ibuprofen, acetylsalicylic acid, diclofenac), immunomodulators or anticoagulants within the past three months. Any individual currently receiving or having previously received immunosuppressive therapy (including systemic steroids, adrenocorticotrophic hormone or inhaled steroids) at a dose or duration potentially associated with hypothalamic-pituitary-adrenal axis suppression within the past year.
22. Use of prescription drugs (excluding contraceptives) or non-prescription drugs or herbal supplements (such as St John's Wort), within 14 days or five half-lives (whichever is longer) prior to Inoculation. Limited use of other non-prescription medications or dietary supplements, not believed to affect volunteer safety or the overall results of the trial, may be permitted on a case-by-case basis following approval by the Sponsor in consultation with an Investigator. Volunteers are requested to refrain from taking non-approved concomitant medications from recruitment until the conclusion of the trial.
23. Cardiac/QT risk:
- Family history of sudden death or of congenital prolongation of the QTc interval or known congenital prolongation of the QTc interval or any clinical condition known to prolong the QTc interval.
- History of symptomatic cardiac arrhythmias or with clinically relevant bradycardia.
- Electrolyte disturbances, particularly hypokalaemia, hypocalcaemia, or hypomagnesaemia.

  ECG abnormalities in the standard 12-lead ECG (at screening, prior to IMP dosing, and prior to inoculation) which in the opinion of an Investigator is clinically relevant or will interfere with the ECG analyses.
24. Any history of malaria or participation in a previous malaria challenge trial or malaria vaccine trial.
25. Must not have had malaria exposure that is considered by the Investigator or delegate to be significant. This includes but is not limited to: history of having travelled to or lived (>2 weeks) in a malaria-endemic region during the past 12 months or planned travel to a malaria-endemic region during the course of the trial; history of having lived for >1 year in a malaria-endemic region in the

past 10 years; history of having ever lived in a malaria-endemic region for more than 10 years inclusive. For endemic regions see https://map.ox.ac.uk/country-profiles/#!/. Bali is not considered a malaria-endemic region.

26. Has evidence of increased cardiovascular disease risk (defined as >10%, 5-year risk for those greater than 35 years of age, as determined by the Australian Absolute Cardiovascular Disease Risk Calculator [http://www.cvdcheck.org.au/]). Risk factors include sex, age, systolic blood pressure (mm/Hg), smoking status, total and HDL cholesterol (mmol/L), and reported diabetes status.
27. History of splenectomy.
28. Individual unwilling to defer blood donations to the Blood Service for at least six months after the EOS visit.
29. Individual who has ever received a blood transfusion.
30. Any recent (<6 weeks) or current systemic therapy with an antibiotic or drug with potential antimalarial activity (e.g. chloroquine, piperaquine phosphate, benzodiazepine, flunarizine, fluoxetine, tetracycline, azithromycin, clindamycin, doxycycline etc.).

**General conditions**

31. Any individual who, in the judgement of an Investigator, is likely to be non-compliant during the trial, or is unable to cooperate because of a language problem or poor mental development.
32. Any individual in the exclusion period of a previous trial according to applicable regulations.
33. Any individual who is an Investigator, research assistant, pharmacist, trial coordinator, or other staff thereof, directly involved in conducting the trial.
34. Any individual without good peripheral venous access.

**Biological status**

35. Positive result on any of the following tests: hepatitis B surface antigen (HBs Ag), anti-hepatitis B core antibodies (anti-HBc Ab), anti-hepatitis C virus (anti-HCV) antibodies, anti-human immunodeficiency virus 1 and 2 antibodies (anti-HIV1 and anti-HIV2 Ab).
36. Recent herpes zoster infection (within the previous 6-months) as determined by clinical history.
37. Positive result for *M. tuberculosis* infection by QuantiFERON-TB Gold assay.
38. Positive urine drug test. Any drug listed in the urine drug screen unless there is an explanation acceptable to an Investigator (e.g., the volunteer has stated in advance that they consumed a prescription or over-the-counter product that contained the detected drug) and the volunteer has a negative urine drug screen on retest by the pathology laboratory. Any individual testing positive for acetaminophen (paracetamol) at screening and/or inoculation day may still be eligible for trial participation, at Investigator's discretion.
39. Positive alcohol breath test.

#### 3.3 LIFESTYLE CONSIDERATIONS

While participating in this trial, volunteers are asked to:

- Refrain from alcohol consumption of more than 20 g/2 units/2 standard drinks per day from 24 hours before first inoculation to Day 28 and from 24 hours before second inoculation until the end of study visit.
- Abstain from any drug habituation until the conclusion of the trial.
- Abstain from any alcohol and drug use for the duration of clinical trial unit confinement.
- Refrain from tobacco use of more than five cigarettes or equivalent per day until the conclusion of the trial.
- Abstain from any tobacco use for the duration of clinical trial unit confinement.
- Refrain from excessive consumption of beverages or food containing xanthine bases including Red Bull, chocolate, coffee etc. (more than 400 mg caffeine per day, equivalent to more than 4 cups of coffee per day).
- Abstain from any consumption of beverages or food containing xanthine bases for the duration of clinical trial unit confinement.
- Refrain from consumption of Seville oranges, and grapefruit or grapefruit juice from 7 days prior to first inoculation until Day 28.
- Refrain from consumption of quinine containing foods/beverages such as tonic water and lemon bitter from inoculation day until the EOS.
- Abstain from strenuous exercise for 24 h before each blood collection for clinical laboratory tests. Volunteers may participate in light recreational activities during studies.

#### 3.4 SCREEN FAILURES

Healthy candidate volunteers who do not fulfil all the inclusion criteria, and/or fulfil any of the exclusion criteria should not be enrolled into the trial without exception. In case of doubt, an Investigator is to confer with the Medical Monitor for agreement. Waivers for inclusion of volunteers who are not meeting all eligibility criteria will not be granted.

#### 3.5 STRATEGIES FOR RECRUITMENT AND RETENTION

Up to 26 volunteers will be enrolled in this trial. It is anticipated that a total of 7 reserve volunteers will be required to ensure adequate volunteer numbers.

Volunteers will be recruited by a general or trial specific advertisement via print, radio, social media, or poster media or educational presentations to students of Queensland universities and/or to the general community, as approved by the HREC(s). No restrictions will apply for ethnic or racial categories.

Volunteers who withdraw or are withdrawn from the trial will be compensated on a fractional basis for their involvement unless they are withdrawn as a consequence of their misconduct.

Reserve volunteers who do not participate in the trial will be compensated for the inconvenience associated with their attendance for screening and attendance on inoculation day.

The monetary value of the compensation for each trial part is documented in the respective Participant Information Sheet.

Volunteers who fail screening due to an underlying medical condition previously unknown to them will be reimbursed for their time, and provided with the appropriate referrals for guidance and counselling for their condition.

### 4 TRIAL INTERVENTION

#### 4.1 TRIAL INTERVENTION(S) ADMINISTRATION

##### 4.1.1 TRIAL INTERVENTION DESCRIPTION

**Malaria challenge agent**

To date, the *P. falciparum* 3D7 challenge agent has been used to challenge 397 malaria-naïve volunteers in 32 completed clinical studies, of which 335 malaria-naïve volunteers were successfully inoculated in 27 studies undertaken on the RBWH Health Campus at QIMR Berghofer/Q-Pharm. The *P. falciparum* 3D7 MCB was produced from an individual with blood type O (RhD) negative who was infected with the parasite by mosquito bite [40]. The 3D7 MCB was cryopreserved, aliquoted into cryovials, and stored in liquid nitrogen under controlled conditions. Refer to the *P. falciparum* 3D7 Investigator's Brochure for more details [20]. 3D7 MCB cryovials will be retrieved from storage, thawed, and used to aseptically prepare the 3D7 challenge agent at the Q-Gen Cell Therapeutics GMP facility at QIMR Berghofer (hereafter referred to as Q-Gen). For the two inoculations proposed in this study, each 3D7 inoculum dose will be prepared aseptically from an aliquot of the *P. falciparum* 3D7 MCB.

**Investigational Medicinal Products**

Riamet® (artemether-lumefantrine [AL])

Riamet® is a fixed combination antimalarial treatment containing artemether and lumefantrine. Artemether inhibits an essential calcium adenosine triphosphatase. The exact mechanism of lumefantrine is unknown.

Jakavi® (ruxolitinib [Rux])

Jakavi® is a kinase inhibitor containing ruxolitinib phosphate. Ruxolitinib mediates the signalling of a number of cytokines and growth factors that are important for haematopoiesis and immune function.

**Antimalarial rescue treatments**

Primacin™ (if required):

Primacin™ is an antimalarial agent containing primaquine (active ingredient) and phosphate. Primaquine phosphate is effective against the sexual forms (gametocytes) of Plasmodia, especially *P. falciparum*, disrupting transmission of the disease by eliminating the reservoir from which the mosquito carrier is infected.

Malarone® (if required):

Malarone® is a fixed combination antimalarial treatment containing atovaquone and proguanil hydrochloride. Atovaquone interferes with the biosynthesis of pyrimidines by blocking the parasite mitochondrial electron transport chain. Proguanil interferes with the biosynthesis of pyrimidines by inhibiting parasite dihydrofolate reductase.

Artesunate (if required): Artesunate is an artemisinin derivative. IV artesunate may be used as a rescue medication if a volunteer cannot tolerate oral drugs. The volunteer would be admitted to, and managed in hospital. This drug is the recommended parenteral treatment for malaria in Australia.

##### 4.1.2 DOSING AND ADMINISTRATION

**Malaria challenge agent**

Each *P. falciparum* 3D7 challenge agent dose will contain parasitised and non-parasitised RBCs, resuspended in 0.9% Sodium Chloride Intravenous Infusion, in a total volume of 2 mL in syringes. The syringes will be double contained following preparation and labelled in accordance with Good Clinical Practice (GCP) guidelines and the *Australian clinical trial handbook: guidance on conducting clinical trials in Australia using 'unapproved' therapeutic goods* [41]. Each volunteer administered 3D7 will be inoculated intravenously with a dose of approximately 2,800 viable *P. falciparum* 3D7-infected erythrocytes in 2 mL of saline for injection.

**Investigational Medicinal Products**

Riamet® (artemether-lumefantrine [AL])

Two courses of Riamet® will be administered to all 26 volunteers during the study (one course after the first malaria inoculation and another course after the second inoculation). Each tablet contains 20 mg artemether and 120 mg lumefantrine. The standard adult dosing regimen will be used in this trial: 6 doses of 4 tablets administered orally twice daily over 3 consecutive days (total course of 24 tablets). Each dose should be taken with food or drinks rich in fat (e.g., milk). No food permitted within 30 minutes prior to AL dosing, or between AL and Rux dosing. Meals are not required to be standardized. Times of meals and completion times will be documented.

The volunteers first treatment course (after the first malaria inoculation) will commence on Day 8 when parasitaemia for the majority of volunteers is expected to be above 5,000 parasites/mL. Earlier treatment for individual volunteers will be initiated if:

- they experience a serious adverse event (SAE) related to the malaria challenge agent, or
- they have a grade 3 AE graded in accordance with the Common Terminology Criteria for Adverse Events (CTCAE) deemed related to malaria and not self-resolved or relieved with concomitant medications, or
- the Investigator considers it necessary for volunteer safety

The second treatment course (after the second malaria inoculation) will commence on an individualised basis, when:

- qPCR parasitaemia reaches ≥50,000 parasites/mL, or
- they have a malaria clinical score >6, and presence of parasitaemia, or
- they experience an SAE related to the malaria challenge agent, or
- they have a CTCAE grade 3 AE deemed related to malaria and not self-resolved or relieved with concomitant medications, or
- the Investigator considers it necessary for volunteer safety.
- **If none of the above criteria for AL administration are reached by Day 118±7 (28 days after second inoculation) then compulsory AL administration must occur.**

Jakavi® (ruxolitinib [Rux])

A single course of Jakavi® will be administered only to the 13 volunteers randomised to the active group (after the first malaria inoculation).

The equivalent of the standard adult dosing regimen according to the PI and CMI will be used: 1 tablet (1× 20 mg) administered orally with 250 mL water b.i.d over 3 consecutive days (6 doses, total course of 6 tablets) and will be taken 2 hours after administration of AL. Participants will be instructed to swallow the tablet whole without biting or chewing.

Doses will be administered by an unblinded staff member (not an investigator). Volunteers will be blindfolded for dosing; other participants will be prevented from witnessing dosing.

No food permitted between AL and Rux, and for at least 1 hour post-Rux. Meals are not required to be standardized. Times of meals and completion times will be documented.

Placebo

A placebo in tablet form will be administered only to the 13 volunteers randomised to the placebo group (after the first malaria inoculation). The same dosing procedures and food restrictions to those described for Rux above will apply.

**Antimalarial rescue treatments**

Primacin® (if required): Volunteers may be treated with Primacin® if gametocytaemia is suspected from parasite lifecycle stage qRT-PCR to ensure complete clearance of gametocytes. If needed, volunteers will take six Primacin® tablets (the total dose of 45 mg primaquine) as a single dose with food. Volunteers that are mildly G6PD deficient will take two Primacin® tablets (the total dose of 15 mg primaquine) as a single dose with food. Volunteers who are severely G6PD deficient will not be administered Primacin®. Volunteers will be reminded of the potential side effects of Primacin® and will be given the CMI for Primacin®.

Malarone® (if required): If an allergy or contraindication to Riamet® develops, Malarone® may be administered at Investigator's discretion. The dose administered will be as recommended by the manufacturer for treatment of malaria. A treatment course of Malarone® consists of four tablets of Malarone® (atovaquone 250 mg, proguanil hydrochloride 100 mg) once daily orally for three days. Volunteers will be reminded of the potential side effects of Malarone® and will be given the CMI for Malarone®.

Artesunate (if required): Intravenous artesunate may be used as a rescue medication if the volunteer cannot tolerate oral drugs. Artesunate dosage will be manged in hospital.

#### 4.2 PREPARATION/HANDLING/STORAGE/ACCOUNTABILITY

##### 4.2.1 ACQUISITION AND ACCOUNTABILITY

**Malaria challenge agent**

The *P. falciparum* 3D7 MCB was produced from an individual with O Rh (D) negative blood who was infected with the parasite by mosquito bite. The 3D7 MCB was cryopreserved, aliquoted into cryovials and stored in liquid nitrogen under controlled conditions. Refer to the 3D7 Investigator's Brochure for more details [20]. On the day of each inoculation a *P. falciparum* 3D7 MCB cryovial will be retrieved from storage, thawed, and used to aseptically prepare the *P. falciparum* 3D7 inoculum at Q-Gen.

The clinical trial unit pharmacist or designee, as nominated by the trial pharmacist, will be responsible for maintaining the accurate *P. falciparum* 3D7 Challenge Agent Accountability Log as per the clinical trial unit standard operating procedures (SOPs).

**Investigational Medicinal Products**

Riamet® (artemether-lumefantrine [AL])

Riamet® (distributed by Novartis Pharmaceuticals Pty Ltd) will be acquired by the clinical trial unit. The supplies will be held in appropriate locked storage conditions at the clinical trial unit until required. The contents of the dispensing label for the drug to be administered to the volunteers will be in accordance with all applicable regulatory requirements.

The number of Riamet® tablets will be inventoried prior to the beginning of trial enrolment on trial accountability logs in regard to condition upon receipt, including lot numbers. The Investigator or qualified designee will ensure that the received drugs are the specified formulation. The site pharmacist or qualified designee is responsible for maintaining an accurate inventory and accountability record of drug supplies for this trial.

Jakavi® (ruxolitinib [Rux])

Jakavi® tablets (distributed by Novartis Pharmaceuticals Pty Ltd) will be acquired by the Sponsor from PCI Pharma. The supplies will be held in appropriate locked storage conditions at the clinical site until required. The contents of the dispensing label for the drug to be administered to the volunteers will be in accordance with all applicable regulatory requirements.

The number of Jakavi® tablets will be inventoried prior to the beginning of trial enrolment on trial accountability logs in regard to condition upon receipt, including lot numbers. The Investigator or qualified designee will ensure that the received drugs are the specified formulation. The site pharmacist or qualified designee is responsible for maintaining an accurate inventory and accountability record of drug supplies for this trial.

Placebo

Placebo tablets will be acquired by the Sponsor from PCI Pharma. The supplies will be held in appropriate locked storage conditions at the clinical site until required. The contents of the dispensing label for the drug to be administered to the volunteers will be in accordance with all applicable regulatory requirements.

The number of placebo tablets will be inventoried prior to the beginning of trial enrolment on trial accountability logs in regard to condition upon receipt, including lot numbers. The Investigator or qualified designee will ensure that the received drugs are the specified formulation. The site pharmacist or qualified designee is responsible for maintaining an accurate inventory and accountability record of drug supplies for this trial.

**Antimalarial rescue treatments**

Primacin® (distributed by Boucher & Muir Pty Limited) and Malarone® (distributed by GlaxoSmithKline Australia Pty Ltd) will be acquired by the clinical trial unit (if required).

Artesunate is the recommended parenteral treatment for malaria in Australia and in the rare event a volunteer/s are unable to complete oral antimalarial rescue treatment, they will admitted to hospital to receive intravenous artesunate.

The clinical trial unit pharmacist or delegate, as nominated by the PI, is responsible for maintaining accurate trial intervention accountability records throughout the trial. Trial interventions include the malaria challenge agent, IMPs and the antimalarial rescue treatments. Dispensing, accountability and documentation will be in accordance with standard procedures.

All products will be inventoried upon receipt by the clinical trial unit pharmacist. The condition of the products at the time of receipt by the pharmacist will be documented, as will the time restrictions of use for the syringes containing the challenge agents. The lot numbers and expiry dates of the challenge agents and antimalarial drugs will be documented. The clinical trial unit pharmacist or delegate will ensure that the received products are the specified formulation.

The storage, handling and the disposal of the challenge agents will be in accordance with approved procedures. All dosages prescribed and dispensed to the volunteers and all dose changes during the trial must be recorded in the eCRFs. All drug supplies are to be used only in accordance with this protocol, and not for any other purpose. All used medications will be fully documented. Used and unused drug containers must be destroyed at the site once drug accountability is final and has been checked by the Sponsor or its delegate, and written permission for destruction has been obtained from the Sponsor.

Trial products and trial accountability logs will be available to the Sponsor or their representative as part of the trial monitoring procedures. Upon completion of the trial, copies of all trial drug management records will be provided to the Sponsor. Original records will be maintained at the clinical trial unit with the rest of the trial records.

##### 4.2.2 FORMULATION, APPEARANCE, PACKAGING, AND LABELING

**Malaria challenge agent**

The malaria challenge agent will contain parasitised and unparasitised RBCs, resuspended in 0.9% Sodium Chloride Intravenous Infusion, in a total volume of 2 mL in syringes. The syringes will be double contained following preparation and labelled in accordance with GCP guidelines and the *Australian clinical trial handbook: guidance on conducting clinical trials in Australia using 'unapproved' therapeutic goods* [41].

**Investigational Medicinal Products**

Riamet® (artemether-lumefantrine [AL])

Riamet® tablets (each containing 20 mg artemether/120 mg lumefantrine) are yellow, round, flat tablets marked with N/C and a score line on one side and CG on the other side. Each carton contains 24 tablets.

Jakavi® (ruxolitinib [Rux])

Jakavi® tablets (containing 20 mg ruxolitinib) will be supplied as elongated, curved white to almost-white tablets with 'L20' on one side and 'NVR' on the other. Each pack contains 56 tablets.

Placebo

Placebo tablets will be prepared by PCI Melbourne and filled with microcrystalline cellulose (MCC) only. PCI Melbourne will fill all tablets into one bulk container. A single panel label will be applied.

**Antimalarial rescue drugs**

Primacin® (if required): Primacin® tablets (each containing 13.2 mg primaquine phosphate equivalent to 7.5 mg primaquine) are round, flat, orange uncoated tablets available in bottles of 28 or 56 tablets.

Malarone® (if required): Malarone® tablets are round, pink and film-coated, and are engraved with GX CM3. Malarone® tablets are supplied in blister packs of 12 or 24 tablets.

The contents of the dispensing labels for the malaria challenge agent, the IMPs, and the antimalarial rescue medications will be in accordance with all applicable regulatory requirements.

##### 4.2.3 PRODUCT STORAGE AND STABILITY

**Malaria challenge agent**

The challenge agent will be prepared at Q-Gen on each inoculation day. The time between preparation of the challenge agent and administration to the volunteer will be a maximum of 4 h. The syringes containing the challenge agent will be stored in a validated transport container at 2-15°C during transportation from Q-Gen to the clinical trial unit and will be immediately transferred to a temperature monitored onsite refrigerator (2-8°C). The clinical trial unit pharmacist will document receipt conditions and time restrictions of use. The challenge agent will then be dispensed to volunteers as per written prescription.

**Investigational Medicinal Products**

Riamet®, Jakavi®, and the placebo tablets must be stored below 30 °C in tightly closed containers, protected from light. All drugs will be held in appropriate locked storage conditions at the clinical trial unit until required.

**Antimalarial rescue drugs**

Primacin® is to be stored below 25°C. Malarone® is to be stored below 30°C.

##### 4.2.4 PREPARATION

**Malaria challenge agent**

The challenge agent will be prepared aseptically at Q-Gen from a frozen cryovial of the *P. falciparum* 3D7 MCB, by trained QIMR Berghofer staff members under the guidance of the Investigator. The infected RBCs will be thawed, washed, resuspended in saline, diluted in a final volume of 2 mL of clinical grade saline, and dispensed into syringes. Any remaining unused infected RBCs will be discarded as per approved SOPs.

**Investigational Medicinal Products**

Riamet®, Jakavi® and the placebo are available as tablets and no preparation is required.

**Antimalarial rescue drugs**

Primacin® and Malarone® are available as tablets and no preparation is required. Artesunate will be prepared and administered in hospital.

#### 4.3 MEASURES TO MINIMISE BIAS: RANDOMISATION AND BLINDING

##### 4.3.1 RANDOMISATION AND BLINDING PROCEDURES

Randomisation procedures will be as described in the pharmacy manual.

Briefly, a randomisation number will be allocated to each volunteer as per the randomisation schedule immediately before the administration of IMPs (AL+Rux or AL+placebo).

The volunteers and the Investigators will be blinded to IMP allocation. Since the placebo tablets will not be matched to Rux tablets in terms of appearance, doses will be administered by an unblinded staff member (not an investigator). Volunteers will be blindfolded for dosing; other participants will be prevented from witnessing dosing. Blinding will be maintained throughout the duration of the trial until all final clinical data have been entered into the database and all data queries have been resolved, and the assignment of volunteers to the analysis sets has been completed. The randomisation list is to be kept strictly confidential, accessible only to authorised persons (e.g., randomisation statistician, pharmacists, and the laboratories that prepare and analyse relevant samples), until the time of unblinding. Interim safety and PK analyses and SDRT meetings will be performed blinded, utilising dummy volunteer numbers which cannot be linked to the volunteer identifiers used in the trial site. The study should not be unblinded except in a medical emergency where knowledge of the study drug received would affect the treatment of the volunteer. If emergency unblinding becomes necessary, the PI should notify the Sponsor and the Medical Monitor, if possible, before unblinding. All cases resulting in an unblinding event will be documented and reported to the Medical Monitor and the Sponsor. If unblinding occurs, the date, time and reason must be recorded in the volunteer’s eCRF and any associated AE report completed.

The level of blinding will be maintained throughout the conduct of the trial, and only when the data are cleaned to an acceptable level of quality will appropriate personnel be unblinded. Any intentional or unintentional breaking of the blind will be reported and explained at the end of the trial, irrespective of the reason for its occurrence.

#### 4.4 TRIAL INTERVENTION COMPLIANCE

**Malaria challenge agent**

On inoculation days (Day 0 and Day 90±7), volunteers will attend the clinical trial unit and will undergo IV cannulation with an appropriate gauge cannula. Placement and patency will be checked by flushing the vein with 5-10 mL of clinical grade saline. The malaria challenge agent will be administered and the cannula will then be removed, and haemostasis ensured by use of an appropriate dressing. The volunteers will be inoculated within the expiry time noted on the syringe.

**Investigational Medicinal Products**

Riamet® (artemether-lumefantrine [AL])

After their first inoculation, all volunteers will be confined in the clinical trial unit for administration of all doses of AL, and will remain confined for a minimum 12 h after the last dose of AL is administered. Trained clinical trial unit staff will ensure volunteers have swallowed the AL tablets after each administration.

All volunteers who have a second inoculation will receive their first dose of AL at the clinical trial unit and take the remaining doses at home. These volunteers will receive a daily phone call or text message from clinical trial unit staff to check on symptoms and ensure compliance and completion of treatment (Riamet®) following the doses taken at home.

Jakavi® (ruxolitinib [Rux])

All volunteers will be confined in the clinical trial unit for administration of all doses of Rux, and will remain confined for a minimum 10 h after the last dose of Rux is administered. Trained clinical trial unit staff will ensure volunteers have swallowed the Rux tablets after each administration.

Placebo

As above, as per Jakavi® (ruxolitinib [Rux]).

**Antimalarial rescue treatments**

The first dose of Primacin™ and Malarone® if required will be administered during visits at the clinical trial unit. The subsequent doses of each treatment may be taken at home and volunteers will receive a phone call or text message from the clinical trial unit staff to ensure compliance and completion of treatments. If a volunteer requires intravenous artesunate, the volunteer will be admitted to hospital and managed there for the duration of the duration of the treatment.

#### 4.5 PRIOR AND CONCOMITANT THERAPY

Prior medications, treatments and procedures are those occurring prior to inoculation. Concomitant medications, treatments, and procedures are those occurring from malaria inoculation until EOS.

Medications to be reported in the eCRF are concomitant prescription medications, over-the-counter medications, and supplements. For this protocol, a prescription medication is defined as a medication that can be prescribed only by an authorised/licensed clinician. Prior and concomitant medications, treatments, and procedures permitted in this trial are outlined in the inclusion and exclusion criteria (Section 3.1 and 3.2).

On inoculation days, volunteers will be questioned about relevant aspects of compliance with the trial protocol, including medication use since their screening visit or continued eligibility visit. Details of all medication use (prescription and over-the-counter, systemic, and topical administration) from the screening visit or continued eligibility visit will be recorded at this time and appropriate action taken. The Investigator may permit the use of ibuprofen up to 1.2 g/day or paracetamol up to 4 g/day for treatment of headache or other pain if required. Ibuprofen is the preferred treatment for headache or other pain. To minimise the risk of liver enzyme elevation, paracetamol is to be avoided if possible; however, paracetamol may be required by some volunteers and, as such, is not prohibited. The exact dose and timing of each dose of any concomitant medication should be recorded (i.e., 'As needed' or 'PRN' or similar notation should be avoided where possible).

##### 4.5.1 RESCUE MEDICINE

The rescue medications used in this trial are Malarone® and Primacin™ (if required). See details in Section 4.1.1. If a volunteer vomits or cannot tolerate oral drugs then artesunate will be administered intravenously as described in Section 4.1.1.

### 5 TRIAL INTERVENTION DISCONTINUATION AND VOLUNTEER DISCONTINUATION/ WITHDRAWAL

#### 5.1 DISCONTINUATION OF TRIAL INTERVENTION

The Sponsor, PI, approving HREC(s), and the TGA independently reserve the right to discontinue the trial at any time for safety or other reasons. This will be done in consultation with the Sponsor where practical. In the event of premature trial termination or suspension, the above-mentioned parties will be notified in writing by the terminator/suspender stating the reasons for early termination or suspension (with the exception of the TGA as it is the Sponsor's responsibility to notify regulatory authorities). After a decision to prematurely terminate the trial or to suspend the trial, the Sponsor and the PI will ensure that adequate consideration is given to protecting the volunteers' interest and safety. The PI must review all volunteers as soon as practicable and complete all required records.

In addition to the classic assessment of SAEs, specific toxicity rules apply for this trial and must be applied when early termination or suspension is considered. When applying toxicity rules, AEs will be graded using the CTCAE grading system for standardised recording and reporting. For the purpose of this protocol, "toxicity" refers to the occurrence of any grade 3 or higher grade AE that is related to an IMP (AL, Rux, and Placebo) or to the malaria challenge agent.

#### 5.2 VOLUNTEER DISCONTINUATION/WITHDRAWAL FROM THE TRIAL

Volunteers are free to withdraw from participation in the trial at any time upon request.

An Investigator may discontinue or withdraw a volunteer from the trial for the following reasons:

- Pregnancy

- Significant trial intervention non-compliance
- If any clinical AE, laboratory abnormality, or other medical condition or situation occurs such that continued participation in the trial would not be in the best interest of the volunteer
- If the volunteer meets an exclusion criterion (either newly developed or not previously recognised) that precludes further trial participation

The reason for volunteer discontinuation or withdrawal from the trial will be recorded on the eCRF. Volunteers who sign the informed consent form and are randomised but do not receive the trial intervention (first inoculation) may be replaced. Volunteers who sign the informed consent form, and are randomised and receive the trial intervention, and subsequently withdraw, or are withdrawn or discontinued from the trial, may be replaced after mutual agreement between the Sponsor and the Investigator. The decision regarding the replacement of volunteers will be documented.

Volunteers who have been inoculated and indicate they wish to withdraw from the trial must complete the full course of Riamet®.

Volunteers will be reminded of the importance of completing their antimalarial treatment. Additionally, the Investigator will demonstrate due diligence in following up with the volunteer to ensure antimalarial therapy has been completed successfully. Volunteers may also be treated with Primacin™ if gametocytaemia is suspected from parasite lifecycle qRT-PCR or by the presence of stable low-level parasitaemia to ensure complete clearance of gametocytes. The decision to administer Primacin™ will be made by the Investigator.

If a volunteer is withdrawn from the trial, the Sponsor will be informed immediately. If there is a medical reason for withdrawal, the volunteer will remain under the supervision of the PI until satisfactory health has returned, or medical/clinical care has been transferred to the volunteer's general practitioner or to a hospital consultant.

The Investigator will make every effort to determine the primary reason for a volunteer's withdrawal from the trial and record this information in the eCRF. If the volunteer is withdrawn from the trial procedures or follow-up for any reason, with the volunteer's permission, medical care will be provided for any SAEs that occurred during participation in the trial until the symptoms of any SAEs are resolved and the volunteer's condition becomes stable.

#### 5.3 LOST TO FOLLOW-UP

For volunteers who are lost to follow-up, the Investigator will apply due diligence by documenting all steps taken to contact the volunteer (e.g., dates of phone calls, registered letter, home visit, etc.) in the source documents.

It will be explained to the volunteers during the consenting process and throughout the trial that they must be readily contactable. The period from inoculation to antimalarial treatment is the critical period for volunteer safety because they will have blood stage parasites. A volunteer will be deemed “missing” if they fail to reply to communication to their personal mobile phone and nominated contact’s number after 24 h. If after 36 h the volunteer fails to respond then an Investigator will organise a home visit. Subsequently, if the volunteer is still absent, an Investigator will request assistance from the local police to locate the missing volunteer. Once the volunteer is found and if the volunteer is still parasitaemic, then at the PI’s discretion the volunteer will be administered Riamet® and if clinically relevant Primacin™.

### 6 TRIAL ASSESSMENTS AND PROCEDURES

#### 6.1 TRIAL SPECIFIC PROCEDURES

Medical history as described below will be elicited at:

- Screening, continued eligibility (Day 85±7) and Inoculation days (Day 0 and Day 90±7)

| Medical/Surgical History includes: |
|---|
| History of all known drug/food allergies and/or asthma |
| Current medications, including over-the-counter and herbal preparations |
| History of substance abuse and recreational drug use |
| History of depression, anxiety, mental illness, emotional problems, use of psychiatric medications and previous psychotherapy |
| Surgical procedures and results |
| Any other current or past medical conditions |

##### 6.1.1 PHYSICAL EXAMINATION

A **full physical examination** includes the following:

| |
|---|
| Weight |
| Height (***Screening only***) |
| Review of systems excluding genitourinary examination and including the following: |
| Head, neck (including thyroid), ears, eyes, nose and throat |
| Heart/circulation |
| Chest |
| Lungs |
| Abdomen |
| Skin |
| Neurological exam |

An **abbreviated physical examination** will include heart/circulation, chest, lungs, skin, and abdomen.

When the **symptom-directed physical examination** is performed, body systems will be reviewed only if clinically indicated.

##### 6.1.2 BECK DEPRESSION INVENTORY

All volunteers will be required to complete the Beck Depression Inventory at screening and at the continued eligibility visit on Day 85±7. This is a validated questionnaire used to screen for depression.

##### 6.1.3 VITAL SIGNS

Vital signs (temperature, heart rate, respiratory rate, and blood pressure) will be measured after the volunteer has rested in the supine position for at least 5 min and in the standing position within 2–3 min when changing from the supine to standing position (blood pressure and heart rate only) at Screening and continued eligibility (Day 85±7)

At all other timepoints, vital signs will be measured after the volunteer has rested in the seated position for at least 5 min.

The acceptable vital sign ranges for trial inclusion are:

| **Parameter** | **Range** |
|---|---|
| Systolic blood pressure | 90-140 mmHg |
| Diastolic blood pressure | 40-90 mmHg |
| Heart rate | 40-100 bpm |

The normal ranges for vital signs once on trial are:

| **Parameter** | **Range** |
|---|---|
| Systolic blood pressure | 90-140 mmHg |
| Diastolic blood pressure | 50-90 mmHg |
| Heart rate | 50-100 bpm |
| Temperature | 35.0-37.5°C |
| Respiratory rate | 10-25 breaths/min |

##### 6.1.4 ELECTROCARDIOGRAPHS (ECG)

A 12-lead ECG will be recorded after the volunteer has rested supine for at least 5 min. ECG tracings will be retained and labelled as per standard procedures at the clinical trial unit and will be recorded in the eCRF. Any clinically significant findings will be discussed with the Medical Monitor and Sponsor and documented as AEs. An Investigator will sign and date each ECG as evidence of their review.

The acceptable ECG ranges for trial inclusion are:

| Parameter | Range |
|---|---|
| PR interval | ≤210 msec |
| QTcF | Males: ≤450 msec<br>Females: ≤470 msec |

The normal ECG ranges once on trial are:

| Parameter | Range |
|---|---|
| PR interval | ≤210 msec |
| QRS | 50–120 msec |
| QTcF | Males: ≤450 msec<br>Females: ≤470 msec |

##### 6.1.5 BLOOD SAMPLING

**Safety**

Blood will be collected for clinical laboratory evaluations including haematology, biochemistry, coagulation profile, serology, QuantiFERON-TB Gold assay, pregnancy testing and/or FSH testing, and for RBC alloantibodies. All safety samples will be sent to national laboratories as named in this protocol.

**Pharmacokinetic**

Blood will be collected to quantify plasma drug concentrations for PK analysis. All PK assays will be performed in Sponsor approved national or international laboratories yet to be determined.

Blood samples will be collected either by direct venepuncture or via an indwelling cannula inserted in a forearm vein (depending on the timepoint).

**Pharmacodynamic**

Blood will be collected to monitor parasitaemia, for immune response assessments and for the pSTAT3 inhibition assay.

Refer to the Laboratory Manual for sample preparation procedures and sample volumes.

All PD assays will be performed at Sponsor approved national or international laboratories as named in this protocol.

**Malaria related research**

Blood collected at already scheduled time-points may be utilised for HREC approved projects to support exploratory malaria related research. This research may include testing for the presence of antibodies against a range of common infections and vaccinations that are known to, or have the potential to affect immune status and development, including but not limited to SARS-CoV-2, influenza, Epstein Barr Virus, and Cytomegalovirus.

##### 6.1.6 URINE SAMPLING

Urine will be collected for urinalysis and drug screens.

##### 6.1.7 MALARIA CLINICAL SCORE

The 14 signs/symptoms frequently associated with malaria are graded using a 4-point scale (absent 0; mild: 1; moderate: 2; severe: 3) and summed to generate a total malaria clinical score (maximum score possible is 42) (Appendix 3). To determine severity of the 14 signs/symptoms we use the Common Terminology Criteria for Adverse Event Reporting (CTCAE) Grade 1 - 5 Version 5.0 Published: November 27, 2017. Mild (1) equates to CTCAE grade 1, Moderate (2) equates to CTCAE grade 2 and Severe (3) equates to CTCAE grade 3 or above.

##### 6.1.8 DIARY CARDS AND THERMOMETERS

Volunteers will be provided with diary cards and instructed to record symptoms they experience and concomitant medications they use during the trial. Volunteers will also be provided with thermometers to record any temperature readings during the trial in the event of symptoms of fever; sublingual temperature will be taken by volunteers at home for practical reasons. The diary cards will be collected at the EOS visit and any information in the diary cards will be entered in the eCRF.

##### 6.1.9 CONCOMITANT MEDICATIONS

The exact dose and frequency of concomitant medications used must be recorded. ‘As needed’ or ‘PRN’ are not suitable terms.

##### 6.1.10 ADVERSE EVENT RECORDING

AEs will be recorded as described in Section 6.5.5.

#### 6.2 LABORATORY PROCEDURES

The required pathology tube types, blood volumes required for each procedure and laboratory processing procedures will be described in the Laboratory Manual.

Any significant deviations from results obtained during screening will be followed until resolution or investigated fully, or until the volunteer is referred to a general practitioner or medical specialist as appropriate. An Investigator will document the clinical significance of all results falling outside of the normal reference ranges. All abnormal laboratory test results judged as being clinically significant will be recorded as AEs.

Blood and urine samples for clinical laboratory assessments will be collected according to clinical trial unit SOPs. The parameters that will be measured are listed below. Additional reflex testing may be conducted by the local laboratory (as per their SOPs) if safety laboratory values for a volunteer fall outside of the normal range/parameters. Unscheduled testing may be performed at Investigator's discretion.

##### 6.2.1 HAEMATOLOGY

| |
|---|
| Full blood count (FBC) with differential |
| White blood cell count (WBC) |
| WBC differential (diff) |
| A manual blood smear should be reviewed if there are immature/abnormal cells detected on the automated differential or if an automated differential was not able to be performed. |
| Neutrophils (NEUT) |
| Lymphocytes (LYM) |
| Monocytes (MON) |
| Eosinophils (EOS) |
| Basophils (BAS) |
| Mean corpuscular volume (MCV) |
| Red blood cell count (RBC) |
| Haemoglobin (HGB) |
| Haematocrit (HCT) |
| Platelet count (PLAT) |
| Red blood cell distribution width (RDW) |
| Reticulocyte count (RETI) |
| Blood Group and Rh(D) tests (***Screening only***) |

##### 6.2.2 BIOCHEMISTRY

**General biochemistry**

| |
|---|
| Sodium (NA) |
| Potassium (K) |
| Chloride (CL) |
| Bicarbonate (BICARB) |
| Glucose (GLUC) |
| Urea |

| |
|---|
| Creatinine (CREAT) |
| Estimated glomerular filtration rate (eGFR) – CKD Epi calculation |
| Albumin (ALB) |
| Globulin |
| Total protein |
| Total bilirubin (BILI) |
| Direct (conjugated) bilirubin (BILDIR) |
| Alkaline phosphatase (ALP) |
| Alanine aminotransferase (ALT) |
| Aspartate aminotransferase (AST) |
| Gamma-glutamyl transferase (GGT) |
| Lactate dehydrogenase (LDH) |
| Calcium (CA) |
| Corrected calcium (CCA) |
| Magnesium (MAG) |
| Phosphate (PHOS) |
| C-Reactive Protein (CRP) (***Not included at screening***) |

**Iron studies**

| |
|---|
| Serum iron |
| Serum ferritin |
| Transferrin or total iron binding capacity (TIBC) |
| Transferrin saturation |

**Lipids**

When lipid testing is performed, volunteers must fast for 8 h prior to sample collection.

| |
|---|
| Total cholesterol |
| High density lipoprotein (HDL) |
| Low density lipoprotein (LDL) |
| Triglycerides |

##### 6.2.3 URINALYSIS

Urine will be tested by dipstick at the clinical trial unit. If there are any abnormalities considered clinically significant in blood, leukocytes, or protein, the urine will be sent for formal laboratory urinalysis per the clinical trial unit standard procedure for microscopy, culture and sensitivity.

| |
|---|
| Glucose (GLUC) |

| |
|---|
| Bilirubin (BILI) |
| Ketone (KETONES) |
| Specific gravity (SPGRAV) |
| Blood |
| pH |
| Protein (PROT) |
| Urobilinogen (UROBIL) |
| Nitrite |
| Leukocytes (WBC) |
| Microscopy, culture and sensitivity (***if required***) |

##### 6.2.4 URINE DRUG SCREEN AND ALCOHOL BREATH TESTS

All volunteers will be questioned about concomitant medications and use of recreational drugs. The urine drug screen may be repeated if the potential volunteer denies usage of any of these agents and the test result is believed to be a false positive.

| **Urine drug screen:** | |
|---|---|
| Amphetamines | Opiates |
| Methamphetamines | Phencyclidine |
| Barbiturates | Tetrahydrocannabinol (cannabis) |
| Benzodiazepines | Tricyclic antidepressants |
| Cocaine | |
| Methadone | |
| **Alcohol breath test** | |

If the results of the urine drug screens or alcohol breath tests are positive, volunteers will not be enrolled in the clinical trial unless there is an explanation acceptable to an Investigator (e.g., the volunteer has stated in advance that they consumed a prescription or over-the-counter product that contained the detected drug) and the volunteer has a negative urine drug screen on retest by the pathology laboratory.

##### 6.2.5 SEROLOGY

| |
|---|
| HIV 1/2 (anti-HIV1 and anti-HIV2 Ab) |
| Hepatitis B (HBsAg, anti-HBc [IgG + IgM if IgG is positive]) |
| Hepatitis C (anti-HCV) |
| Hepatitis A (anti-HAV) (IgM) - performed from stored sample for testing, in case of suspected viral infection |

| |
|---|
| Hepatitis E (anti-HEV) (IgM) - performed from stored sample for testing, in case of suspected viral infection |
| Epstein-Barr virus - performed from stored sample for testing, in case of suspected viral infection |
| Cytomegalovirus - performed from stored sample for testing, in case of suspected viral infection |

##### 6.2.6 PREGNANCY TESTING

At screening, and the continued eligibility visit Day 85±7 a serum β-hCG pregnancy test will be conducted in all female subjects, and FSH levels will be tested in post-menopausal females (at least one year post-menopausal). At all other required visits a urine β-hCG pregnancy tests will be conducted in WOCBP. If a subject is found to have a positive urine pregnancy test at any stage during the study, a confirmatory serum β-hCG will be performed. If confirmed positive, the subject will be discontinued from the study and rescued with Riamet®. Please refer to Section **6.5.9** for pregnancy-specific reporting procedures and follow-up.

##### 6.2.7 COAGULATION PROFILE

Blood for Prothrombin Time (PT), Activated Partial Thromboplastin Time (APTT) and International Normalised Ratio (INR) testing will be collected at screening.

##### 6.2.8 RBC ALLOANTIBODIES AND G6PD TESTING

Blood for RBC alloantibody testing will be collected for volunteers at screening, Day 28, continued eligibility visit (Day 85±7) and at the EOS visit.

Blood for G6PD testing will be collected at screening only. This test will not be done at the continued eligibility visit for the second inoculation.

##### 6.2.9 SAFETY SERUM RETENTION SAMPLES

Blood for safety serum retention samples will be collected at times outlined in Section **1.3.2-1.3.4.** Safety serum retention samples will be stored indefinitely at QIMR Berghofer for any retrospective safety assessments.

##### 6.2.10 QUANTIFERON-TB GOLD ASSAY

Blood to detect latent *M. tuberculosis* infection will be collected at screening only.

##### 6.2.11 PHARMACOKINETIC TESTING

**Drug concentration measurements (first inoculation)**

Blood sampling for PK analysis will be performed at scheduled timepoints from immediately before dosing with AL (Day 8) until Day 28.

Blood samples will be collected either by direct venepuncture or via an indwelling cannula inserted in a forearm vein. Details of the volume of blood collected are listed in the blood draw schedule.

A description of the required lab procedures will be described in the Laboratory Manual.

The actual sample collection date and time will be entered in the PK blood collection section of the eCRF. Any sampling problems will be documented in the eCRF.

**Pharmacokinetic analytical methods**

Plasma concentrations of Rux, AL and DHA will be quantified using a validated method by LC MS/MS in the selected reaction monitoring mode using heated electrospray ionisation in positive ion mode.

Plasma samples will be precipitated with five volume equivalents of a mixture of acetonitrile/methanol (4/1, v/v) containing the internal standard. After centrifugation, the samples will be diluted with a mixture of water/acetonitrile (1/1, v/v). An aliquot of 2 µL of the sample will be injected onto the high-performance liquid chromatography (HPLC) system.

##### 6.2.12 PHARMACODYNAMIC TESTING

**Malaria 18S qPCR (first and second inoculation)**

Blood will be collected to monitor parasitaemia using 18S qPCR. Refer to the Laboratory Manual for the sample preparation procedure and volume.

Additional blood may be collected for parasite lifecycle stage qRT-PCR to evaluate the presence of sexual parasite stages (gametocytes) and other parasite lifecycle stages in the blood. The qRT-PCR may target genes including but not limited to: the female gametocyte-specific transcript *pfs25*, the male gametocyte-specific transcript *PfMGET*, and the ring-stage transcript *pfSBP-1* as appropriate. The samples may be taken at timepoints indicated by malaria 18S qPCR, or at the Investigator’s discretion. If gametocytes are detected volunteers will be prescribed Primacin™.

Microscopic examination for evidence of parasitaemia or gametocytaemia may be conducted at the Investigator’s discretion.

**pSTAT3 assay (first innoculation)**

Blood sampling to perform the pSTAT3 assay will be performed at scheduled time-points. The assay is a validated whole blood assay and pSTAT3 levels will be determined by ELISA.

**Immune response assessments**

Blood sampling for immune response assessments will be performed at scheduled timepoints. The assays described in **appendix 3** will be performed. Time-points for immune response assessments may change based on data analysis from previous cohorts at the Investigator's discretion (refer to Lab Manual for each cohort).

#### 6.3 ALLOWED TIME WINDOWS

The following time windows will be allowed for blood sampling to quantify plasma drug concentrations, to monitor malaria parasitaemia and for pSTAT3 assay and immune response assessments:

| Time point | Tolerance window |
|---|---|
| **Pharmacokinetic/Pharmacodynamic** | |
| **In confinement (first inoculation)** | |
| Pre-dose (T=0 h) | - 4 h to 0 h |
| 1-2 hs inclusive after Rux/placebo administration | ± 2 min |
| 3-10 hs inclusive after Rux/placebo administration | ± 5 min |
| 11-72hs inclusive after Rux/placebo administration | ± 30 min |
| **Outpatient (first inoculation)** | |
| 79-168 h inclusive | ± 120 min |
| 169-672 hours inclusive | ± 24 h |
| **Outpatient (second inoculation)** | |
| Day 90±7 – AL administration (malaria monitoring) | ± 12 h |
| AL administration | - 180 min to 0 hr |
| AL administration – AL+3 | ± 24 h |
| AL+7- EOS | ± 72 h |

#### 6.4 SPECIMEN PREPARATION, HANDLING, STORAGE AND SHIPMENT

The allowed time windows for trial procedures and sample collections as described in **Section 6.3** will be followed. If the observation time and blood sampling time coincide, for precision of timing, blood collection will take precedence over other procedures scheduled at the same time.

Blood will be collected into tubes containing the appropriate anticoagulant. Samples will be processed according to the laboratory requirements.

Biological samples will be retained for the time required for assessment for analysis, and may then be discarded. Safety serum retention samples will be stored indefinitely with the permission of the volunteers for any retrospective safety assessments.

Samples collected will be shipped to nominated local or international laboratories for assessment. The clinical trial unit staff will be responsible for shipment of samples to analytical laboratories for testing. Samples must be packed securely together with completed shipment forms in shipping containers together with sufficient dry ice as per shipper procedures.

Refer to the Laboratory Manual for more details on samples preparation, handling, storage and shipment.

#### 6.5 ADVERSE EVENTS AND SERIOUS ADVERSE EVENTS

##### 6.5.1 DEFINITION OF ADVERSE EVENTS (AE)

An AE is any untoward medical occurrence, i.e., unfavourable and unintended sign (including an abnormal laboratory finding), symptom or disease that occurs in a volunteer during the course of the trial. An AE does not necessarily have a causal relationship with trial treatments or procedures.

AEs include, but are not limited to:

- A new symptom, sign or medical condition.
- A disease or medical condition detected or diagnosed during the course of the trial even though it may have been present prior to the start of the trial.
- An exacerbation of a pre-existing medical condition/disease.
- An increase in frequency or intensity of a pre-existing episodic disease or medical condition.
- Continuous persistent disease or symptoms present at trial start that worsen following the start of the trial.
- An abnormal assessment (e.g., change on physical examination, ECG finding) if it represents a clinically significant finding that was not present at trial start or worsened during the course of the trial.
- An abnormal laboratory test result if it represents a clinically significant finding (e.g., CTCAE grade 2 or above), symptomatic or not, which was not present at trial start or worsened during the course of the trial or led to dose reduction, interruption or permanent discontinuation of trial treatment.

Borderline abnormal laboratory findings and other objective assessments should NOT be routinely captured and reported as AEs. However, abnormal laboratory findings or other objective measurements that meet the following criteria should be captured and reported in the AE section of the eCRF:

- The result meets the criteria for reporting as an SAE.
- The test result is associated with accompanying symptoms, and/or
- It requires additional diagnostic testing or medical/surgical intervention, and/or
- It leads to a change in trial dosing, or discontinuation from the trial, significant additional concomitant drug treatment, or other therapy, and/or
- It is considered by an Investigator to be clinically significant or represent a clinically significant change from baseline.

Merely repeating an abnormal test, in the absence of any of the above conditions, does not constitute an AE. Any abnormal test result that is determined to be an error does not require reporting as an AE. Surgical procedures themselves are not AEs; they are therapeutic measures for conditions that may, or may not, be AEs.

##### 6.5.2 DEFINITION OF SERIOUS ADVERSE EVENTS (SAE)

An SAE is defined as an event that fulfils at least one of the following criteria:

- Results in death.
- Is life-threatening
  - The term "life-threatening" in the definition of "serious" refers to an event in which the volunteer was at immediate risk of death at the time of the event; it does not refer to an event which hypothetically might have caused death if it was more severe.
- Requires inpatient hospitalisation or prolongs existing hospitalisation, unless this is for:
  - Elective or pre-planned treatment for a pre-existing condition that is unrelated to the trial and has not worsened since the start of the trial.
  - Cosmetic surgery, or for social reasons, or respite care in the absence of any deterioration in the volunteer's general condition.
- Results in persistent or significant disability/incapacity.
- Is a congenital abnormality/birth defect.
- Is considered medically important
  - Medical and scientific judgement should be exercised in deciding whether other AEs are to be considered serious, such as important medical events that may not be immediately life-threatening but may jeopardise the volunteer or may require intervention to prevent one of the other outcomes listed in the definition above. Examples of such events are: intensive treatment in an emergency room or at home for allergic bronchospasm; blood dyscrasias; convulsions that do not result in hospitalisation; development of drug dependency or drug abuse.
- Constitutes a possible Hy's Law case
  - Hy's Law case is defined as a volunteer with any value of alanine or aspartate aminotransferase greater than 3×ULN together with an increase in total bilirubin to a value greater than 2×ULN and not associated to an alkaline phosphatase value greater than 2×ULN (FDA Guidance on Drug Induced Liver Injury: Premarketing Clinical Evaluation [2009]).

A **Suspected Unexpected Serious Adverse Reaction (SUSAR)** is any SAE where a causal relationship with a trial intervention (malaria challenge agent, Riamet®, Jakavi®, placebo, Malarone® or Primacin™ or Artesun®) is at least a reasonable possibility, and the event is not listed in the IB.

##### 6.5.3 CLASSIFICATION OF AN ADVERSE EVENT

###### 6.5.3.1 SEVERITY OF EVENT

In addition to determining whether an AE fulfils the criteria for an SAE or not, the severity of AEs experienced by volunteers will be recorded in accordance with the Common Terminology Criteria for Adverse Events (CTCAE) v5.0, published 27 November 2017.

The severity of AEs will be graded as follows:

- Grade 1: Mild; asymptomatic or mild symptoms; clinical or diagnostic observations only; intervention not indicated.
- Grade 2: Moderate; minimal, local or non-invasive intervention indicated; limiting age-appropriate instrumental activities of daily living.
- Grade 3: Severe or medically significant but not immediately life-threatening; hospitalisation or prolongation of hospitalisation indicated; disabling; limiting self-care activities of daily living.
- Grade 4: Life-threatening consequences; urgent intervention indicated.
- Grade 5: Death related to AE.

A mild, moderate, or severe AE may or may not be serious. These terms are used to describe the intensity of a specific event. Medical judgement should be used on a case-by-case basis.

Seriousness, rather than severity assessment, determines the regulatory reporting obligations.

For guidance for assigning severity of the malaria, the purpose-designed malaria clinical score will be used.

###### 6.5.3.2 RELATIONSHIP TO TRIAL INTERVENTION

An Investigator must assess the relationship of each event to the malaria challenge agent, the IMP, and the antimalarial rescue medications (separately) and decide whether, in his or her medical judgement, there is a reasonable possibility that the event may have been caused by any of the trial interventions. Where possible, a distinction should be made between events considered related to the malaria challenge agent, the IMPs, and the antimalarial rescue medications.

If there is no valid reason for suggesting a relationship, then the AE should be classified as “not related”. Alternatively, if there is any valid reason for suspecting a cause-and-effect relationship between the malaria challenge agent, the IMPs, or the antimalarial rescue medications and the occurrence of the AE (even if undetermined or untested), then the AE should be considered as “related” to whichever product is relevant. This should be documented in the volunteer’s clinical file (source data) and eCRF.

The following may guide this assessment:

**Related/suspected**:

The temporal relationship between the event and the administration of the malaria challenge agent and/or IMPs and/or the antimalarial rescue medications is compelling and/or follows a known or suspected response pattern to that product, and the event cannot be explained by the volunteer's medical condition, other therapies or accident.

**Not related/not suspected**:

The event can be readily explained by other factors such as the volunteer's underlying medical condition, concomitant therapy or accident and no plausible temporal or biologic relationship exists between the challenge agent and/or IMPs and/or the antimalarial rescue medications and the event.

In addition to the assessments of relationship to the trial intervention/s, the Investigator should comment on the AE record in the eCRF whether an AE is related to the trial participation of the volunteer (trial procedures etc.).

Of note, a sign or symptom associated with malaria infection (confirmed by a positive *P. falciparum* 18S qPCR at the onset of the event) that is of expected intensity, frequency and duration for the individual volunteer in the context of this trial is considered to be an malaria challenge agent-related event.

###### 6.5.3.3 EXPECTEDNESS

An AE is regarded as an unexpected event if its nature or severity is not consistent with the applicable reference safety information (IB for the *P. falciparum* 3D7 challenge agent [20] and the IBs and/or approved manufacturer's prescribing information for marketed drugs).

Events that add significant information about the specificity, severity or frequency of previously described reactions, are also regarded as unexpected.

Expected AEs from the IMPs are listed in the Riamet® CMI and Jakavi® CMI. Expected AEs from the antimalarial rescue drugs are listed in the Malarone® CMI and Primacin® CMI.

##### 6.5.4 TIME PERIOD AND FREQUENCY FOR EVENT ASSESSMENT AND FOLLOW-UP

All AEs must be documented and followed up by an Investigator until:

- the event is resolved, or
- no further medically relevant information in relation to the event can be expected, and
- an Investigator considers it justifiable to terminate the follow-up

Events that are unresolved at the time of the volunteer's last follow-up visit should continue to be followed up by an Investigator for as long as medically indicated or until the volunteer is referred to a general practitioner or medical specialist as appropriate. The Sponsor retains the right to request additional information for any volunteer with ongoing AE(s)/SAE(s) at the end of the trial, if judged necessary.

All AEs should be treated appropriately. An Investigator will decide upon the appropriate action to be taken in response to an AE, which may include one or more of the following:

- no action taken (i.e., further observation only), or
- administration of a concomitant medication, or
- hospitalisation or prolongation of current hospitalisation (event to be reported as an SAE), or
- other.

In a case of occurrence of an SAE, regardless of whether or not it is judged to be related to any of the trial treatments or procedures, the volunteer will receive appropriate care under clinical supervision until all the symptoms of the SAE have diminished or resolved and the volunteer’s condition has improved.

For ongoing AEs, care will be provided for a period of time as specified in the clinical trial unit work instruction protocols. However, if the nature of the ongoing AE is determined by an Investigator as not being associated with any of the trial treatments or procedures, the volunteer will be advised to visit his/her general practitioner for further clinical care that might be required.

##### 6.5.5 ADVERSE EVENT REPORTING

It is Investigator’s responsibility to document and report all AEs occurring in the clinical trial whether spontaneously reported by the volunteer, observed by an Investigator (either directly or by laboratory or other assessments), or elicited by general questioning. The period of observation for collection of AEs extends from the time of inoculation with the malaria challenge agent to EOS. These AEs must be recorded on specific AE pages of the eCRF. Events reported prior to this will be recorded as medical history, unless the symptoms worsen during the trial.

The following information should be recorded for all AEs:

- Description
- Dates and times of onset and resolution
- Duration in hours
- Time of onset relative to IMP administration, inoculation with the malaria challenge agent, and/or administration of antimalarial rescue medications
- Seriousness (SAE or not)
- Severity
    - If severity of an AE changes, only one AE will be reported, with the highest severity recorded in the eCRF and listings and tables. In the clinical file (source data), the description of the AE will report the various severities observed over time.
    - If the AE resolves and then reoccurs, then two AEs will be reported.
- Action taken in response to the AE (including treatment given).
- Outcome.
- Relationship to the trial treatments or procedures (causality assessment), including malaria infection, antimalarial medication, or any other treatment or procedure conducted during the trial.

All malaria-specific AEs will be tabulated and results graded according to a purpose-designed malaria clinical score (**Appendix 2**).

##### 6.5.6 SERIOUS ADVERSE EVENT REPORTING

An Investigator will take immediate appropriate action in response to SAEs to ensure volunteer safety and in an attempt to identify the cause of the event. An Investigator will notify the Sponsor at ** of any SAE within 24 h of becoming aware of the event. The notification should be in writing by email and documented on a standard SAE reporting form.

An Investigator will complete a follow-up SAE report within 14 days of the SAE, unless no further information is available in which case the follow-up report will be provided as soon as new information becomes available. The follow-up SAE report will be sent to the same recipients as the initial report as described above. Other supporting documents may be requested by these parties and will be provided by an Investigator as soon as possible.

Any SAE that meets the criteria of a SUSAR will be reported to the TGA by the Sponsor in accordance with the Sponsor's reporting procedures.

##### 6.5.7 REPORTING EVENTS TO VOLUNTEERS

Not applicable.

##### 6.5.8 ADVERSE EVENT OF SPECIAL INTEREST (AESI)

An AE of special interest (AESI), whether serious or non-serious, is one of scientific and medical concern specific to the Sponsor's product or program, for which ongoing monitoring and rapid communication by an Investigator to the Sponsor could be appropriate. Such an event might require further investigation in order to characterise and understand it. Depending on the nature of the event, rapid communication by the trial Sponsor to other parties (e.g., regulators) might also be warranted (CIOMS VI, ICH E2F, 2010). Any abnormalities listed below should be reported as an AESI.

**Hepatic:**

- Any ALT or AST value above 5×ULN.
- An elevation in bilirubin 2×ULN.
- Any AST or ALT value above 2×ULN and (total bilirubin >1.5×ULN or INR >1.4).
- Any AST or ALT value above 2×ULN with the appearance of fatigue, nausea, vomiting, right upper quadrant pain or tenderness, fever, rash and/or eosinophilia (eosinophil percent or count above the ULN).

**Cardiac:**

- QTcF at any time >480 msec.

- Bundle branch block (except right bundle branch block that was present prior to IMP administration)
- Any arrhythmia, except:
  - sinus bradycardia that is clinically asymptomatic and not associated with any other relevant ECG abnormalities
  - sinus tachycardia that is clinically asymptomatic, is associated with a body temperature >38.0°C, and not associated with any other relevant ECG abnormalities
  - respiratory sinus arrhythmia
  - wandering atrial pacemaker, or
  - isolated single premature atrial/ventricular complex (i.e., no bigeminy, trigeminy, couplets, triplets or salvos) that does not occur more than once in a particular ECG tracing

**Haematological:**

- Haemoglobin drop >20.0 g/L from baseline (prior to inoculation with the malaria challenge agent)
- Absolute neutrophil count (ANC) <0.5 × $10^9$/L.
- Platelet count <75 × $10^9$/L.

**Dermatological:** *†

Clinical signs of possible cutaneous adverse reactions (clearly related to malaria challenge agent inoculation and/or IMP administration; this does not include irritation from dressings and/or ECG dots) such as:

- Dermatitis
- Rash including
  - erythematous
  - macular
  - papular
  - maculopapular
  - pruritic
  - pustular
  - vesicular

*If one of these cutaneous reactions is observed, pictures of the lesions should be obtained when feasible.
†Dermatological AEs need not be reported as AESIs if clearly unrelated to the malaria challenge agent or IMPs (e.g. rash from cannula dressing or ECG dots).

All AESIs, including those that do not meet the definition of an SAE, must be notified to the PI. The notification should be documented on a standard AESI reporting form. The notification of follow-up information will follow the same procedure and timelines as the initial report.

##### 6.5.9 REPORTING OF PREGNANCY

Pregnancy in a female volunteer or in a male volunteer's female partner during the trial must be reported and followed up. Pregnancy does not constitute an AE as such and the pregnancy outcome will not be recorded in the eCRF unless it is considered to be an AE.

An Investigator must notify the Sponsor in an expedited manner of any pregnancy occurring from the date of informed consent signature until 90 days after administration of IMP.

The same reporting process described for AEs and AESIs in should be followed. In all cases, the pregnancy must be followed until birth of the child, and the outcome of the pregnancy and birth reported as above by completing the appropriate Section of the Pregnancy Report Form used for the initial notification. The timelines of the reporting vary depending on the outcome as follows:

- Normal outcomes should be reported within 45 days of birth/delivery.
- Abnormal outcomes should be reported in an expedited manner as described for an SAE.

An additional SAE Report Form must be completed if the volunteer or volunteer's partner sustains an SAE, whereas a Parent-Child/Foetus Report must be completed should the child/foetus sustain an event.

#### 6.6 STATISTICAL CONSIDERATIONS

The following sections describe the statistical analysis as it is foreseen during the planning phase of trial. A detailed Statistical Analysis Plan (SAP) will be finalised and approved prior to database lock and will provide details of all analyses to be performed as well as the format of listings and tables to be provided for completion of the clinical study report (CSR). Any deviations from the SAP will be described and justified in the final CSR.

#### 6.7 STATISTICAL HYPOTHESES

We hypothesise that volunteers co-administered AL+Rux will have greater anti-parasitic immune responses of IFNγ/IL10 (Tr1) compared with the immune responses of volunteers administered with AL+placebo.

#### 6.8 SAMPLE SIZE DETERMINATION

The planned sample size in this study is 26 volunteers, with 13 volunteers randomised to AL+RUX and 13 volunteers randomised to AL+placebo. Sample size calculations were based on one of the secondary endpoints of the study, namely the anti-parasitic immune response (IFNγ and IL-10 levels) by treatment regimen. It was considered appropriate to base the sample size on this endpoint because it represents the basis of the rationale of this study, that RUX adjunctive treatment can enhance Th1 CD4+ T cells (characterised by IFNγ production) but not Tr1 regulatory CD4+ T cells (characterised by IL-10 production). Nevertheless, the planned sample size is also considered appropriate to assess the primary endpoint of this study associated with safety and tolerability, namely the incidence, severity, and relationship of observed and self-reported AEs by treatment regimen. The Australian Therapeutic Goods Administration (TGA) guidance on the design and conduct of clinical trials in Australia and suggests between 10 and 100 participants in phase 1 human pharmacology studies (https://www.tga.gov.au/sites/default/files/australian-clinical-trial-handbook.pdf). Thus the planned sample size of 26 volunteers is within this range and will allow assessment of the safety and tolerability of RUX adjunctive treatment in the context of induced blood-stage malaria.

Sample size calculations were based on data obtained from a study which measured cytokine production following *P. falciparum* parasite stimulation of peripheral blood mononuclear cells (PBMCs) collected from volunteers participating in previous *P. falciparum* induced blood stage malaria trials [42]. The natural log transformed IL-10 and IFNγ response at day 14 post-inoculation was found to be normally distributed with a mean (SD) of 6.70 pg/mL (0.472) and 6.01 pg/mL (1.20) respectively. With 13 volunteers per group in the current study, a two-sided two sample t-test will have 80% power to detect a true difference at Day 16 (168 hr after treatment) in the mean IL-10, 0.541 pg/mL which is equivalent to a 8% change from the mean of 6.70 pg/mL and IFNγ response of 1.374 pg/mL which is equivalent to 22.9% change from the mean of 6.01 pg/mL with a type I error rate of 5%.

#### 6.9 POPULATIONS FOR ANALYSES

The safety population will include all volunteers who are inoculated with the malaria challenge agent on Day 0. Other analysis populations will be defined in the SAP.

#### 6.10 STATISTICAL ANALYSES

##### 6.10.1 GENERAL APPROACH

Continuous variables will be summarised with the number of observations, mean, standard deviation, median, quartiles, minimum and maximum. Categorical variables will be summarised with the number of observations and the numbers and percent from each category.

##### 6.10.2 ANALYSIS OF THE PRIMARY ENDPOINT (SAFETY)

The primary objective is to assess the safety and tolerability of co-administration of AL+Rux to healthy volunteers in the context of *P. falciparum* IBSM. For safety analysis, the frequency tabulations of abnormal values for the parameter will be presented for treatment group by visit. Summary statistics for vital signs, body weight, laboratory assessment, and ECG will be presented for treatment group by visit. All AEs/SAEs/AESIs seen during the trial period will be listed. The frequency of AEs/SAEs/AESIs that occur will be recorded and will be assessed for causality, severity, seriousness, and expectedness.

Vital signs, routine safety laboratory data, and ECG parameters will be presented in data listings and will be summarised descriptively in tables by treatment group and by protocol specified time-points. Where applicable, both absolute values and change from baseline and inoculation will be presented and listings of clinically relevant abnormal laboratory results will be generated.

All AE data will be summarised by treatment, dose (if varied during trial), MedDRA system organ class, MedDRA preferred term, and severity. Vital signs, routine safety laboratory data, and ECG parameters will be summarised descriptively by treatment, dose (if varied during trial), and timepoint. Both absolute values and change from baseline (AL+Rux or AL+placebo administration, and inoculation with the malaria challenge agent) will be presented.

The overall number and percentage of volunteers with at least one AE/SAE/AESI will be tabulated over the entire trial period.

##### 6.10.3 ANALYSIS OF THE SECONDARY ENDPOINTS

The secondary endpoints are PK parameters, PD parameters (pSTAT3, antimalarial activity, and immune response parameters).

**Pharmacokinetic parameters**

PK parameters of artemether, DHA, lumefantrine, and Rux will be estimated using non-compartmental methods from plasma concentration-time data:

- $AUC_{last}$, $AUC_{0-\infty}$, $C_{max}$ (first and last dose), $t_{max}$ (first and last dose), elimination $t_{1/2}$, $t_{lag}$, $C_{168h}$ (for lumefantrine only), CL/F, $V_z$/F, and λz.

For calculation of descriptive statistics of plasma concentrations, values below the lower limit of quantitation will be set to LLoQ.

PK parameters will be determined using STATA® (version 14.0 or higher).

**Pharmacodynamic parameters**

***pSTAT3 levels***

The pSTAT3 inhibition ex-vivo on whole blood cells will be assessed using a validated ELISA. pSTAT3 inhibition will be expressed as a percentage. The following analyses will be conducted:

- Within-volunteer comparison of each volunteer change from baseline (pre AL+Rux or AL+placebo administration)
- Between-volunteers comparison of volunteers who receive AL+Rux and volunteers who receive AL+placebo

The percentage value of pSTAT3 inhibition may be transformed using an appropriate transformation such as a logit transformation, for analysis. Within-volunteer changes in from baseline will be assessed using paired t-test, or non-parametric tests if appropriate. Differences at specified times and in changes from inoculation with the malaria challenge agent (first inoculation) and 12 h after first administration of AL between the two treatment groups will be assessed using a two sample t-test, or non-parametric tests if appropriate.

***Antimalarial activity and parasite growth parameters***

***Parasite reduction ratio*** **(first inoculation)**

The parasite reduction ratio (PRR) of asexual parasites will be based on the decay of parasitaemia after drug treatment determined by malaria 18S qPCR. The PRR for asexual parasites will be estimated using the slope of the optimal fit of the log-linear relationship of the parasitaemia decay [43]. The optimal fit can be derived using summarised replicate parasitaemia data, which have been cleaned by dealing with potential outliers, values below the limit of detection and non-detectable values. The optimal fit of the log-linear parasitaemia-by-time relationship is determined by using left and right censoring to systematically remove the potential lag phase and tail phase of the parasitaemia decay. The decay rate, estimated by the slope coefficient from the log-linear decay regression of qPCR data, will be calculated for each volunteer. The overall treatment group specific PRR (AL+Rux and AL+placebo) will be estimated with its 95% CI by calculating the weighted average slope estimate and corresponding standard error using an inverse-variance method. Only data from volunteers who have optimal regression models with appropriate fit will contribute toward the treatment group-specific PRR. Details regarding the calculation will be in the SAP.

***Parasite clearance half-life*** **(first inoculation)**

The parasite clearance half-life ($Pt_{½}$) will be derived from the optimal decay rate. Details regarding the calculation will be in the SAP.

***Percentage of volunteers with recrudescence of parasitaemia*** **(first inoculation and second inoculation)**

The percentage of volunteers with recrudescence of parasitaemia following treatment with AL+Rux or AL+placebo will be determined by the number of volunteers who experience recrudescence. Recrudescence is determined retrospectively by visual inspection of volunteer parasitaemia data. This may include identification of different parasite lifecycle stages (e.g. 18S, *pfs25*, and *SBP-1*), and the parasitaemia curve.

***Time to parasitaemia*** **(first and second inoculation)**

Time to parasitaemia will be determined as the first time-point that all replicates of 18S qPCR are detected after the second homologous *P. falciparum* IBSM infection.

***Parasite multiplication rate*** **(first and second inoculation)**

The PMR will be calculated by applying a log-linear or sine-wave growth model to the summarised replicate parasitaemia data from patency to before treatment. Mixed-effects models will be used to obtain parameter estimates for each treatment group. The growth rate parameter from the model will be transformed into the PMR for reporting. Details regarding the calculation will be in the SAP.

Differences of antimalarial activity parameters of the parasite clearance rates ($PRR_{48}$ and parasite clearance half-life) between treatment groups will be assessed using the omnibus test, and recrudescence rates after first inoculation will be compared between groups using chi-squared (or Fisher's exact test). Time to parasitemia will be compared between groups using two-sample t-test or Kruskall Wallis test, or using a chi-squared test or Fisher's exact test if time to parasitemia is treated as a categorical variable. The PMR at the second inoculation will be compared between groups by either using an interaction term of time by group in the mixed effects model or compared using two sample t-test of the individual growth rates.

**Immune response assessments**

All volunteers who are inoculated with *P. falciparum* 3D7 challenge agent will be included in the immune response analyses for the following parameters as described in **appendix 3**:

For continuous parameters, changes to responses between baseline (prior to inoculation), and subsequent trial timepoints for each treatment group will be compared by paired t-tests or non-parametric tests.

Differences between AL+Rux and AL+placebo groups will be compared by normalising individuals' responses to baseline, and comparing magnitude and/or fold changes between groups by two sample t-test or non parametric -tests at each timepoint.

To assess differences in changes in immune parameters over time between AL+Rux and AL+placebo groups, responses will be analysed by generalised estimate equations and/or mixed effects models and differences assessed by the interaction term of time and treatment group.

Associations between proportions and fold induced immune response with pSTAT3 levels and PMR/AUC in secondary infection will be assessed by correlation analysis.

##### 6.10.4 BASELINE DESCRIPTIVE STATISTICS

Demographic data will be summarised by descriptive statistics and will include total number of observations, mean, standard deviation, and range for continuous variables that are normally distributed for medians and interquartile ranges for non-parametric continuous variables, and number and percentages for categorical variables.

Volunteer disposition will be summarised and presented in a flow diagram. Trial completion, trial withdrawals, exclusions, and violations will be summarised and the reasons for withdrawal, exclusions, and violations will be listed.

Medical history, current medical conditions, prior and concomitant medications, results of laboratory screening tests, drug and alcohol screening tests, and any other relevant baseline information will be listed by volunteer and treatment group.

##### 6.10.5 PLANNED INTERIM ANALYSES

This trial has no formal interim analyses other than review of safety, tolerability, and PK by the SDRT.
The SDRT will conduct the following reviews:

1. Review of safety, tolerability, and RBC alloantibody status from all volunteers enrolled, up to and including Day 85±7.

###### 6.10.6 SUB-GROUP ANALYSES

Not applicable.

###### 6.10.7 TABULATION OF INDIVIDUAL VOLUNTEER DATA

All individual volunteer data will be listed by measure and timepoint.

###### 6.10.8 EXPLORATORY ANALYSES

Analysis will assess waning of immune responses between treatment groups over time by assessing parameters as described above, with paired and two sample t-tests or non-parametric tests, and generalised estimate equations and/or mixed effects models

### 7 SUPPORTING DOCUMENTATION AND OPERATIONAL CONSIDERATIONS

#### 7.1 REGULATORY, ETHICAL, AND TRIAL OVERSIGHT CONSIDERATIONS

##### 7.1.1 ETHICAL STANDARD

The trial will be conducted in accordance with the protocol approved by the HREC(s), the principles of the Declaration of Helsinki (Recommendations guiding Medical Doctors in Biomedical Research Involving Human Participants, Fortaleza, Brazil 2013), the NHMRC National Statement on Ethical Conduct in Human Research (2007, updated 2018) and the Integrated Addendum to ICH E6 (R1): Guideline for Good Clinical Practice E6 (R2) (November 2016) — with introductory comments of the Australian TGA.

The PI will minimise any discomfort experienced by volunteers during the trial. The only invasive procedures will be the IV inoculation of the malaria challenge agent and the blood collection by cannulation/venepuncture. The total volume of blood drawn from each volunteer will not exceed 470 mL in any 30-day period during trial participation.

##### 7.1.2 ETHICAL REVIEW

The protocol, Participant Information Sheets, and Informed Consent Forms will be reviewed by the approving HREC(s) and no trial activities will be initiated prior to approval. All amendments and addenda to the protocol and consent forms will similarly be submitted to the approving HREC(s) for approval prior to their implementation.

Changes to the final trial protocol can only be made with the prior consent of the PI, the Sponsor and the approving HREC(s). All such changes must be attached to (or incorporated into) the final protocol and communicated to all relevant members of the clinical trial unit staff and, if appropriate, to trial volunteers. All deviations from this trial protocol will be included in the trial master file and included in the CSR. All deviations and amendments will be reported to the Sponsor at the end of each cohort. The types of amendments are discussed below.

Administrative or minor changes

Administrative or minor changes include but are not limited to changes in trial staff or contact details (e.g., Sponsor instead of contract research organisation monitors). Amendments for administrative or minor changes may be suitable for executive review (expedited) by the approving HREC(s).

Substantial amendment

Significant changes require a substantial amendment. Significant changes include but are not limited to: new data affecting the safety of volunteers, change of the objectives/endpoints of the trial, eligibility criteria, dose regimen, trial assessments or procedures, treatment or trial duration with or without the need to modify the Participant Information Sheet and Informed Consent Form. Substantial amendments are to be approved by the HREC(s). The implementation of a substantial amendment can only occur after formal approval from the Sponsor, HREC(s), and PI.

Urgent amendment

An urgent amendment might become necessary to preserve the safety of the volunteers included in the trial. The requirements for approval should in no way prevent any immediate action being taken by the PI or the Sponsor in the best interests of the volunteers. Therefore, if deemed necessary, the Investigator can implement an immediate change to the protocol for safety reasons with notification to the medical monitor as soon as practicably possible. This means that, exceptionally, the implementation of urgent amendments will occur before submission to, and approval by, the HREC(s). In such cases, the PI must notify the Sponsor within 24 hours. A related substantial amendment will be written and submitted to HREC(s) as soon as practicable but no later than 7 working days, together with a description of the steps that have already been taken in regard to implementation of this amendment.

HREC approval of future research

In the event that the PI or the Sponsor want to perform testing on the samples that is not described in the protocol, additional approval will be sought from the approving HREC(s). This may be done if a volunteer consents to blood storage for use in future research.

##### 7.1.3 INFORMED CONSENT PROCESS

###### 7.1.3.1 CONSENT AND OTHER INFORMATIONAL DOCUMENTS PROVIDED TO VOLUNTEERS

Each potential volunteer will be given the trial Participant Information Sheet and Informed Consent Form that describes in detail the trial interventions, trial procedures, and risks.

All volunteers will receive an Informed Consent for Blood Storage and an option to grant permission to be contacted about involvement in future trials.

Volunteers will be given the CMI for Riamet®, Jakavi®, Primacin® and Malarone®.

###### 7.1.3.2 CONSENT PROCEDURES AND DOCUMENTATION

During the initial screening visit/recruitment, potential volunteers will read the Participant Information Sheet. An Investigator will explain the trial via the Participant Information Sheet and the candidate volunteers will be encouraged to ask questions. Potential volunteers will be informed that participation is voluntary and that they may withdraw from the trial at any time, without prejudice. Potential volunteers will have the opportunity to discuss the trial with their family or think about it prior to agreeing to participate.

Individuals willing to be considered for inclusion in the trial will sign and date the Informed Consent Form in the presence of an Investigator. Volunteers will be given a copy of their signed Informed Consent Form. The conduct of the informed consent process will be documented in the source document (including the date) and the form will be signed before the volunteer undergoes any trial-specific procedures; only once the volunteer has consented to the trial may trial-specific screening activities commence. See Section **1.3.1** for further details.

##### 7.1.4 TRIAL DISCONTINUATION AND CLOSURE

The Sponsor, PI, approving HREC(s), and regulatory authorities independently reserve the right to discontinue the trial at any time for safety or other reasons. Circumstances that may warrant termination or suspension include, but are not limited to:

- determination of unexpected, significant, or unacceptable risk to volunteers
- demonstration of efficacy that would warrant stopping
- insufficient compliance with protocol requirements
- data that are not sufficiently complete and/or evaluable
- determination that the primary endpoint has been met
- determination of futility

This will be done in consultation with the Sponsor where practical. In the event of premature trial termination or suspension, the above-mentioned parties will be notified in writing by the terminator/suspender stating the reasons for early termination or suspension (with the exception of the Sponsor's responsibility for notifying the regulatory authorities). After such a decision, the Sponsor and the Investigator will ensure that adequate consideration is given to the protection of the volunteers' interest and safety. The Investigator must review all volunteers as soon as practical and complete all required records.

Refer to Section **5.1** for discontinuation of trial intervention.

##### 7.1.5 CONFIDENTIALITY AND PRIVACY

Volunteers will be informed that their data will be held on file by the clinical trial unit and that these data may be viewed by staff of the clinical trial unit (including, where necessary, staff of the clinical trial site other than the named Investigators).

In the event of a notifiable disease being discovered during the trial, the appropriate Public Health authorities will be notified in accordance with the Queensland Public Health Regulation 2018.

Upon request, the Investigator(s)/institution(s) will permit direct access to source data and documents for trial-related monitoring, audits, HREC review, and regulatory inspection(s) by the Sponsor (or their appropriately qualified delegates) and regulatory authorities.

Volunteers will also be informed that a report (CSR) of the trial will be submitted to the Sponsor and may also be submitted to regulatory authorities and perhaps for publication, but that they will only be identified in such reports by their trial identification number, their sex and age. The Investigators will undertake to hold all personal information in confidence.

Volunteers will be informed that samples collected for the purposes described in the protocol will be sent to the Sponsor's nominated national or international laboratory for assessment.

##### 7.1.6 FUTURE USE OF STORED SPECIMENS AND DATA

As part of the trial, safety serum retention samples will be stored indefinitely at QIMR Berghofer for retrospective safety assessments that may later be indicated. Volunteers consent to this storage and the use of the sample for safety assessments when they sign the Informed Consent Form for the trial.

For all other samples, consent must be obtained from the volunteers to store and use their samples for future research. Consent will be obtained via the Informed Consent for Blood Storage that volunteers receive during recruitment/screening. Volunteers can decide if they want their samples to be used for future research or if they want their samples destroyed at the EOS. A volunteer's decision can be changed at any time prior to the EOS by notifying the trial doctors or nurses in writing. However, if a volunteer consents to future use and some of their blood has already been used for research purposes, the information from that research may still be used.

Any future research using the stored samples that is beyond the current trial will be reviewed by the study Sponsor and the approving HREC(s). All samples will be stored at QIMR Berghofer in accordance with the laboratory SOPs. The Investigator will ensure that confidentiality will be maintained continuously in all future research that involves use of these samples. The vials containing the samples of the consented volunteers will be coded and the identifying information will not be released to any unauthorised third party. The volunteers can also choose (via the Informed Consent for Blood Storage Form) for their samples to be re-labelled with only the trial number, malaria strain, and visit timepoint. No genetic testing will be performed on the stored samples without obtaining consent from the volunteers. The stored samples will not be sold or used directly for production of any commercial product. There are no benefits to volunteers in the collection, storage, and subsequent research use of their samples. Reports about future research done with volunteer samples will NOT be kept in volunteer health records, but a volunteer's samples may be kept with the trial records or in other secure areas.

##### 7.1.7 KEY ROLES AND TRIAL GOVERNANCE

| | |
|---|---|
| Principal Investigator | A/Professor Bridget Barber, MBBS, DTM&H, MPH, FRACP, PhD<br>USC clinical Trials Centre (visiting Medical Officer) and<br>QIMR Berghofer Medical Research Institute<br>Level 5, 300C Herston Road<br>Herston Qld 4006, Australia<br>Mobile: +61 424 737 153<br> |
| Co-Investigators | Prof. James McCarthy MBBS<br>Royal Melbourne Hospital<br>Melbourne, Australia<br>Tel: +61 414 424 659<br><br><br>Indika Preethimal Leelasena RACGP<br>Health Hub Morayfield<br>19-31 Dickson Rd<br>Morayfield 4506<br>Queensland, Australia<br>Tel: +61 (0)7 54563965<br><br><br>Nischal Sahai RACGP<br>Health Hub Morayfield<br>19-31 Dickson Rd<br>Morayfield 4506<br>Queensland, Australia<br>Tel: +61 (0)7 54563965<br> |
| Study Sponsor | QIMR Berghofer Medical Research Institute<br>300 Herston Rd, Herston, QLD 4006<br>Tel: +61 (0)7 3362 0222 |
| Authorised Sponsor Signatory | Prof. Grant Ramm, PHD<br>Deputy Director<br>QIMR Berghofer Medical Research Institute |
| Sponsor Monitor | Caron Hookway<br>Independent consultant<br> |
| Institutional Ethics Committee | QIMR Berghofer Medical Research Institute Human Research Ethics Committee (QIMR Berghofer HREC; EC00278)<br>Locked Bag 2000<br>Royal Brisbane and Women's Hospital |

| | |
|---|---|
| | Brisbane Qld 4029, Australia<br>Tel: +61 (0)7 3362 0117<br> |
| Statistician | Stacey Llewellyn<br>QIMR Berghofer Medical Research Institute<br>300 Herston Rd<br>Herston QLD 4006, Australia<br>Tel: +61 (0)7 3362 0492<br> |
| Independent Medical Monitor and SDRT team Chairperson | Andrew Redmond, FRACP<br>Infectious Diseases Unit Royal Brisbane and Women's Hospital<br>Herston QLD 4029<br>Tel: +61 (0) 7 3646 8761<br> |
| Independent Malaria Expert | Professor Dennis Shanks MD<br>Australian Defence Force Malaria and Infectious Diseases Institute<br>Gallipoli Barracks, Enoggera QLD 4051, Australia<br>Tel: +61 (0)7 3332 4931<br> |
| Clinical Study Centre | USC Clinical Trials Units<br>1. Moreton Bay<br>Health Hub Morayfield<br>19-31 Dickson Road Morayfield QLD 4506 Australia<br>Tel: +61 (0)7 54563965<br><br><br>2. South Bank<br>Building A2, SW1 Complex<br>52 Merivale Street South Brisbane QLD 4101<br>Tel: +61 (0)7 5409 8630<br> |
| Laboratories | <u>Clinical laboratory measurements:</u><br>Central Laboratory<br>Tel: +61 (0)3121 4444 (24 hours)<br>Freecall: 1800 677 491<br><br><u>Parasite quantification in blood samples:</u><br>Ms. Claire Wang<br>Queensland Paediatric Infectious Diseases Laboratory<br>(Q-PID), SASVRC, Level 8, Centre for Children's Health Research<br>62 Graham Street |

| | |
|---|---|
| | South Brisbane, QLD 4101, Australia<br>Tel: +61 (0)7 3069 7462<br><br><u>Drug concentration measurements in blood samples:</u><br>Dr Mike Edstein<br>Head, Drug Evaluation at Australian Army Malaria Institute<br>Australian Defence Force Malaria and Infectious Diseases Institute<br>Gallipoli Barracks, Enoggera QLD 4051, Australia<br>Tel: +61 (0)7 3332 4931<br> |
| Serious AE and AESI reporting | QIMR Berghofer Medical Research Institute<br>300 Herston Rd, Herston, QLD 4006<br> |

##### 7.1.8 SAFETY OVERSIGHT

The SDRT will be responsible for decisions related to the safety of volunteers and the continuation of the trial. The SDRT will review the clinical and laboratory safety data and available pSTAT3 data, RBC alloantibody status, parasitaemia data, immune response data as well as the recorded AEs, AESIs, and SAEs.

Additionally, the SDRT will meet to assess any event/(s) that trigger the discontinuation and/or suspension rules or as needed to provide a recommendation and findings to the approving HREC(s) and the PI.

In addition, the SDRT has the authority to modify the design of the trial in the following ways:

a. a dose of Rux <20 mg can be recommended by the SDRT based on safety, tolerability and PK data
b. The frequency and timing of malaria 18S qPCR, PK, pSTAT3, and immune response assessment blood collection can be altered as long as blood volume is not greater than 470 mL in any 30-day period

##### 7.1.9 CLINICAL MONITORING

It will be the Sponsor's responsibility to ensure that the trial is monitored in accordance with the requirements of GCP. The conduct of the trial will be reviewed internally by the clinical trial unit in accordance with their SOPs and work instructions, and GCP guidelines. The trial will be monitored according to the SOPs of the Sponsor and all serious breaches, suspected breaches and protocol deviations will be reported to the Sponsor. Serious breaches that impact volunteer safety or data integrity will also be reported to the approving HREC.

During the trial, trial monitor(s) (on behalf of the Sponsor) will visit the clinical trial unit regularly to check the completeness of volunteer records, accuracy of eCRF entries, adherence to the protocol and to GCP, the progress of enrolment, and to ensure that trial interventions are being stored, dispensed, and accounted for according to specifications. Key trial personnel will be required to be available to assist the monitor during these visits.

The Investigator will be required to give the monitor access to all relevant source documents to confirm their consistency with the eCRF entries. At a minimum, the Sponsor will require full verification for the presence of informed consent, adherence to the inclusion/exclusion criteria, and documentation of SAEs, AESI and AEs, and the recording of data that was used for all primary and safety variables. Additional checks of the consistency of the source data with the eCRFs will be performed according to the trial-specific monitoring plan. No information captured in the source documents about the identity of the volunteers will be disclosed.

##### 7.1.10 QUALITY ASSURANCE AND QUALITY CONTROL

The clinical trial unit will perform internal quality management of trial conduct, data and biological specimen collection, documentation and completion.

Quality control procedures will be implemented beginning with the data entry system and data quality control checks that will be run on the database that will be generated. Any missing data or data anomalies will be communicated to the site(s) for clarification/resolution.

The clinical trial unit will provide direct access to source data/documents, and reports for the purpose of monitoring and auditing by the Sponsor, and inspection by regulatory authorities.

Data management, including the development and management of a secure database, will be performed in accordance with regulatory requirements. The designated data management vendor will review the data entered into the eCRFs by clinical trial unit staff for completeness and accuracy. A formal querying process will be followed whereby the data management team will request the site personnel to clarify any apparent erroneous entries or inconsistencies and will request additional information from the site as required.

Medical history/current medical conditions and AEs will be coded using the Medical Dictionary for Regulatory Activities (MedDRA) terminology (version 20.1 or higher). Prior and concomitant medications will be coded using the WHO Drug Dictionary Enhanced (WHO DDE; March 2014 or later).

After all data have been captured and reviewed, all queries have been resolved with the site, and any protocol non-compliances that were identified during the data management processes have been confirmed by the site, the database will be declared to be complete and accurate. The database will be locked and made available for data analysis. All blinded personnel may be unblinded at this time. Any changes to the database after that time may only be made by the data manager, in consultation with the Sponsor and in accordance with documented database unlock and relock procedures.

Clinical monitoring will be conducted as described in Section 7.1.9.

Audits may be carried out by Sponsor quality assurance representatives, local authorities or authorities to whom information on this trial has been submitted. All documents pertinent to this trial must be made available for such inspections after adequate notice of intention to audit.

##### 7.1.11 DATA HANDLING AND RECORD KEEPING

###### 7.1.11.1 DATA COLLECTION AND MANAGEMENT RESPONSIBILITIES

Each volunteer will have a clinical file (source data) and eCRF (for protocol specific data) into which relevant data will be recorded. All recording in source documents will be done only in black ink. Corrections will only be made by drawing a single line through the incorrect entry, writing the correction in the nearest practicable space, and initialing and dating the correction. Use of correction fluids is not allowed.

A log of names, signatures, and initials of all staff authorised to enter data into a volunteer's clinical file and eCRF will be kept. Upon completion of each trial visit, all eCRFs will be reviewed internally by the clinical trial unit for omissions or apparent errors, so that these can be corrected without delay. Any corrections made after the review and signature of the PI will be noted in the audit trail and will required reauthorisation (electronic sign off) by the PI.

###### 7.1.11.2 TRIAL RECORDS RETENTION

All source data, clinical records, and laboratory data relating to the trial will be retained in the archive of the clinical trial unit for a minimum of 15 years after the completion of the trial. Data will be available for retrospective review or audit by arrangement with the Chief Executive Officer of the clinical trial unit. Written agreement from the Sponsor must precede destruction of the same.

##### 7.1.12 PROTOCOL DEVIATIONS

<u>Protocol deviation:</u> any departure, change, and/or addition from the trial design or procedures defined in the protocol that has received approval by the competent authorities and favourable opinion from the approving HREC.

<u>Suspected serious breach:</u> a report that is judged by the reporter as a possible serious breach, but has yet to be formally confirmed as a serious breach by the Sponsor.

<u>Serious breach:</u> a breach of GCP or the protocol that is likely to affect to a significant degree: a) the safety or rights of a trial volunteer, and/or b) the reliability and robustness of the data generated in the clinical trial.

<u>Note to File (NTF):</u> a record that documents in detail actions taken, important decisions made, or explains a sequence of events where no other detailed record exists to enable the conduct of the trial to be reconstructed.

**Reporting requirements:**

- All protocol deviations will be documented in the trial master file and included in the CSR.
- All NTFs, protocol deviations, suspected serious breaches, and serious breaches will be viewed by the PI's delegate and signed by the PI.
- All protocol deviations, suspected serious breaches and serious breaches will be reported and assessed at each SDRT meeting.
- All serious breaches will be reported by the clinical trial unit to the Sponsor and the approving HREC(s) as early as possible, but within 7 days.
- All minor protocol deviations will be captured in a log and reported by the clinical trial unit to the PI's delegate and signed by the PI every 14 days.
- Minor protocol deviation logs will be submitted by the clinical trial unit to the Sponsor and approving HREC via inclusion with the annual report.

##### 7.1.13 PUBLICATION AND DATA SHARING POLICY

The data management, statistical, and medical writing team appointed by the Sponsor will collaborate to provide a detailed CSR upon conclusion of the trial. The CSR will include appendices of all tables and listings generated during the analyses of data. The tables and listings will be provided by the Sponsor. The Sponsor will undertake to ensure that all safety observations made during the conduct of the trial are documented in the CSR.

Publication and reporting of results and outcomes of this trial will be accurate and honest, and undertaken with integrity and transparency. The Sponsor recognises that QIMR Berghofer and the PI have a responsibility to ensure that results of scientific interest arising from the trial are appropriately published and disseminated. Publication of results will be volunteered to fair peer-review. Authorship will be given to all persons providing significant input into the conception, design, and execution or reporting of the research according to the QIMR Berghofer Policy on the Criteria for Authorship 2018. No person who is an author, consistent with this definition, will be excluded as an author without his/her permission in writing. Authorship will be discussed between researchers prior to trial commencement (or as soon as possible thereafter) and reviewed whenever there are changes in participation. Acknowledgement will be given to collaborating institutions and hospitals and other individuals and organisations providing finance or facilities. All disputes about authorship will be reviewed by the QIMR Berghofer Director.

The Sponsor will ensure that the key design elements of this protocol are posted in a publicly accessible database such as Australian New Zealand Clinical Trials Registry (ANZCTR) or Clinicaltrials.gov. In addition, upon trial completion and finalisation of the CSR, the results of this trial will be submitted for publication in an open access journal and/or posted in a publicly accessible database of clinical trial results.

##### 7.1.14 CONFLICT OF INTEREST POLICY

Bridget is named as a CIB on the awarded NHMRC Ideas Grant, which wil fund the clinical trial.

#### 7.2 LIABILITY/INDEMNITY/INSURANCE

The Sponsor will ensure sufficient insurance is available to enable it to indemnify and hold the Investigators and relevant staff as well as any hospital, institution, ethics committee or the like, harmless from any claims for damages for unexpected injuries, including death, that may be caused by the volunteer's participation in the trial but only to the extent that the claim is not caused by the fault or negligence of the volunteer(s) or Investigators. The Sponsor adheres to the guidelines of Medicines Australia for injury resulting from participation in a company sponsored trial, including the provision of 'No-fault clinical trial insurance'.

#### 7.3 ABBREVIATIONS

| | |
|---|---|
| AE | adverse event |
| AESI | adverse event of special interest |
| AL | artemether/lumefantrine (Riamet® in this trial) |
| ALT | Alanine aminotransferase |
| ANC | absolute neutrophil count |
| APTT | Activated Partial Thromboplastin Time |
| AST | Aspartate aminotransferase |
| BDI | Beck Depression Inventory |
| b.i.d | twice-daily (medication) |
| CMI | Consumer Medication Information |
| CRP | C-reactive protein |
| CSR | Clinical study report |
| CTCAE | Common Terminology Criteria for Adverse Events |
| DHA | dihydroartemisinin |
| ECG | electrocardiograph |
| eCRF | electronic Case Report Form |
| EOS | End of study |
| FDA | (US) Food & Drug Administration |
| FSH | Follicle stimulating hormone |
| G6PD | glucose-6-phosphate dehydrogenase |
| GCP | Good Clinical Practice |
| GMP | Good Manufacturing Practice |
| HREC | Human Research Ethics Committee |
| IB | Investigator's Brochure |
| IBSM | induced blood stage malaria |
| ICH | International Conference on Harmonisation |
| ICMJE | International Committee of Medical Journal Editors |
| IL-10 | interleukin 10 |
| IMP | Investigational medicine product |
| INR | International Normalised Ratio |
| IV | intravenous |
| LFT | liver function test |
| LSLV | last volunteer, last visit |
| MCB | master cell bank |
| MedDRA | Medical Dictionary for Regulatory Activities |
| NTF | Note to file |
| PBMC | peripheral blood mononuclear cell |
| $Pt_{½}$ | parasite clearance half-life |
| PD | Pharmacodynamics |
| PI | Principal Investigator |
| PK | Pharmacokinetics |

| | |
|---|---|
| PMR | Parasite multiplication rate |
| PRR | Parasite reduction rate |
| pSTAT3 | phospho-signal transducer and activator of transcription 3 |
| PT | Prothrombin Time |
| qd | one a day (medication) |
| Q-Gen | Q-Gen Cell Therapeutics |
| qPCR | quantitative polymerase chain reaction |
| RBC | Red blood cell |
| Rux | Ruxolitinib (Jakavi® in this trial) |
| SAE | Serious Adverse Event |
| SAP | Statistical Analysis Plan |
| SDRT | Safety Data Review Team |
| SIV | Site initiation visit |
| SoA | Schedule of Activities |
| SOP | Standard Operating Procedure |
| SUSAR | Suspected Unexpected Serious Adverse Reaction |
| Tfh | T follicular helper (cell) |
| TGA | (Australian) Therapeutic Goods Administration |
| ULN | Upper limit of normal |
| WOCBP | Women of childbearing potential |

41. Australian Government Department of Health Therapeutic Goods Administration, *Australian clinical trial handbook: Guidance on conducting clinical trials in Australia using 'unapproved' therapeutic goods. Version 2.2*. 2018: Woden, ACT.

42. Montes de Oca, M., et al., *Type I interferons regulate immune responses in humans with blood-stage Plasmodium falciparum infection.* Cell Rep, 2016. **17**(2): p. 399-412.

43. Marquart, L., et al., *Evaluating the pharmacodynamic effect of antimalarial drugs in clinical trials by quantitative PCR.* Antimicrob Agents Chemother, 2015. **59**(7): p. 4249-59.

### 9 APPENDICES

#### Appendix 1: Sponsor Approved Clinically Acceptable Trial Inclusion Laboratory Ranges

| Test | Unit | Laboratory Normal Ranges | | Acceptable Inclusion Range | | Gender acceptable inclusion range | |
|---|---|---|---|---|---|---|---|
| | | Male | Female | Low | High | Male | Female |
| **Biochemistry** | | | | | | | |
| Sodium | mmol/L | 135-145 | 135-145 | 130 | 150 | | |
| Potassium | mmol/L | 3.5-5.5 | 3.5-5.5 | 3.0 | 5.5 | | |
| Chloride | mmol/L | 95-110 | 95-110 | 85 | 120 | | |
| Calcium (Corr) | mmol/L | 18 years: 2.20-2.65 | 18 years: 2.20-2.65 | 2.05 | 2.75 | | |
| | | 19-55 years: 2.10-2.60 | 19-55 years: 2.10-2.60 | 2.05 | 2.67 | | |
| Urea | mmol/L | 18-29 years: 3.0-7.5 | 18-29 years: 2.5-6.5 | N/A | 1.75xULN | N/A – 13.1 | N/A – 11.4 |
| | | 30-49 years: 3.0-8.0 | 30-49 years: 2.5-7.0 | N/A | 1.75xULN | N/A – 14.0 | N/A – 12.3 |
| | | 50-55 years: 3.5-8.5 | 50-55 years: 3.0-8.0 | N/A | 1.75xULN | N/A – 14.9 | N/A – 14.0 |
| Urate | mmol/L | 0.20-0.50 | 0.15-0.40 | N/A | 1.75xULN | N/A – 0.88 | N/A – 0.70 |
| Creatinine | umol/L | 60-110 | 45-85 | N/A | 1.0xULN | N/A – 110 | N/A – 85 |
| Creatine kinase | U/L | 45-250 | 30-150 | N/A | ≤5.0xULN | ≤1250 | ≤750 |
| eGFR | mL/min/1.73m$^2$ | ≥59 | ≥59 | N/A | 1.55xULN<br>91 | | |
| Glucose Fasted | mmol/L | 3.6-6.0 | 3.6-6.0 | N/A | 1.0xULN<br>6 | | |
| Total Bilirubin | umol/L | 4-20 | 3-15 | N/A | 1.25xULN | N/A – 25 | N/A – 19 |
| ALP | U/L | 18-19 years: 60-200 | 18 years: 45-120 | N/A | 1.5xULN | N/A – 300 | N/A – 180 |
| | | 20-55 years: 35-110 | 19-49 years: 20-105 | N/A | 1.5xULN | N/A – 165 | N/A – 158 |

| Test | Unit | Laboratory Normal Ranges | | Acceptable Inclusion Range | | Gender acceptable inclusion range | |
|---|---|---|---|---|---|---|---|
| | | Male | Female | Low | High | Male | Female |
| | | | 50-55 years: 30-115 | N/A | 1.5xULN | | N/A – 173 |
| AST | U/L | 10-40 | 10-35 | N/A | 1.25xULN | N/A – 50 | N/A – 44 |
| ALT | U/L | 5-40 | 5-30 | N/A | 1.25xULN | N/A – 50 | N/A – 38 |
| GGT | U/L | 5-50 | 5-35 | N/A | 1.5xULN | N/A – 75 | N/A – 53 |
| Cholesterol | mmol/L | 3.9-5.5 | 3.9-5.5 | N/A | 6.5 | | |
| HDL Cholesterol | mmol/L | 0.9-1.5 | 1.1-1.9 | 0.8 | N/A | | |
| LDL Cholesterol | mmol/L | 0-4 | 0-4 | N/A | 5.4 | | |
| **Haematology** | | | | | | | |
| Hb | g/L | 135-175 | 115-165 | | | 130-180 | 110-170 |
| Plats | $x10^9/L$ | 150-400 | 150-400 | 145 | 450 | | |
| WCC | $x10^9/L$ | 3.5-10.0 | 3.5-12.0 | 3.5 | 13.2 | | |
| Neuts | $x10^9/L$ | 1.5-6.5 | 1.5-8.0 | 1.5 | 8.8 | | |
| Lymphs | $x10^9/L$ | 1.0-4.0 | 1.0-4.0 | 1.0 | 4.4 | | |
| **Urine** | | | | | | | |
| Protein (dipstick) | | N/A | N/A | N/A | 30mg/dL | | |
| Ketones (dipstick) | | N/A | N/A | N/A | 80mg/dL | | |
| Red Blood Cells (MCS) | | <10 | <20 | | | <20 | <20* |
| White Blood Cells (MCS) | | <10 | <10 | N/A | <10 | | |
| Casts (MCS) | | Not seen | Not seen | N/A | <2/high power field | | |

* Results >20 are NCS if female volunteer is menstruating.

**Appendix 2: Malaria Clinical Score**

The malaria clinical score gives an indication of the severity of the induced malaria infection in each volunteer. Fourteen signs/symptoms frequently associated with malaria are graded using a 4-point scale (absent 0; mild: 1; moderate: 2; severe: 3) and summed to generate a total malaria clinical score (maximum score possible is 42). Severity will be graded in accordance with the CTCAE Version 5.0 Published: November 27, 2017. Mild (1) equates to CTCAE grade 1, Moderate (2) equates to CTCAE grade 2 and Severe (3) equates to CTCAE grade 3. The criteria are shown in the table below.

| Symptom or sign | Clinical Score/CTCAE grade | | | |
|---|---|---|---|---|
| | Absent (0) | Mild (1) | Moderate (2) | Severe (3) |
| | | CTCAE 1 | CTCAE 2 | CTCAE 3 |
| Headache | | Mild pain | Moderate pain; limiting instrumental activities of daily living (ADL) | Severe pain; limiting self-care ADL |
| Myalgia | | Mild pain | Moderate pain; limiting instrumental ADL | Severe pain; limiting self-care ADL |
| Arthralgia | | Mild pain | Moderate pain; limiting instrumental ADL | Severe pain; limiting self-care ADL |
| Fatigue | | Fatigue relieved by rest | Fatigue not relieved by rest; limiting instrumental ADL | Fatigue not relieved by rest; limiting self-care ADL |
| Malaise | | Uneasiness or lack of well-being | Uneasiness or lack of well-being; limiting instrumental ADL | Uneasiness or lack of well-being limiting self-care ADL |
| Chills | | Mild sensation of cold; shivering; chattering of teeth | Moderate tremor of the entire body; narcotics indicated | Severe or prolonged, not responsive to narcotics |
| Sweating/hot spells | | Mild sweating/hot spells not affecting ADL | Moderate sweating/hot spells; narcotics indicated | Severe or prolonged, not responsive to narcotics |
| Reduced appetite | | Loss of appetite without alteration in eating habits | Oral intake altered without significant weight loss or malnutrition; oral nutritional supplements indicated | Associated with significant weight loss or malnutrition (e.g. inadequate oral caloric and/or fluid intake); tube feeding or TPN indicated |

| Symptom or sign | Clinical Score/CTCAE grade | | | |
|---|---|---|---|---|
| Nausea | | Loss of appetite without alteration in eating habits | Oral intake decreased without significant weight loss, dehydration or malnutrition | Inadequate oral caloric or fluid intake; tube feeding, TPN, or hospitalisation indicated |
| Vomiting | | Intervention not indicated | Outpatient IV hydration; medical intervention indicated | Tube feeding, TPN, or hospitalisation indicated |
| Abdominal discomfort | | Mild pain | Moderate pain; limiting instrumental ADL | Severe pain; limiting self-care ADL |
| Fever | | 38.0-38.9°C | ≥39.0-39.9°C | ≥40.0°C |
| Tachycardia | | HR ≥100 Asymptomatic, intervention not indicated | HR ≥100 Symptomatic; non-urgent medical intervention indicated | HR ≥100 Urgent medical intervention indicated |
| Hypotension | | SBP ≤80 Asymptomatic, intervention not indicated | SBP ≤80 Symptomatic; non-urgent medical intervention indicated | SBP ≤80 Urgent medical intervention indicated |
| Total Score | | | | |
| Maximum 3 x 14 = 42 | | | | |

**Appendix 3. Immune response assessment assays.**

| Samples and Assay | Details |
|---|---|
| **Plasma antibodies** | • Measure anti-parasitic antibody levels and immunoglobulin isotypes.<br>• Measure anti-parasitic antibody functional capacities, including ability to inhibit invasion, fix compliment, bind FcγRII and FcγRIII, and mediate opsonic phagocytosis by monocytes. |
| **Plasma cytokines** | • Measure levels of cytokines, including IFNγ, TNF, IL-10, IL-6, IL-1β, IL-17, IL-5, IL-4 and IL-8. |
| **Flow cytometry on peripheral blood mononuclear cells (PBMCs)** | • $CD4^+$ T cells, including Th1, Tr1, Th2, Th17, Tfh and Treg cells, as well as activation markers and cytokines, including IFNγ, IL-10, Ki67, PD1, LAG3, CTLA4, CD45RA, CD127, CD45RO, CCR7, CD62L, CD25, ICOS, CD38 and CD95.<br>• $CD8^+$ T cells, including activation and phenotypic markers, including IFNγ, IL-10, Ki67, PD1, LAG3, CTLA4, CD45RA, CD127, CD45RO, CCR7, CD62L, CD25, CD95, granulysin, granzyme B and perforin.<br>• B cells, including antibody secreting cells, plasmablasts and memory B cells, as well as activation markers, including CD38, GL7 and antibody isotype switching.<br>• Innate-like lymphoid cells, including NK, NKT, γδT and MAIT cells, as well as activation markers, including IFNγ, IL-10, PD1, LAG3, CTLA4, CD45RA, CD127, CD45RO, CCR7, CD62L, CD25, ICOS, CD38 and CD95.<br>• Innate cells, including dendritic cells (DCs), macrophages, monocytes and neutrophils, as well as activation markers, including IL-6, TNF, CD80, CD86 and HLA-DR. |
| **Antigen recall responses for PBMCs** | • Culture PBMCs in the presence of parasite antigen for 24 and 72 hours and measure cytokine production, including IFNγ, TNF, IL-10, IL-6, IL-1β, IL-17, IL-5, IL-4 and IL-8.<br>• Culture PBMCs in the presence of parasite antigen for 24 and 72 hours and measure $CD4^+$ T cell proliferation by CFSE dilution and intracellular cytokine production, including IFNγ, TNF and IL-10. |
| **Cryopreservation of PBMCs** | Cells cryopreserved for future assays to be determined based on results from above, including cell subset isolation and RNAseq, CRISPR/Cas9 gene modifications, proteomics and metabolomics studies. |
